# Economic and social impacts of COVID-19 and public health measures: results from an anonymous online survey in Thailand, Malaysia, the United Kingdom, Italy and Slovenia

**DOI:** 10.1101/2020.10.26.20209361

**Authors:** Anne Osterrieder, Giulia Cuman, Wirichada Pan-ngum, Phaik Kin Cheah, Phee-Kheng Cheah, Pimnara Peerawaranun, Margherita Silan, Miha Orazem, Ksenija Perkovic, Urh Groselj, Mira Leonie Schneiders, Tassawan Poomchaichote, Naomi Waithira, Supa-at Asarath, Bhensri Naemiratch, Supanat Ruangkajorn, Lenart Skof, Natinee Kulpijit, Constance R.S. Mackworth-Young, Darlene Ongkili, Rita Chanviriyavuth, Mavuto Mukaka, Phaik Yeong Cheah

**Affiliations:** Mahidol Oxford Tropical Medicine Research Unit, Faculty of Tropical Medicine, Mahidol University, Bangkok, 10400, Thailand; Centre for Tropical Medicine & Global Health, Nuffield Department of Medicine, University of Oxford, Oxford, UK; Paediatric Ethics Committee; Research Ethics Committee, University Hospital of Padua, Padua, Italy; Department of Tropical Hygiene, Faculty of Tropical Medicine, Mahidol University, Bangkok, 10400, Thailand; Faculty of Arts and Social Science, Universiti Tunku Abdul Rahman, Kampar, Malaysia; Emergency and Trauma Department, Sabah Women and Children’s Hospital, Ministry of Health Malaysia, Kota Kinabalu, Malaysia; Department of Statistical Sciences, University of Padua, Padua, Italy; Faculty of Medicine, University of Ljubljana, Ljubljana, Slovenia; Department of Radiation Oncology, Institute of Oncology Ljubljana, Ljubljana, Slovenia; Institute for Social Studies, Science and Research Centre Koper, Koper, Slovenia; Department of Endocrinology, Diabetes and Metabolic Diseases, University Children’s Hospital, University Medical Center – University Children’s Hospital Ljubljana, Ljubljana, Slovenia; Ethox Centre, Nuffield Department of Population Health, University of Oxford, Oxford, UK; The SoNAR-Global Network; Institute for Philosophical Studies, Science and Research Centre Koper, Koper, Slovenia; Department of Global Health and Development, London School of Hygiene and Tropical Medicine, London, UK; Emergency and Trauma Department, Queen Elizabeth Hospital, Ministry of Health Malaysia, Kota Kinabalu, Malaysia

**Keywords:** COVID-19, SARS-CoV-2, non-pharmaceutical interventions, restrictions, social distancing, quarantine, isolation, compliance, information, fake news, social, ethical, behavioural, survey

## Abstract

In the absence of a vaccine and widely available treatments for COVID-19, governments have relied primarily on non-pharmaceutical interventions to curb the pandemic. To aid understanding of the impact of these public health measures on different social groups we conducted a mixed-methods study in five countries (‘SEBCOV - Social, ethical and behavioural aspects of COVID-19’). Here we report the results of the SEBCOV anonymous online survey of adults.

Overall, 5,058 respondents from Thailand, Malaysia, the United Kingdom, Italy and Slovenia completed the self-administered survey between May and June 2020. Post-stratification weighting was applied, and associations between categorical variables assessed.

Among the five countries, Thai respondents appeared to have been most, and Slovenian respondents least, affected economically. Overall, lower education levels, larger households, having children under 18 in the household, being 65 years or older and having flexible/no income were associated with worse economic impact. Regarding social impact, respondents expressed most concern about their social life, physical health, and mental health and wellbeing.

There were large differences between countries in terms of voluntary behavioural change, and in compliance and agreement with COVID-19 restrictions. Overall, self-reported compliance was higher among respondents reporting a high understanding of COVID-19. UK respondents felt able to cope the longest and Thai respondents the shortest with only going out for essential needs or work, with 60% and 26% respectively able to cope with 29 days or longer. Many respondents reported seeing news that seemed fake to them, the proportion varying between countries, and with education level and self-reported levels of understanding of COVID-19.

Our data showed that COVID-19 public health measures have uneven economic and social impacts on people from different countries and social groups. Understanding the factors associated with these impacts can help to inform future public health interventions and mitigate their negative consequences on people’s lives.

**Summary:** *What is already known?:*

- COVID-19 public health measures and lockdowns most negatively affect those who are socio-economically disadvantaged.
- Misinformation about COVID-19 is widespread.

*What are the new findings?:*

- In the countries in which we conducted our survey, lower education levels, larger households, having children under 18 in the household, being 65 years or older and having flexible/no income were associated with worse economic impact.
- There were large differences between countries in terms of voluntary change of behaviour, as well as compliance and agreement with COVID-19 related public health measures.
- Younger age and lower education levels appear to be associated with lower self-perceived levels of understanding of COVID-19.
- A significant proportion of the population received conflicting information and news that seemed fake to them, in particular about coronavirus being an engineered modified virus.

*What do the new findings imply?:*

- Our findings imply that there are significant differences in how people from different social groups and different countries experienced COVID-19 and related public health measures, and any support initiatives should take this into account.
- Our findings confirm that communication around COVID-19 could be improved, and help identify specific areas to target (e.g. origin of virus) and specific groups of people who may benefit most from improved communication (e.g. younger people, those with lower levels education).

## Introduction

COVID-19 is a respiratory disease caused by the novel coronavirus ‘severe acute respiratory syndrome coronavirus 2’ (SARS-CoV2), which is transmitted through droplets, close contact, and aerosols^1,2^. The SARS-CoV2 outbreak was first reported in December 2019 in Wuhan, China^3^, with the World Health Organization declaring it Public Health Emergency of International Concern on 30^th^ January 2020 and a global pandemic on 11^th^ March 2020^1^.

In the absence of a vaccine or widely available and effective pharmaceutical treatments, the impact of COVID-19 is being mitigated using non-pharmaceutical interventions (NPIs)^4,5^. Examples of NPIs include: social distancing (or ‘physical distancing’) measures, such as isolation of sick individuals, quarantine of exposed individuals, contact tracing, voluntary shielding, avoiding crowds, or closures of workplaces and schools; travel-related restrictions; and personal protective measures, such as hand hygiene and wearing face masks^4,6,7^. Scientific evidence indicates that NPIs are effective measures to contain a pandemic and ease pressures on health care systems^6-12^. However, authorities and policy makers need to consider the societal, economic and ethical impacts of these public health measures, in particular on vulnerable groups^13,14^. Such groups might be disproportionally affected by NPIs and/or might be unable to comply with them^15^, e.g. due to loss of income when having to isolate at home, crowded living conditions^14^, or not being able to afford masks^16^.

As the COVID-19 pandemic continues, evidence is urgently needed to understand how people perceive and experience NPIs, which groups are disproportionally negatively affected by NPIs, and how communication is perceived by various social groups^17^. This understanding is important so that the policies can be improved to minimize the negative impact of COVID-19 on people’s lives, and to improve communications.

Here we report the highlights of an online survey conducted in Southeast Asia (Thailand and Malaysia, both upper middle-income countries), and Europe (the United Kingdom, Italy and Slovenia, all high-income countries) between May 1 to June 30, 2020 as part of the mixed-methods study ‘Social, ethical and behavioural aspects of COVID-19’ (SEBCOV)^18^. These findings help to address an evidence gap as identified by the global research community in a recent study on COVID-19 research priorities^19^, which identified public health messaging, compliance and trust in public health interventions, and evaluation of these interventions in varied settings as areas of high priority^19^.

### Methods

#### Survey development

The survey contained five sections with 36 questions (single-answer multiple choice and five-point Likert scales) on (1) socio-demographic information; (2) income, occupation status and economic impacts of COVID-19 restrictions; (3) sources of, preferences and perceptions regarding COVID-19 related communication, and the occurrence of ‘fake news’ (untrue information presented as news); and (4) perceived levels of understanding of COVID-19 and NPIs, agreement with NPIs, voluntary behavioural changes, and concerns and coping strategies relating to restrictions^20^. The Malaysia and UK surveys were administered in English, with the other surveys translated into the respective country languages. The self-administered online survey was set up using the ‘JISC Online surveys’ platform^21^.

### Public involvement

The survey questions were pilot-tested with 25 people from participating countries, and revised accordingly based on feedback. In addition, the Bangkok Health Research Ethics Interest Group, a public involvement group set up by the Mahidol Oxford Tropical Medicine Research Unit (MORU)^22^, discussed the study and the survey questions in a dedicated virtual meeting. Selected questions were tested using an adapted cognitive testing technique using the “thinking out loud” approach^23^, and the collaborative virtual sticky notes board ‘Padlet’^24^.

### Participant selection and recruitment

Adults of any age residing in Thailand, Italy, Malaysia, United Kingdom (UK) or Slovenia at the time of the study were eligible to take part. Participants needed to be able to use a computer or smart phone to access the survey and provide online consent to participate.

The survey was open from 1^st^ May to 30^th^ June 2020 (1^st^-30^th^ June for Slovenia due to late start). Participants were recruited using a combination of approaches: snowball sampling through personal and professional networks (via email, social media and messenger apps, mailing lists, and organisations such as the Medical Chamber^25^ in Slovenia); a polling company^26^ in Thailand; and through promoted posts on Facebook. Facebook allows users to ‘boost’ posts to selected demographic audiences for a small fee, so that the post appears on their Facebook newsfeed^27^. To achieve more balanced responses in the categories of gender, education level and geographic distribution, promoted Facebook posts were targeted at people with primary or lower/secondary education in UK and Malaysia; potential participants in Wales, Scotland and Northern Ireland in the UK; and at men in the UK and Italy.

### Sample size

Each country aimed to recruit a minimum sample of 600 respondents, exceeding the 40-200 respondents recommended for a mixed-methods study^28^. A minimum sample size of 600 respondents is adequate to estimate the prevalence of a response assuming a 50% prevalence rate, with 95% confidence and with a precision of 4%. The 50% prevalence is the standard assumption for precision sample size calculations when the true prevalence is not available, as this gives the highest sample size for a binomial distribution for a desired level of precision.

### Statistical analysis

To simplify analysis, answers in the following categories were combined as follows: “slightly agree/highly agree” were combined into one “agree”, category, and “slightly/strongly disagree” responses into one “disagree” category (Suppl. Tables 23-27). To understand the distribution of the basic demographic variables in the respondent sample, the observed frequencies and sample characteristics are reported using unweighted percentages (Suppl. Table 1). The characteristics for the rest of the variables are presented using the observed survey frequency counts followed by weighted percentages (Suppl. Tables 2-37). Post-stratification weighting was used to align the composition of the respondents’ sample with the known distribution of the whole population’s characteristics, reducing sampling error. Weights were computed considering three stratifying variables that were available from population census data from each country^29^, namely, gender, age and education level. Weights were obtained as the ratio between the proportion of each possible combination of the three variables in the whole country population and the correspondent proportion in the respondent sample.

Survey data was analysed using Stata 15.0 software^30^. Frequency counts and percentages were used to summarise categorical data. Associations between categorical variables were assessed using Pearson’s Chi-squared test. P-values have been provided in the tables and considered statistically significant below the two-sided alpha=0.05 level. All p-values presented in the tables are for global tests of significance. Practical significance was taken into account when interpreting differences in the results.

## Results

At the time of the inception of this study, governments in Thailand, Malaysia, Italy, the UK and Slovenia had initiated public health measures, using varying degrees of “lockdowns” to curb the pandemic. Figure 1 shows a visualization of the ‘Stringency Index’ (SI) of the public health responses of the five government over the study period, drawing upon data provided by the Oxford COVID-19 Government Response Tracker (OxCGRT)^31^. The OxCGRT tool tracks government policies and interventions from more than 180 countries on standardized indicators, and aggregates the data into a ‘Stringency Index’ for each country on a scale from 0-100, with 100 indicating the strictest response^31^. For example, Italy had the strictest public health measures in early May (SI = 93) and then gradually lifted and reintroduced restrictions, whereas restrictions in the UK remained at around the same level (SI = 69-76) throughout the study period. Restrictions in Slovenia were substantially eased from June onwards (SI = 33).

**Figure 1:**
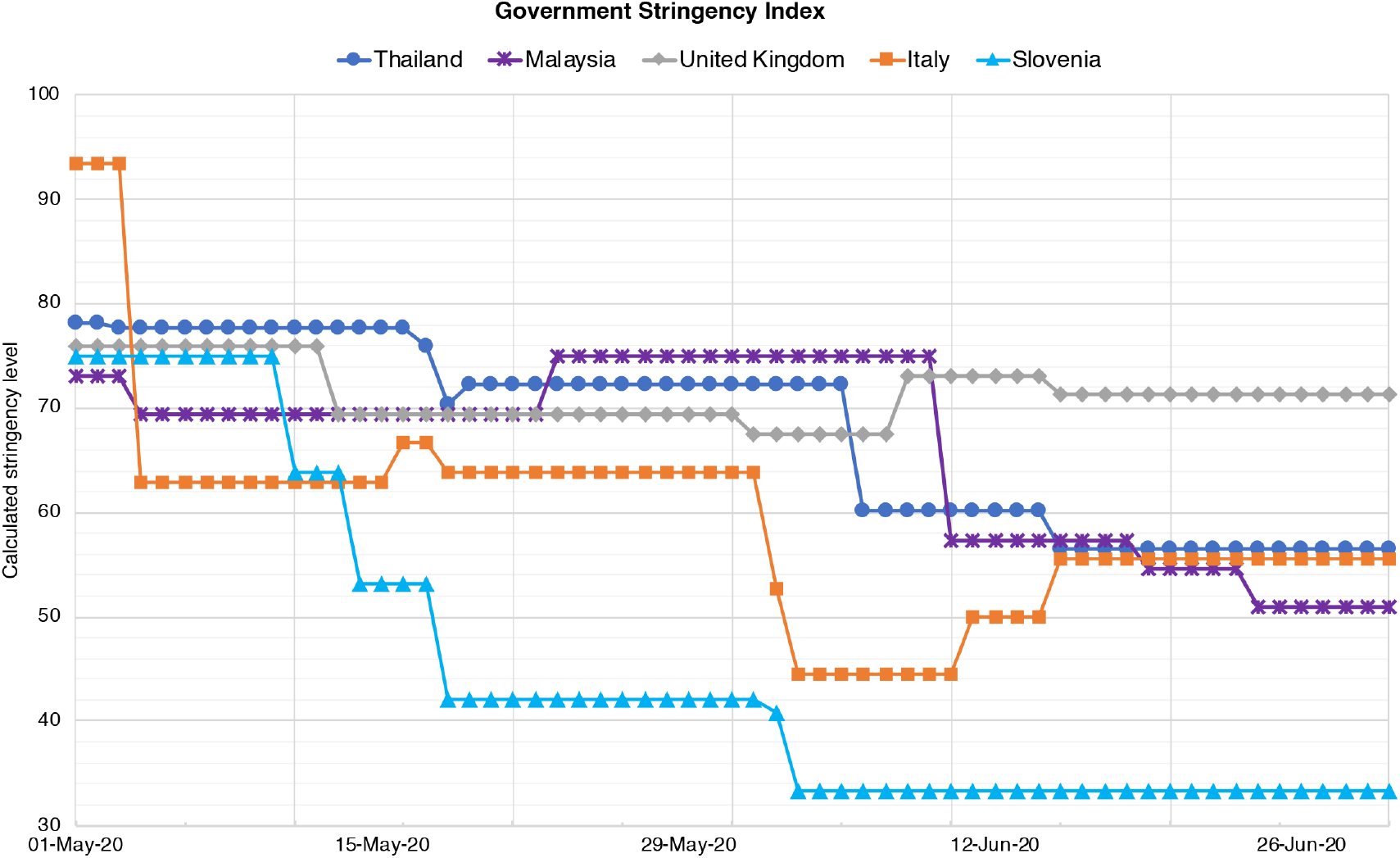
Government stringency indices in Thailand, Malaysia, UK, Italy and Slovenia between 1^st^ May – 30^th^ June 2020. A higher score indicates a stricter government response, i.e. 100 = strictest^31^.

### Characteristics of survey respondents

A total of 5,058 participants took part in the survey: 1,476 respondents from Thailand, 827 from Malaysia, 1,009 from the UK, 712 from Italy, and 1,034 from Slovenia (Suppl. Table 1, unweighted data). Overall, around 40% identified as male, around 60% as female, and around 1% as ‘other/prefer not to say’. Of all respondents, 26% were aged 18-34 years old, 65% were 35-64 years old, and 10% fell into the 65+ age group. Thirty three percent had primary or lower (from here on referred to as ‘primary’) or secondary education, whereas 67% had tertiary education. Overall, 10% of respondents lived in large households with six or more people. Fifty nine percent of respondents received a fixed income (salary/benefits/pension), 31% had flexible income (contract and freelance), and 10% received no or ‘other income’. Thirty six percent lived with children under 18 years in their household, and 29% reported that they or a household member belonged to a “vulnerable group” (persons aged 70 or older, pregnant women, or people with serious health conditions). Nineteen percent of respondents were healthcare provider/workers. Supplementary Table 1 provides the breakdown by country. All results in the following subsections are presented as weighted percentages.

### Economic impacts of COVID-19 and public health measures

In order to understand the economic impacts of COVID-19, respondents who had been working before the pandemic (paid or unpaid work) were asked whether COVID-19 had created any work-related inconvenience for them. Overall, 56% of respondents said that they experienced loss of earnings, 44% reduction of working hours, 36% closure of workplace and 14% job loss (Fig. 2, Suppl. Table 2). Seventy five percent reported that they continued to work during COVID-19. Of all respondents, 53% expressed financial concerns, and 32% worried about professional/career progression. Our results indicated that the most affected country was Thailand, with 85% of respondents reporting loss of earnings, 23% loss of job, and 86% expressing financial concerns (Suppl. Table 2). Slovenian respondents reported the least severe economic impacts e.g. 30% reported loss of earnings, 3% reported loss of job, and 28% had financial concerns.

**Figure 2:**
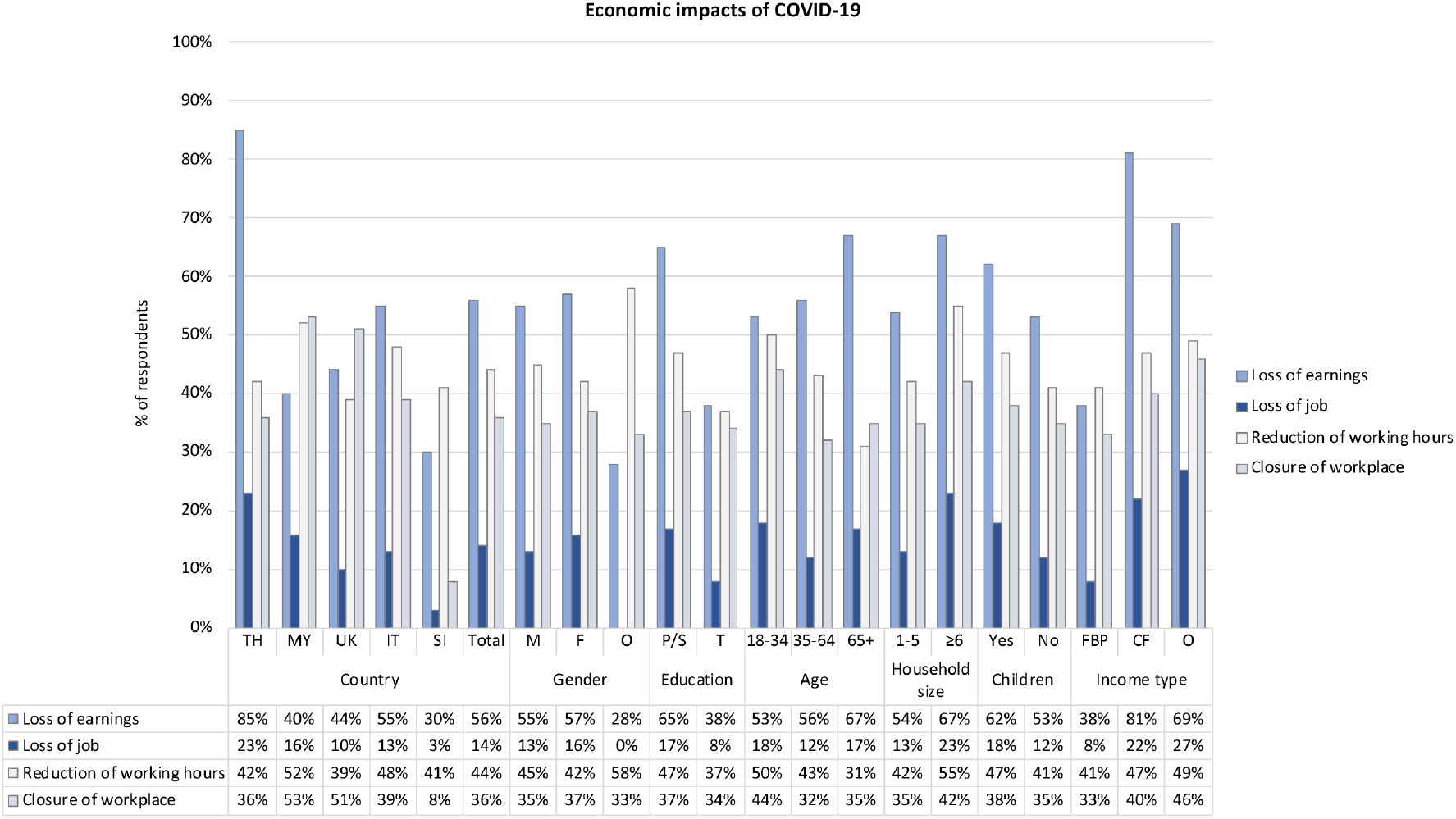
Bar chart showing how respondents from the following demographic groups were affected economically by COVID-19: at country level (TH = Thailand, MY = Malaysia, UK = United Kingdom, IT = Italy, SI = Slovenia), gender (M = male, F = female, O = Other/prefer not to say); education level (P/S = primary or lower/secondary, T = tertiary); age (18-34 years old, 35-64 years old, 65+ years old); household size (1-5 people, ≥6 people); living with children under 18 years (Y = yes, N = no); and type of income (FBP = fixed/benefits/pension, CF = contract/freelance, O = other/no income).

To investigate the impact of public health measures on different social groups, we analyzed responses based on gender, level of education, age group, household size, whether respondents lived with children under 18 years old, and income type.

Overall, there were no significant differences between male, female and respondents who identified as ‘other/prefer not to say’, and who had been working before COVID-19, in terms of loss of earnings, loss of job, reduction of working hours and closure of workplace (Fig. 2, Suppl. Table 3). Overall, fewer women continued to work during COVID-19 (71% women vs 78% men; p=0.010). The trend was similar at country level, except for Malaysia (73% women versus 67% men; Suppl. Table 3).

Overall, 65% of respondents with primary and secondary education who had been working before COVID-19 reported a loss of earnings, compared to 38% of respondents with tertiary education (p<0.001; Fig. 2, Suppl. Table 4). More respondents with primary/secondary education lost their job (17% versus 8%; p<0.001), and had their working hours reduced (47% versus 37%; p<0.001). Fewer respondents with primary/secondary education continued to work (71%, versus 83%, p<0.001), and 59% reported financial concerns (versus 41%; p<0.001). This trend was mirrored at country level. Respondents with primary/secondary education were most affected in Thailand, where 90% reported loss of earnings, 24% reported loss of job, and 89% reported financial concerns (Suppl. Table 4). Only 65% of respondents with primary/secondary education in Malaysia (versus 90% with tertiary education) and 59% in Italy (versus 79%) continued to work during COVID-19.

In order to assess whether age was a factor associated with economic impact, respondents were divided into three age groups in the analysis: 18-34 year olds, 35-64 year olds, and over 65 year olds (Fig. 2, Suppl. Table 5). There were no significant differences between age groups regarding loss of earnings (p=0.102) or loss of job (p=0.054). However, the 18-34 year olds appeared to be most affected through reduction of working hours (p=0.005) and closure of workplace (p=0.003). Only 71% of 18-34 year olds and 68% of 65+ year olds continued to work during COVID-19, compared to 78% of 35-64 year olds (p=0.025). Analysing by country, the 65+ year olds reported highest loss of earnings in Malaysia (57%) and Slovenia (39%). This age group was particularly affected in Italy, where 87% of 65+ year olds reported loss of earnings and 42% reported loss of job. In all countries except for Thailand, fewer 65+ year olds continued to work during COVID-19.

Overall, larger households and having children under 18 in the household appeared to be associated with worse economic impacts (Fig. 2, Suppl. Tables 6 and 7). Overall, 67% of respondents whose household included 6 people or more reported loss of earnings (compared to 54% of households with 1-5 people; p=0.013), and 23% reported loss of job (compared to 13%; p=0.009; Suppl. Table 6). Respondents with children reported a higher loss of earnings compared to respondents without children (62% versus 53%; p=0.005), and higher job loss (18% versus 12%; p=0.008; Suppl. Table 7). Analysing by country, respondents living with children appeared to be particularly affected in Thailand and Malaysia.

We also analysed responses according to three types of income: fixed income (e.g. fixed salary, benefits or pension), flexible income (e.g. contract, freelance), and other/no income (Fig. 2; Suppl. Table 8). We did not ask for amount of income. Overall, respondents with fixed income were less affected economically than those with flexible or other/no income. Of the latter only 38% reported loss of earnings, compared to 81% of respondents with flexible income and 69% of respondents with other/no income (p<0.001). Only 8% of people with fixed income had lost their job, compared to 22% with flexible income and 27% with other/no income (p<0.001). At country level, the trends were similar (Suppl. Table 8). Fewer people with flexible or other/no income continued to work in Malaysia (42% with flexible/25% with no/other income, compared to 83% with fixed income; p<0.001), UK (57%/62%, compared to 79%; p<0.001), Italy (51%/15%, compared to 81%; p<0.001) and Slovenia (57%/59%, compared to 84%; p<0.001).

### Social impacts of COVID-19 and public health measures

We asked respondents if they were concerned about the following areas of life if advised no physical contact/not allowed to go out/allowed to go out only for essential needs: caring responsibilities, physical health, recreational pursuits, sports, mental health and wellbeing, living arrangements, infrastructure (e.g. access to transport, internet), social, and religious and spiritual needs/aspects (Suppl. Table 9). Overall, respondents expressed most concern about their social life (64%), their physical health (59%), and their mental health and wellbeing (58%). This trend was largely similar in individual countries, except for Thailand, where caring responsibilities attracted the most concern (62%); Malaysia, where 58% were concerned about religion and spirituality; and Slovenia, where 65% of people worried about recreational aspects. In general, there were no major differences between gender, age groups, education level, household size, living with children or income type (Suppl. Tables 10-15). Overall, those who were most worried about caring responsibilities were women (52%, versus 42% men, p<0.001; Suppl. Table 10), 35-64 year olds (53%, versus 46% of 18-34 year olds and 32% of 65+ year olds, p<0.001; Suppl. Table 11), people with primary/secondary education (49%, versus 43% with tertiary education, p=0.002; Suppl. Table 12), and people with children (64%, versus 38% of those without children, p<0.001; Suppl. Table 14).

We asked respondents how many days they could cope with not going out except for essential needs/work, with answer options ranging from one to 59 days or more. In total, 44% of respondents said that they could cope for 29 days or longer (Suppl. Table 16). However, coping time varied significantly between countries (p<0.001): in the UK, 60% of people felt they would be able to cope for 29 days or longer, whereas in Thailand, only 26% of respondents said that they could cope this long. Overall, gender, age, and household size did not appear to be associated with coping time (Suppl. Tables 17-19). Factors that appeared to be associated with lower coping times were living with children under 18 years (p=0.004, Suppl. Table 20), having primary/secondary education (p<0.001, Suppl. Table 21), and receiving flexible income (p<0.001; Suppl. Table 22). Indicators varied at country level.

### Compliance and acceptance of public health measures

Next, we explored which factors were associated with compliance and agreement with public health measures. Of all respondents, 67% reported that they had changed their social behaviour *before* government restrictions were implemented (Fig. 3; Suppl. Table 23). There were significant differences at country level (p<0.001): 93% of Thai respondents reported voluntary pre-restriction behaviour change, followed by the UK (68%) and Malaysia (64%). Slovenian (47%) and Italian respondents (47%) reported the lowest levels of voluntary pre-restriction behaviour change. Overall, 92% of respondents had used sanitizer products and alcohol, 82% avoided physical contact with anyone, and 79% avoided physical contact with only vulnerable groups. In Thailand and Malaysia, 96% and 95% of respondents indicated that they had been using personal protective equipment (PPE; e.g. face masks and gloves), compared to only 33% in UK, 55% in Italy, and 67% in Slovenia (p<0.001). We also asked respondents how much they agreed with quarantine/isolation/social distancing measures and the statement that these are a necessary strategy to help control COVID-19 (Suppl. Table 23). There was a significant difference between countries (p<0.001): agreement with public health measures was highest amongst respondents from Thailand (94%) and lowest amongst those from Slovenia (around 75%).

**Figure 3:**
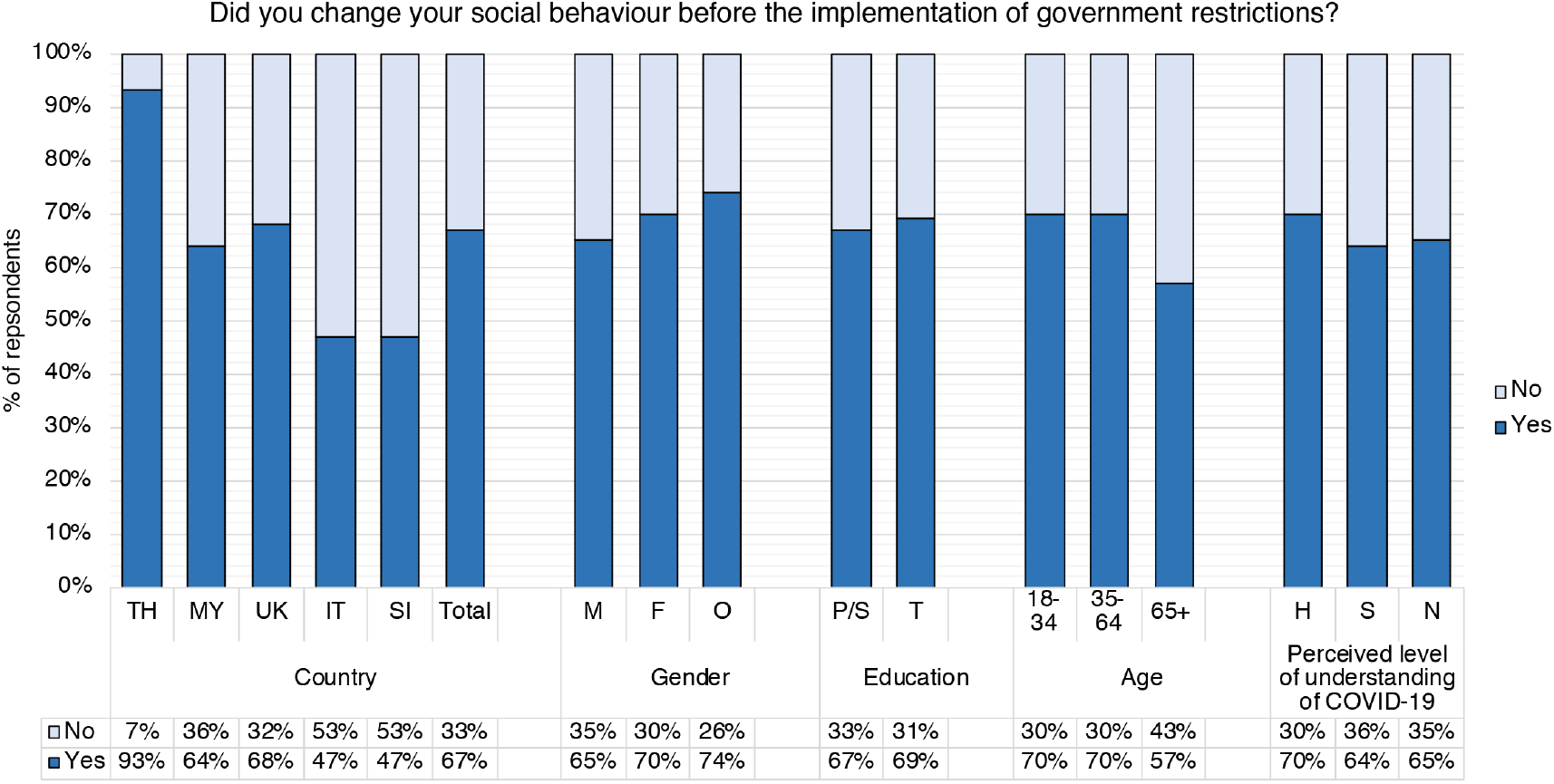
Breakdown of responses to the question “Did you change your social behaviour before the implementation of government restrictions?” by country (TH = Thailand, MY = Malaysia, UK = United Kingdom, IT = Italy, SI = Slovenia) and demographic groups: gender (M = male, F = female, O = other/prefer not to say); education level (P/S = primary or lower/secondary, T = tertiary); age (18-34 years old, 35-64 years old, 65+ years old); self-reported/perceived level of understanding of COVID-19 (H = high/very high/expert level, S = some, N = a little/none at all).

Overall, fewer male than female respondents changed their social behaviour before the government implemented official restrictions (65% and 70%, respectively, p=0.039; Fig. 3, Suppl. Table 24). At country level, fewer men than women reported changing their social behaviour voluntarily, except in Thailand, where reported changes among men and women were similar (94%/92%, p=0.426). Overall, there were no significant differences between men and women when asked about how much they agreed with public health measures and the statement that these are a necessary strategy to help control COVID-19 (p=0.191; Suppl. Table 24).

When it came to education level, there were no significant differences between respondents with primary/secondary and those with tertiary education regarding voluntary behaviour change before government-imposed restrictions (p=0.369), and agreement with public health measures and the statement that these are a necessary strategy to help control COVID-19 (p=0.304; Fig. 3, Suppl. Table 25). Indicators varied at country level.

Overall, 70% of 18-34 year olds and 70% of 35-64 year olds indicated that they had voluntarily changed their behaviour before government restrictions, compared to only 57% of 65+ year olds (p=0.004; Fig. 3, Suppl. Table 26). This trend was similar at country level, except in Italy where 57% of 65+ year olds were most likely to change their behaviour, compared with 44% of 18-34 and 44% of 35-64 year olds. Overall, agreement with voluntary restrictions was similar across age groups (p=0.271; Suppl. Table 26), but fewer 65+ year expressed agreement with restrictions that were government-enforced (p=0.003). Respondents over 65 years old in Slovenia reported the lowest agreement with the statement that quarantine/isolation/social distancing are a necessary strategy to help control COVID-19 (67%), compared to 96% in Thailand and 100% in Malaysia.

Lastly, self-reported levels of understanding of COVID-19 did not significantly affect voluntary change of behaviour (p=0.091), or agreement with public health measures (p=0.688; Suppl. Table 27).

### Level of understanding of COVID-19

We asked respondents to indicate their perceived level of understanding of COVID-19. Overall, 59% of respondents indicated a ‘high/very high’ level of understanding, 36% reported ‘some’ understanding, and only 5% reported ‘very little/none’ (Fig. 4, Suppl. Table 28). There were significant differences at country level (p<0.001): perceived levels of understanding were highest in Slovenia (66% reported ‘high/very high’, and 30% ‘some’ understanding) and Thailand (63% ‘high/very high’ and 33% ‘some’), and lowest in Italy, with 47% reporting ‘high/very high’, and 50% reporting ‘some’ level of understanding.

**Figure 4:**
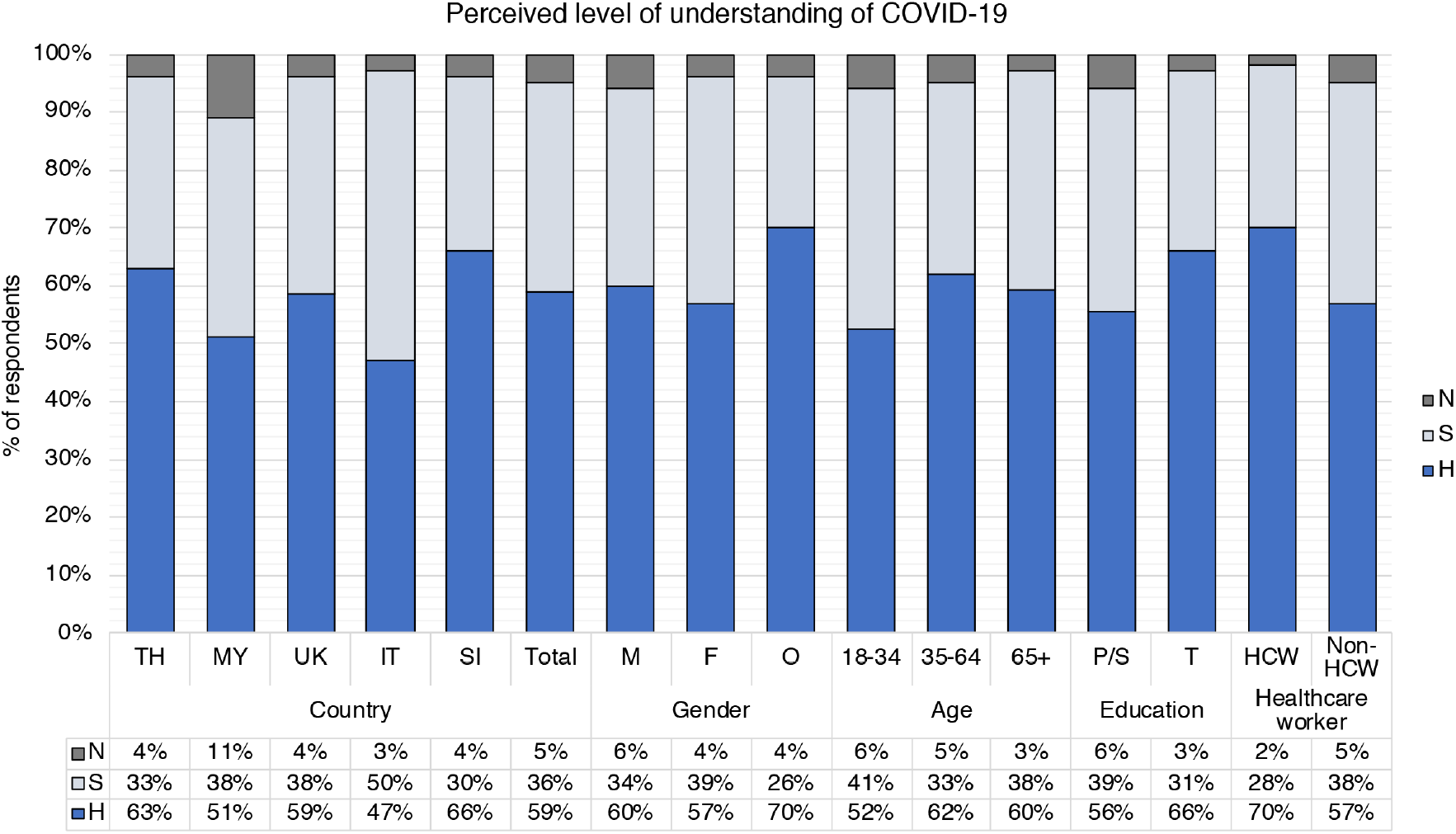
Breakdown of responses to the question “How would you rate your level understanding of the current quarantine/isolation/social distancing requirements for COVID-19?” Self-reported/perceived level of understanding of COVID-19 ((H = high/very high/expert level, S = some, N = a little/none at all) shown by country (TH = Thailand, MY = Malaysia, UK = United Kingdom, IT = Italy, SI = Slovenia) and demographic groups: gender (M = male, F = female, O = other/prefer not to say); age (18-34 years old, 35-64 years old, 65+ years old); education level (P/S = primary/secondary, T = tertiary); healthcare worker status (HCW = healthcare worker, Non-HCW = non-healthcare worker).

To probe for factors associated with perceived level of understanding of COVID-19, we broke down responses by gender, age, education and healthcare worker status (Fig. 4, Suppl. Table 29). Overall, there was no significant difference between men, women and people who identified as other or preferred not to say (p=0.058; Fig. 4, Suppl. Table 29). Age appeared to be a factor, as only 52% of 18-34 year old respondents self-reported ‘high/very high’ understanding compared to 62% of 35-64 year olds and 60% of 65+ year olds (p=0.033). Overall, fewer respondents with primary and secondary education self-reported ‘high/very high’ understanding (56% indicated ‘high/very high’ compared to 66% with tertiary education, p<0.001). Lastly, healthcare worker status was associated with perceived higher understanding (p=0.001). This trend was similar at country level, except for Malaysia, where 49% of healthcare workers reported ‘high/very high’ understanding compared to 52% of non-healthcare workers (p=0.805) (Suppl. Table 29).

Overall, higher levels of perceived understanding of COVID-19 were associated with higher levels of perceived understanding of public health measures (p<0.001; Suppl. Table 30). For example, 88% of respondents who self-reported ‘high/very high’ understanding of COVID-19 and 50% who reported ‘some’ understanding felt that they had a ‘high/very high’ level of understanding of public health measures. In contrast, only 27% of respondents who reported ‘very little/no’ understanding of COVID-19 indicated a high understanding of public health measures.

### Information about COVID-19, unclear information and fake news

When respondents were asked how they receive/received information about COVID-19 (Suppl. Table 31), most reported traditional mass media (TV, radio, newspapers; 93%), followed by online methods (websites, email; 83%) and social media and messenger apps (79%). When asked about their preferences for receiving information, the top three responses were traditional mass media (78%), government or institution’s website (77%), and online (76%). There were no significant differences based on gender (Suppl. Table 32). Fewer respondents over 65 years said that they had used online channels or social media and messenger apps, and they expressed significantly lower preference for these channels too. For example, only 66% of over 65 year olds wanted to receive information online, compared to 78%/79% of the other age groups (p<0.001), and only 52% of over 65 year olds expressed preference for social media and messenger apps, compared to 64%/64% (p=0.005; Suppl. Table 33). Overall, most respondents with primary/secondary education and those with tertiary education had received information through traditional mass media, and social media/messenger apps (Suppl. Table 34). Fewer respondents with primary/secondary education had used online channels in the form of websites and emails (79% versus 92%, p<0.001), and more had received face-to-face information compared to those with tertiary education (43% versus 35%, p<0.001; Suppl. Table 34). However, both education level groups indicated that their preferred methods of communication were mass media channels, online methods and government/institutions’ websites.

We asked respondents if they had seen unclear or conflicting information about COVID-19 in nine categories relating to infection, symptoms and various public health measures. Overall, between 36-54% of respondents indicated that they had seen such information. Ways to avoid the infection (54%), government support schemes (52%) and testing (51%) were identified as the most unclear areas (Suppl. Table 35). Thailand reported the lowest levels of seeing unclear or conflicting information in most categories (around 35-40%), while respondents in the UK reported the highest levels in most categories (around 55-70%). Overall, those with tertiary education reported significantly higher levels of seeing unclear information than those with primary/secondary education in almost all categories (p<0.001) except government support schemes (Suppl. Table 36).

When asked “Have you come across news about the following COVID-19 topics that seemed fake to you?”, overall 63% of respondents had encountered news on “Coronavirus as an engineered modified virus”, 60% reported seeing “general spread of fear”, and 51% had come across seemingly fake news about “numbers of infected/deceased people”, “home-made recipes to make sanitizer products” and “alternative drugs/cure” (Fig. 5, Suppl. Table 35). Thailand reported the lowest percentages in all ‘fake news’ categories, with a range of 27-42% (Suppl. Table 35). Overall, respondents with tertiary education reported significantly higher levels of seeing ‘fake news’ in all categories compared to those with primary/secondary education (p<0.001; Fig. 5, Suppl. Table 36). For example, only 56% of people with primary/secondary education reported coming across fake news about “coronavirus as an engineered modified virus” versus 79% of those with tertiary education (p<0.001). There did not appear to be an association between self-reported levels of understanding of COVID-19 and seeing unclear/conflicting information or ‘fake news’ (Suppl. Table 37).

**Figure 5:**
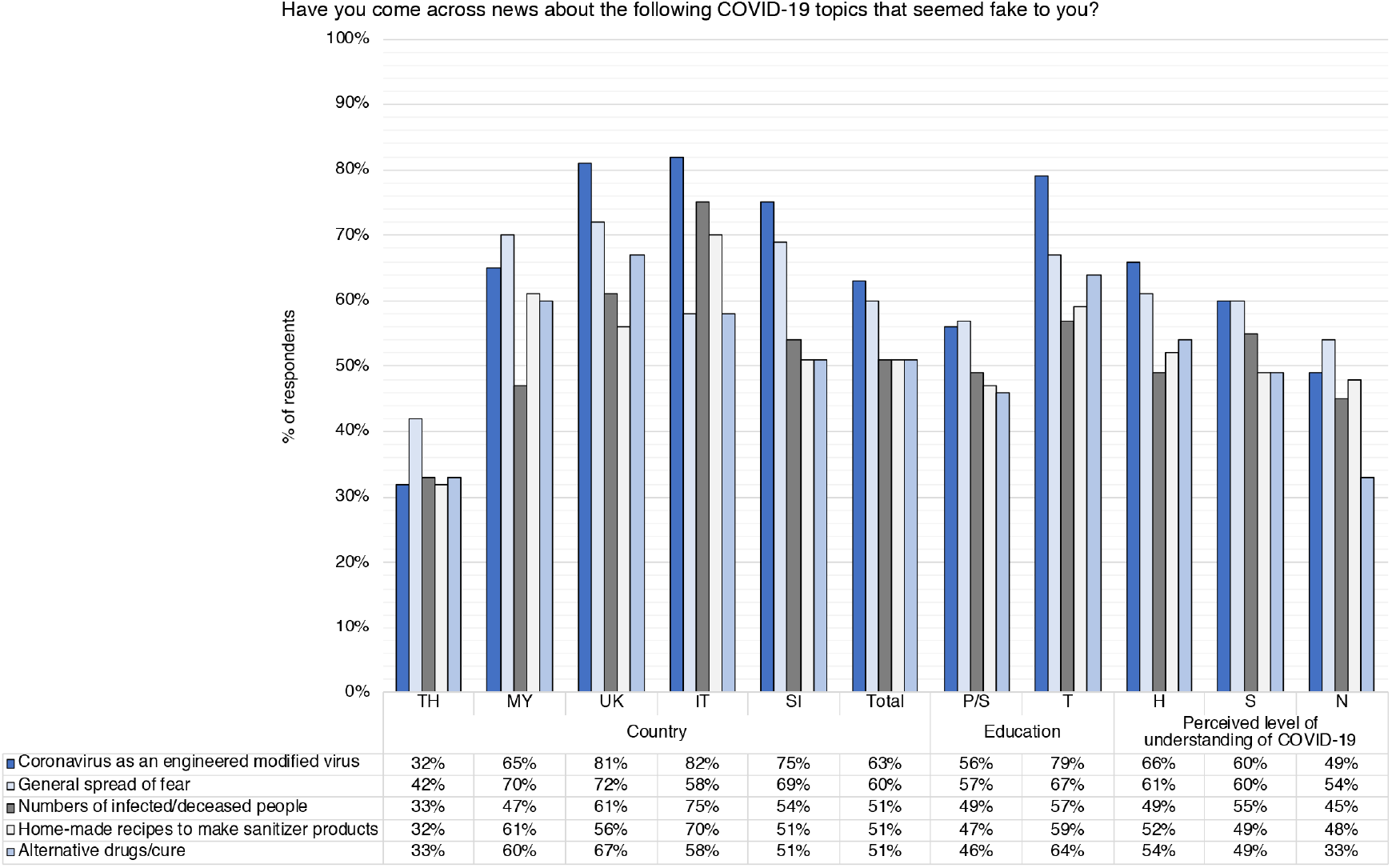
Diagram showing how many survey respondents had come across five ‘fake news’ categories, in response to the question “Have you come across news about the following COVID-19 topics that seemed fake to you?”. Breakdown by country (TH = Thailand, MY = Malaysia, UK = United Kingdom, IT = Italy, SI = Slovenia), gender (M = male, F = female, O = other/prefer not to say), age (18-34 years old, 35-64 years old, 65+ years old), education level (P/S = primary or lower/secondary, T = tertiary), and perceived level of understanding of COVID-19 (H = high/very high/expert level, S = some, N = a little/none at all).

## Discussion

Our results indicate how public health measures that were in place between 1^st^ May and 30^th^ June 2020 affected a cohort of over 5,000 respondents across five countries, and thus contribute new data and insights to these research areas.

### Who was most affected by COVID-19 public health measures?

Overall, lower education levels, larger households, having children under 18 in the household, being 65 years or older, and having flexible/no income were associated with worse economic impact. This confirms that COVID-19 public health measures have greater negative impacts on already disadvantaged groups. Overall, it appeared that the 35-64 year old age group was less affected than 18-34 year olds and people older than 65 years. Possible explanations for this could be the types of sectors that younger and older people work in (e.g low paid or service industries)^32,33^, or for older workers, shielding guidance issued by governments, lower levels of digital skills for remote working^34^, or discrimination in the form of ageism^32,35^. There were no significant differences between gender groups in our overall analysis. However, other studies have shown that COVID-19 has had a greater impact on women (e.g. women are more likely to have temporary contracts^36,37^ and disproportionally carry the burden of unpaid care^38,39^). A more detailed gender analysis to further break down our survey results is currently underway.

Our results showed that among the countries surveyed, respondents from Thailand were most affected. Thailand is a middle-income country with a large informal economy, and relies heavily on the tourism industry (15% GDP)^40^. Thailand also had a high government stringency index during the period of the study (Fig. 1), which included closure of borders, businesses and nighttime curfews^41^. This meant that many informal street vendors and those working in the tourism industry (e.g. tour operators) had no income or lost their jobs.

Overall, about two thirds of respondents were most concerned about the effects of public health measures on their social life, their physical health, and their mental health and wellbeing. These findings resonate with other studies showing the substantial negative impact of COVID-19 restrictions on mental health, wellbeing and social life^42-44^.

### Self-reported compliance and behavioural changes

A number of quantitative online surveys have examined experiences, knowledge, attitude and perceptions towards COVID-19 and public health measures, at country level^36,45-54^, and among different social groups^55-58^. Our findings show that self-reported compliance and behavioural change seemed to differ between countries. For example, respondents in Thailand indicated significantly higher levels of compliance, acceptance of public health measures and voluntary behavioural change compared to other countries. Although our survey was unable to implicate causality, it may contribute to better understanding of why Thailand has the lowest number of COVID cases relative to its population among the countries who took part in the survey^59^. Some of our results with regards to gender and age were similar to trends reported in other studies. For example, results from a Hong Kong study showed that female respondents, and those who reported higher levels of understanding of COVID-19, were more likely to adopt social distancing measures^60^. Similarly, a Chinese study found that men and those with a lower COVID-19 knowledge score were less likely to avoid crowded places or wear a mask outside. Using survey data from 27 countries, Daoust^55^ observed that compliance was not higher in older people even though they might be expected to comply more due to being a risk group. Similarly, our data showed that overall and in Malaysia, UK and Slovenia, far fewer respondents over 65 years reported changing their behaviour voluntarily before official restrictions came into place. However, overall, over 80% of respondents in all three age groups expressed agreement when asked if they would comply voluntarily or with government-mandated restrictions (Suppl. Table 26).

### Improving COVID-19 communication

Our findings indicated that younger age and lower education levels appeared to be associated with lower self-perceived/subjective levels of understanding of COVID-19. Also, higher self-reported levels of understanding of COVID-19 seemed to be associated with higher levels of understanding of public health measures. A recent modelling study suggests that self-imposed public health measures combined with fast spreading of disease awareness in the population can help reduce transmission of the virus^11^. Our findings suggest that specific groups of people, such as those with primary/secondary education levels and those 18-34 year old, may benefit most from targeted COVID-19 communication initiatives.

In terms of channels of communications, the three most popular channels across countries were traditional mass media, government or institutional websites, and online media. Similar results emerged from a recent survey carried out in the Netherlands, Germany and Italy^52^. However, respondents in Thailand reported that they preferred to receive information face-to-face, especially those with primary/secondary education. This suggests that in order for communication strategies to be effective, they need to be sensitive to population preferences and tailored to local contextual factors (e.g. levels of connectivity, literacy^61^).

Our survey showed that a significant proportion of the population received conflicting information and news that seemed fake to them, in particular about coronavirus being an engineered modified virus. These findings confirm other reports that misinformation and what has been termed the COVID-19 ‘infodemic’ is widespread^56,62,63^. More efforts should be made to curb misinformation and disinformation, taking into account the needs of different groups^44^.

### Strengths and limitations

Our online survey enabled us to capture people’s experiences and concerns in multiple domains, in five countries, all of which had restrictions in place, during the relatively early stage of the COVID-19 pandemic. To our knowledge, the SEBCOV study was one of the largest international mixed-methods studies conducted on the impact of COVID-19. To maximise the number of respondents and the likelihood of getting honest answers, the survey was completely anonymous. Due to the relatively large sample of respondents in each country, we were able to compare population segments (e.g. men versus women or younger versus older people) in our overall cohort and at country level. We did not aim to obtain nationally representative samples and acknowledge that although we used weighting strategies in our analysis, our results may not be fully representative of the populations in the respective countries. Overall, there was a high proportion of respondents who were healthcare workers (19%), and some variation in this proportion between countries. This may have influenced the country level analysis, in particular in the areas of perceived understanding, compliance/agreement and communication preferences.

Because the survey was online, only people who were literate, had internet access, and had access to computers or smartphones could take part. Due to COVID-19 related restrictions, it was not possible to conduct face-to-face data collection to reach groups who were illiterate in the language of the survey, or who did not have access to online technology. This is likely to have biased our data towards more educated and economically advantaged populations. Our study was also subject to response bias and other biases arising from self-reporting and recall. Lastly, as a cross-sectional survey, our data only sheds light on the prevalence of certain phenomena and opinions of respondents but does not imply causality.

The results of the survey reported here form part of a mixed-methods study, which also includes an in-depth qualitative study, the findings of which are currently being analysed and will be published separately. Combined, our results may help explain some of the trends reported in this survey, as well as the differences between countries and social groups. We have also conducted a preliminary analysis of unweighted Thai survey responses during May 2020, which includes more detailed breakdowns by regions within Thailand^64^.

## Conclusion

NPIs such as lockdowns and social distancing measures to mitigate transmission of COVID-19 exert substantial negative economic and social impacts^44^. Our data confirmed that NPIs have unequal effects on different countries and different social groups within countries, and contributes to an important body of research showing that lockdowns most negatively affect those who are socio-economically disadvantaged^50,53^. As such, this study helps to expose some of the social and economic inequalities resulting from COVID-19 and public health measures. Our findings provide an indication of the social groups who may be most in need of support during pandemics, so that existing social inequalities are not perpetuated and worsened. Lastly, in order to mitigate the impacts of COVID-19, we need effective communication^19^, and our data can help to inform future strategies.

## Data Availability

The Mahidol Oxford Tropical Medicine Research Unit recognizes the value of sharing individual level data. We aim to ensure that data generated from all our research are collected, curated, managed and shared in a way that maximizes their benefit. Data underlying this publication are available upon request to the Mahidol Oxford Tropical Medicine Research Uni Data Access Committee at https://www.tropmedres.ac/units/moru-bangkok/bioethics-engagement/data-sharing.

https://zenodo.org/record/4049821

https://doi.org/10.12688/wellcomeopenres.15813.2

## Ethics approval

Ethics approval was granted by Oxford Tropical Research Ethics Committee (OxTREC, reference no.520-20), covering all countries; the Faculty of Tropical Medicine Ethics Committee, Thailand (FTMEC, ref: MUTM 2020-031-01); the Medical Research and Ethics Committee (MREC), Ministry of Health Malaysia (MOH), Malaysia, ref: NMRR-20-595-54437 (IIR), and the Universiti Tunku Abdul Rahman (Utar) Scientific and Ethical Review Committee (SERC, ref: (U/SERC/63/2020), Malaysia; and the National Medical Ethics Committee of the Republic of Slovenia (0120-237/2020/7). Additional ethics committee approval from Italy was not required for the study to be conducted there.

## Acknowledgements

We would like to thank all participants who took part in our study. We also thank the people who helped to test and promote the survey, and the members of the Bangkok Health Research and Ethics Group for their input into the study. We would like to thank the SoNAR-Global Network for the social science expertise in the development of this project and the ODK core team for their input in design of the quantitative survey. Lastly, we would like to thank Prof. Nicholas PJ Day, Prof Richard Maude and Dr Tamara Giles-Vernick for critical reading of the manuscript.

## Contributorship statement

AO and PYC oversaw the whole project and wrote the initial draft of the manuscript. AO, GC, WP, PKC, PC, MS, MLS, TS, NW, SA, BN, SR, NK, DO, RC and PYC developed the survey and translations. AO, GC, WP, PC, LS led the project in the UK, Italy, Thailand, Malaysia and Slovenia, respectively. MM and PP conducted the statistical analysis, figures and tables, with critical input from MS, AO and PYC. MLS critically reviewed the manuscript, figures and tables. All authors implemented the survey, contributed to the draft paper, and approved the final version of the paper. PYC conceived the project and is the guarantor of the paper.

## Transparency declaration

The corresponding author (manuscript guarantor) affirms that this manuscript is an honest, accurate, and transparent account of the study being reported; that no important aspects of the study have been omitted; and that any discrepancies from the study as planned (and, if relevant, registered) have been explained.

