## Supplementary Tables 1-37 for "Economic and social impacts of COVID-19 and public health measures: results from an anonymous online survey in Thailand, Malaysia, the United Kingdom, Italy and Slovenia"

**Notes**

- There are a total of 37 tables in this document. Suppl. Table 1 reports the distribution of the basic demographic variables in the respondent sample (N= number of respondents), followed by unweighted percentages (unweighted %) in brackets. The values displayed in the cells in Suppl. Tables 2-37 show the number of respondents (N) who replied ‘yes’ to the respective survey categories, followed by weighted percentages (weighted %) in brackets.
- Because of rounding to the nearest integer, percentages do not always add up to 100% exactly.
- For gender, due to small number in the “other/prefer not to say“ category, p-values are presented for comparison between the male and female groups only.

**Contents**

**Supplementary tables for “Economic and social impacts of COVID-19 and public health measures: results from an anonymous online survey in Thailand, Malaysia, the United Kingdom, Italy and Slovenia”**	1

Suppl. Table 1 Distribution of respondents by demographic characteristics and country (unweighted data)	4

Suppl. Table 2 Breakdown of economic impacts of COVID-19 and concerns by country	5

Suppl. Table 3 Breakdown of economic impacts of COVID-19 and concerns by country and gender	6

Suppl. Table 4 Breakdown of economic impacts of COVID-19 and concerns by country and education level	7

Suppl. Table 5 Breakdown of economic impacts of COVID-19 and concerns by country and age group	8

Suppl. Table 6 Breakdown of economic impacts of COVID-19 and concerns by country and household size	9

Suppl. Table 7 Breakdown of economic impacts of COVID-19 and concerns by country and whether or not living with children under 18	10

Suppl. Table 8 Breakdown of economic impacts of COVID-19 and concerns by country and type of income	11

Suppl. Table 9 Breakdown of concerns if advised/not allowed physical contact by country	12

Suppl. Table 10 Breakdown of concerns if advised/not allowed physical contact by country and gender	13

Suppl. Table 11 Breakdown of concerns if advised/not allowed physical contact by country and age group	14

Suppl. Table 12 Breakdown of concerns if advised/not allowed physical contact by country and education level	15

Suppl. Table 13 Breakdown of concerns if advised/not allowed physical contact by country and household size	16

Suppl. Table 14 Breakdown of concerns if advised/not allowed physical contact by country and whether or not living with children under 18	17

Suppl. Table 15 Breakdown of concerns if advised/not allowed physical contact by country and income type	18

Suppl. Table 16 Breakdown of maximum number of days that people thought they could cope by country	19

Suppl. Table 17 Breakdown of maximum number of days that people thought they could cope by country and gender	20

Suppl. Table 18 Breakdown of maximum number of days that people thought they could cope by country and age group	21

Suppl. Table 19 Breakdown of maximum number of days that people thought they could cope by country and household size	22

Suppl. Table 20 Breakdown of maximum number of days that people thought they could cope by country and whether or not living with children under 18	23

Suppl. Table 21 Breakdown of maximum number of days that people thought they could cope by country and education level	24

Suppl. Table 22 Breakdown of maximum number of days that people thought they could cope by country and type of income	25

Suppl. Table 23 Breakdown of behavioural changes and acceptance of government public health measures by country	26

Suppl. Table 24 Breakdown of behavioural changes and acceptance of government public health measures by country and gender	27

Suppl. Table 25 Breakdown of behavioural changes and acceptance of government public health measures by country and education level	29

Suppl. Table 26 Breakdown of behavioural changes and acceptance of government public health measures by age group	30

Suppl. Table 27 Breakdown of behavioural changes and acceptance of government public health measures by self-reported level of understanding of COVID-19	32

Suppl. Table 28 Breakdown of self-reported level of understanding of COVID-19 by country	34

Suppl. Table 29 Breakdown of self-reported level of understanding of COVID-19 by demographic characteristics	35

Suppl. Table 30 Breakdown of self-reported understanding of public health measures by self-reported level of understanding of COVID-19	36

Suppl. Table 31 What were the three most common ways people received communication on COVID-19, and what are the three most preferred ways to receive COVID-19 communications? Breakdown by country	37

Suppl. Table 32 What were the three most common ways people received communications on COVID-19, and what are the three most preferred ways to receive COVID-19 communications? Breakdown by country and gender	38

Suppl. Table 33 What were the three most common ways people received communications on COVID-19, and what are the three most preferred ways to receive COVID-19 communications? Breakdown by country and age group	39

Suppl. Table 34 What were the three most common ways people received communications on COVID-19, and what are the three most preferred ways to receive COVID-19 communications? Breakdown by country and education level	40

Suppl. Table 35 Most prevalent topic areas with unclear or conflicting COVID-19 information, and most prevalent ‘fake news’, breakdown by country	41

Suppl. Table 36 Most prevalent topic areas with unclear or conflicting COVID-19 information, and most prevalent ‘fake news’, breakdown by country and education level	42

Suppl. Table 37 Most prevalent topic areas with unclear or conflicting COVID-19 information, and most prevalent ‘fake news’, breakdown by country and self-reported level of understanding of COVID-19	43

### Suppl. Table 1 Distribution of respondents by demographic characteristics and country (unweighted data)

| Variable and categories | Thailand  (N=1,476) | Malaysia  (N=827) | UK  (N=1,009) | Italy  (N=712) | Slovenia  (N=1,034) | Total  (N=5,058) |
| --- | --- | --- | --- | --- | --- | --- |
| Gender |  |  |  |  |  |  |
| Male | 704 (48) | 298 (36) | 426 (42) | 222 (31) | 366 (35) | 2,016 (40) |
| Female | 766 (52) | 525 (63) | 572 (57) | 490 (69) | 662 (64) | 3,015 (60) |
| Other/prefer not to say | 6 (0) | 4 (0) | 11 (1) | 0 (0) | 6 (1) | 27 (1) |
| Age (years) |  |  |  |  |  |  |
| 18-34 | 223 (15) | 350 (42) | 140 (14) | 272 (38) | 308 (30) | 1,293 (26) |
| 35-64 | 1,152 (78) | 442 (53) | 616 (61) | 383 (54) | 676 (65) | 3,269 (65) |
| 65+ | 101 (7) | 35 (4) | 253 (25) | 57 (8) | 50 (5) | 496 (10) |
| Education level |  |  |  |  |  |  |
| Primary or lower/ secondary | 909 (62) | 82 (10) | 247 (24) | 217 (30) | 202 (20) | 1,657 (33) |
| Tertiary | 567 (38) | 745 (90) | 762 (76) | 495 (70) | 832 (80) | 3,401 (67) |
| Household structure |  |  |  |  |  |  |
| Living alone | 134 (9) | 74 (9) | 206 (20) | 106 (15) | 97 (9) | 617 (12) |
| Living only with partner/spouse | 173 (12) | 95 (11) | 391 (39) | 192 (27) | 210 (20) | 1,061 (21) |
| Living with partner/spouse and children; living as single parent with children | 847 (57) | 312 (38) | 260 (26) | 188 (26) | 518 (50) | 2,125 (42) |
| Living with other relatives/non-relatives/other | 322 (22) | 346 (42) | 152 (15) | 226 (32) | 209 (20) | 1,255 (25) |
| Household size |  |  |  |  |  |  |
| 1 | 107 (7) | 68 (8) | 222 (22) | 106 (15) | 128 (12) | 631 (12) |
| 2 | 171 (12) | 121 (15) | 439 (44) | 230 (32) | 220 (21) | 1,181 (23) |
| 3-5 | 995 (67) | 457 (55) | 333 (33) | 360 (51) | 605 (59) | 2,750 (54) |
| ≥6 | 203 (14) | 181 (22) | 15 (1) | 16 (2) | 81 (8) | 496 (10) |
| Type of income |  |  |  |  |  |  |
| Fixed salary/benefits/pension | 546 (37) | 524 (63) | 705 (70) | 347 (49) | 847 (82) | 2,969 (59) |
| Contract and freelance | 849 (58) | 158 (19) | 227 (22) | 244 (34) | 103 (10) | 1,581 (31) |
| Other/no income | 81 (5) | 145 (18) | 77 (8) | 121 (17) | 84 (8) | 508 (10) |
| Living with children under 18 | 664 (45) | 346 (42) | 186 (18) | 144 (20) | 497 (48) | 1,837 (36) |
| Living with vulnerable group* | 457 (31) | 230 (28) | 367 (36) | 151 (21) | 280 (27) | 1,485 (29) |
| Healthcare provider/worker** | 239 (16) | 213 (26) | 118 (12) | 64 (9) | 341 (33) | 975 (19) |

Values in cells are n (%)

* Persons aged 70 or older; pregnant woman; people with serious health conditions

** Included respondents who were not working before COVID-19

### Suppl. Table 2 Breakdown of economic impacts of COVID-19 and concerns by country

Values in cells are n (weighted %) of respondents who replied ‘yes’.

| Variable and categories | Thailand | Malaysia | UK | Italy | Slovenia | Total | P-value |
| --- | --- | --- | --- | --- | --- | --- | --- |
| If you were working before COVID-19, has COVID-19 created any inconvenience for you? | N=1,255 | N=613 | N=630 | N=526 | N=929 | N=3,953 |  |
| Loss of earnings | (N=1,248) 1,012 (85) | (N=556) 155 (40) | (N=584) 226 (44) | (N=496) 260 (55) | (N=867) 219 (30) | (N=3,751) 1,872 (56) | <0.001 |
| Loss of job | (N=1,191) 233 (23) | (N=532) 44 (16) | (N=551) 51 (10) | (N=471) 59 (13) | (N=832) 15 (3) | (N=3,577) 402 (14) | <0.001 |
| Reduction of  working hours | (N=1,210) 492 (42) | (N=546) 228 (52) | (N=570) 201 (39) | (N=484) 233 (48) | (N=862) 319 (41) | (N=3,672) 1,473 (44) | 0.107 |
| Closure of workplace | (N=1,207) 425 (36) | (N=562) 289 (53) | (N=591) 296 (51) | (N=484) 167 (39) | (N=833) 63 (8) | (N=3,677) 1,240 (36) | <0.001 |
| Did you continue to work during COVID-19? | (N=1,255) 1,019 (79) | (N=613) 532 (70) | (N=630) 460 (70) | (N=526) 388 (67) | (N=929) 768 (79) | (N=3,953) 3,167 (75) | 0.011 |
| What are/were your concerns if advised no physical contact/not allowed to go out/allowed to go out only for essential needs? | N=1,476 | N=827 | N=1,009 | N=712 | N=1,034 | N=5,058 |  |
| Financial (e.g. loss of income, loss of job) | (N=1,466) 1,215 (86) | (N=775) 419 (60) | (N=950) 271 (32) | (N=678) 315 (41) | (N=1,015) 302 (28) | (N=4,884) 2,522 (53) | <0.001 |
| Professional/ career progression | (N=1,414) 607 (42) | (N=759) 418 (52) | (N=942) 198 (24) | (N=670) 224 (22) | (N=1,001) 219 (17) | (N=4,786) 1,666 (32) | <0.001 |

### Suppl. Table 3 Breakdown of economic impacts of COVID-19 and concerns by country and gender

M = male; F = female; O = other/prefer not to say. Values in cells are n (weighted %) of respondents who replied ‘yes’.

| Variable and categories | Thailand | | | Malaysia | | | UK | | | Italy | | | Slovenia | | | Total | | | |
| --- | --- | --- | --- | --- | --- | --- | --- | --- | --- | --- | --- | --- | --- | --- | --- | --- | --- | --- | --- |
| Gender | **M** | **F** | **O** | **M** | **F** | **O** | **M** | **F** | **O** | **M** | **F** | **O** | **M** | **F** | **O** | **M** | **F** | **O** | **P-value (for total**  **M vs F)** |
| If you were working before COVID-19, has COVID-19 created any inconvenience for you? | N=606 | N=645 | N=4 | N=230 | N=380 | N=3 | N=261 | N=363 | N=6 | N=184 | N=342 | N=0 | N=332 | N=591 | N=6 | N=1,613 | N=2,321 | N=19 |  |
| Loss of earnings | (N=604) 508 (83) | (N=640) 502 (86) | (N=4) 2 (50) | (N=210) 75 (42) | (N=343) 80 (37) | (N=3) 0 (0) | (N=245) 97 (45) | (N=333) 128 (43) | (N=6) 1 (17) | (N=177) 99 (54) | (N=319) 161 (57) |  | (N=314) 82 (29) | (N=548) 135 (31) | (N=5) 2 (40) | (N=1,550) 861 (55) | (N=2,183) 1,006 (57) | (N=18) 5 (28) | 0.531 |
| Loss of job | (N=576) 104 (20) | (N=611) 129 (25) | (N=4) 0 (0) | (N=202) 17 (18) | (N=327) 27 (15) | (N=3) 0 (0) | (N=233) 21 (19) | (N=313) 30 (11) | (N=5) 0 (0) | (N=168) 19 (10) | (N=303) 40 (17) |  | (N=301) 3 (1) | (N=526) 12 (4) | (N=5) 0 (0) | (N=1,480) 164 (13) | (N=2,080) 238 (16) | (N=17) 0 (0) | 0.157 |
| Reduction of  working hours | (N=586) 225 (41) | (N=620) 265 (43) | (N=4) 2 (50) | (N=205) 85 (57) | (N=338) 141 (46) | (N=3) 2 (67) | (N=240) 90 (41) | (N=324) 107 (37) | (N=6) 4 (67) | (N=174) 94 (52) | (N=310) 139 (43) |  | (N=315) 128 (44) | (N=541) 188 (39) | (N=6) 3 (50) | (N=1,520) 622 (45) | (N=2,133) 840 (42) | (N=19) 11 (58) | 0.179 |
| Closure of workplace | (N=581) 194 (35) | (N=622) 231 (37) | (N=4) 0 (0) | (N=208) 109 (48) | (N=351) 178 (60) | (N=3) 2 (67) | (N=251) 124 (50) | (N=334) 169 (51) | (N=6) 3 (50) | (N=172) 65 (38) | (N=312) 102 (41) |  | (N=302) 19 (7) | (N=526) 43 (9) | (N=5) 1 (20) | (N=1,514) 511 (35) | (N=2,145) 723 (37) | (N=18) 6 (33) | 0.365 |
| Did you continue to work during COVID-19? | (N=606) 508 (84) | (N=645) 507 (75) | (N=4) 4 (100) | (N=230) 198 (67) | (N=380) 332 (73) | (N=3) 2 (67) | (N=261) 198 (72) | (N=363) 258 (67) | (N=6) 4 (67) | (N=184) 144 (74) | (N=342) 244 (60) |  | (N=332) 295 (85) | (N=591) 469 (74) | (N=6) 4 (67) | (N=1,613) 1,343 (78) | (N=2,321) 1,810 (71) | (N=19) 14 (74) | 0.010 |
| What are/were your concerns if advised no physical contact/not allowed to go out/allowed to go out only for essential needs? | N=704 | N=766 | N=6 | N=298 | N=525 | N=4 | N=261 | N=363 | N=6 | N=222 | N=490 | N=0 | N=366 | N=662 | N=6 | N=2,016 | N=3,015 | N=27 |  |
| Financial | (N=700) 592 (85) | (N=760) 619 (86) | (N=6) 4 (67) | (N=279) 155 (62) | (N=492) 261 (59) | (N=4) 3 (75) | (N=411) 113 (34) | (N=529) 154 (31) | (N=10) 4 (40) | (N=214) 113 (44) | (N=464) 202 (38) |  | (N=361) 110 (27) | (N=648) 188 (29) | (N=6) 4 (67) | (N=1,965) 1,083 (54) | (N=2,893) 1,424 (53) | (N=26) 15 (58) | 0.806 |
| Professional/ career progression | (N=675) 278 (41) | (N=733) 326 (42) | (N=6) 3 (50) | (N=270) 137 (53) | (N=485) 279 (51) | (N=4) 2 (50) | (N=409) 84 (26) | (N=523) 108 (22) | (N=10) 6 (60) | (N=211) 92 (26) | (N=459) 132 (18) |  | (N=354) 77 (14) | (N=641) 141 (19) | (N=6) 1 (17) | (N=1,919) 668 (32) | (N=2,841) 986 (31) | (N=26) 12 (46) | 0.597 |

### Suppl. Table 4 Breakdown of economic impacts of COVID-19 and concerns by country and education level

P/S = primary or lower/secondary education; T = tertiary education. Values in cells are n (weighted %) of respondents who replied ‘yes’.

| Variable and categories | Thailand | | Malaysia | | UK | | Italy | | Slovenia | | Total | | |
| --- | --- | --- | --- | --- | --- | --- | --- | --- | --- | --- | --- | --- | --- |
| Education level | **P/S** | **T** | **P/S** | **T** | **P/S** | **T** | **P/S** | **T** | **P/S** | **T** | **P/S** | **T** | **P-value (for total)** |
| If you were working before COVID-19, has COVID-19 created any inconvenience for you? | N=785 | N=470 | N=53 | N=560 | N=122 | N=508 | N=136 | N=390 | N=160 | N=769 | N=1,256 | N=2,697 |  |
| Loss of earnings | (N=780) 725 (90) | (N=468) 287 (62) | (N=50) 21 (42) | (N=506) 134 (28) | (N=116) 55 (58) | (N=468) 171 (34) | (N=126) 75 (58) | (N=370) 185 (52) | (N=150) 56 (36) | (N=717) 163 (24) | (N=1,222) 932 (65) | (N=2,529) 940 (38) | <0.001 |
| Loss of job | (N=744) 164 (24) | (N=447) 69 (16) | (N=50) 9 (19) | (N=482) 35 (7) | (N=108) 12 (13) | (N=443) 39 (9) | (N=123) 18 (14) | (N=348) 41 (12) | (N=140) 7 (4) | (N=692) 8 (1) | (N=1,165) 210 (17) | (N=2,412) 192 (8) | <0.001 |
| Reduction of  working hours | (N=762) 332 (43) | (N=448) 160 (37) | (N=48) 25 (55) | (N=498) 203 (40) | (N=110) 42 (49) | (N=460) 159 (32) | (N=125) 63 (47) | (N=359) 170 (49) | (N=144) 72 (46) | (N=718) 247 (35) | (N=1,189) 534 (47) | (N=2,483) 939 (37) | <0.001 |
| Closure of workplace | (N=753) 262 (36) | (N=454) 163 (37) | (N=48) 28 (55) | (N=514) 261 (49) | (N=116) 51 (48) | (N=475) 245 (52) | (N=130) 59 (44) | (N=354) 108 (31) | (N=137) 14 (8) | (N=696) 49 (7) | (N=1,184) 414 (37) | (N=2,493) 826 (34) | 0.180 |
| Did you continue to work during COVID-19? | (N=785) 613 (78) | (N=470) 406 (86) | (N=53) 34 (65) | (N=560) 498 (90) | (N=122) 73 (59) | (N=508) 387 (77) | (N=136) 75 (59) | (N=390) 313 (79) | (N=160) 115 (74) | (N=769) 653 (85) | (N=1,256) 910 (71) | (N=2,697) 2,257 (83) | <0.001 |
| What are/were your concerns if advised no physical contact/not allowed to go out/allowed to go out only for essential needs? | N=909 | N=567 | N=82 | N=745 | N=247 | N=762 | N=217 | N=495 | N=202 | N=832 | N=1,657 | N=3,401 |  |
| Financial | (N=904) 828 (89) | (N=562) 387 (68) | (N=75) 46 (62) | (N=700) 373 (55) | (N=232) 64 (34) | (N=718) 207 (31) | (N=205) 96 (39) | (N=473) 219 (46) | (N=193) 71 (29) | (N=822) 231 (27) | (N=1,609) 1,105 (59) | (N=3,275) 1,417 (41) | <0.001 |
| Professional/ career progression | (N=865) 326 (39) | (N=549) 281 (54) | (N=72) 36 (50) | (N=687) 382 (59) | (N=228) 21 (16) | (N=714) 177 (31) | (N=198) 42 (15) | (N=472) 182 (37) | (N=192) 37 (13) | (N=809) 182 (22) | (N=1,555) 462 (30) | (N=3,231) 1,204 (36) | 0.004 |

### Suppl. Table 5 Breakdown of economic impacts of COVID-19 and concerns by country and age group

Values in cells are n (weighted %) of respondents who replied ‘yes’.

| Variable and categories | Thailand | | | Malaysia | | | UK | | | Italy | | | Slovenia | | | Total | | | |
| --- | --- | --- | --- | --- | --- | --- | --- | --- | --- | --- | --- | --- | --- | --- | --- | --- | --- | --- | --- |
| Age group | **18-34** | **35-64** | **65+** | **18-34** | **35-64** | **65+** | **18-34** | **35-64** | **65+** | **18-34** | **35-64** | **65+** | **18-34** | **35-64** | **65+** | **18-34** | **35-64** | **65+** | **P-value (for total)** |
| If you were working before COVID-19, has COVID-19 created any inconvenience for you? | N=155 | N=1,027 | N=73 | N=219 | N=378 | N=16 | N=104 | N=466 | N=60 | N=190 | N=324 | N=12 | N=259 | N=646 | N=24 | N=927 | N=2,841 | N=185 |  |
| Loss of earnings | (N=154) 103 (78) | (N=1,021) 851 (89) | (N=73) 58 (80) | (N=207) 48 (43) | (N=334) 98 (34) | (N=15) 9 (57) | (N=100) 32 (49) | (N=427) 168 (41) | (N=57) 26 (46) | (N=185) 97 (51) | (N=299) 155 (54) | (N=12) 8 (87) | (N=253) 67 (31) | (N=595) 144 (29) | (N=19) 8 (39) | (N=899) 347 (53) | (N=2,676) 1,416 (56) | (N=176) 109 (67) | 0.102 |
| Loss of job | (N=148) 36 (28) | (N=972) 183 (20) | (N=71) 14 (22) | (N=204) 22 (26) | (N=314) 20 (10) | (N=14) 2 (13) | (N=98) 10 (13) | (N=401) 35 (9) | (N=52) 6 (8) | (N=181) 22 (12) | (N=282) 35 (12) | (N=8) 2 (42) | (N=248) 6 (3) | (N=567) 9 (3) | (N=17) 0 (0) | (N=879) 96 (18) | (N=2,536) 282 (12) | (N=162) 24 (17) | 0.054 |
| Reduction of  working hours | (N=147) 73 (53) | (N=991) 401 (42) | (N=72) 18 (23) | (N=206) 85 (57) | (N=325) 136 (49) | (N=15) 7 (50) | (N=100) 31 (43) | (N=416) 145 (36) | (N=54) 25 (45) | (N=182) 87 (50) | (N=292) 143 (50) | (N=10) 3 (16) | (N=249) 99 (47) | (N=593) 212 (39) | (N=20) 8 (38) | (N=884) 375 (50) | (N=2,617) 1,037 (43) | (N=171) 61 (31) | 0.005 |
| Closure of workplace | (N=151) 66 (46) | (N=984) 340 (35) | (N=72) 19 (24) | (N=207) 93 (55) | (N=340) 184 (48) | (N=15) 12 (83) | (N=100) 57 (56) | (N=434) 215 (49) | (N=57) 24 (44) | (N=185) 76 (49) | (N=289) 85 (32) | (N=10) 6 (86) | (N=246) 27 (14) | (N=570) 35 (6) | (N=17) 1 (3) | (N=889) 319 (44) | (N=2,617) 859 (32) | (N=171) 62 (35) | 0.003 |
| Did you continue to work during COVID-19? | (N=155) 120 (77) | (N=1,027) 838 (80) | (N=73) 61 (81) | (N=219) 195 (57) | (N=378) 330 (82) | (N=16) 7 (43) | (N=104) 79 (69) | (N=466) 346 (72) | (N=60) 35 (56) | (N=190) 134 (69) | (N=324) 250 (70) | (N=12) 4 (13) | (N=259) 209 (77) | (N=646) 540 (81) | (N=24) 19 (72) | (N=927) 737 (71) | (N=2,841) 2,304 (78) | (N=185) 126 (68) | 0.025 |
| What are/were your concerns if advised no physical contact/not allowed to go out/allowed to go out only for essential needs? | N=223 | N=1,152 | N=101 | N=350 | N=442 | N=35 | N=140 | N=616 | N=253 | N=272 | N=383 | N=57 | N=308 | N=676 | N=50 | N=1,293 | N=3,269 | N=496 |  |
| Financial | (N=220) 161 (83) | (N=1,145) 985 (89) | (N=101) 69 (78) | (N=338) 198 (60) | (N=408) 211 (64) | (N=29) 10 (42) | (N=134) 59 (48) | (N=581) 195 (35) | (N=235) 17 (6) | (N=270) 138 (50) | (N=356) 168 (48) | (N=52) 9 (20) | (N=305) 92 (31) | (N=664) 205 (36) | (N=46) 5 (4) | (N=1,267) 648 (59) | (N=3,154) 1,764 (58) | (N=463) 110 (30) | <0.001 |
| Professional/ career progression | (N=215) 126 (52) | (N=1,106) 452 (39) | (N=93) 29 (31) | (N=336) 238 (65) | (N=395) 173 (43) | (N=28) 7 (26) | (N=134) 76 (52) | (N=572) 118 (17) | (N=236) 4 (2) | (N=269) 122 (43) | (N=350) 99 (23) | (N=51) 3 (1) | (N=303) 108 (34) | (N=654) 109 (15) | (N=44) 2 (1) | (N=1,257) 670 (51) | (N=3,077) 951 (28) | (N=452) 45 (11) | <0.001 |

### Suppl. Table 6 Breakdown of economic impacts of COVID-19 and concerns by country and household size

Values in cells are n (weighted %) of respondents who replied ‘yes’.

| Variable and categories | Thailand | | Malaysia | | UK | | Italy | | Slovenia | | Total | | |
| --- | --- | --- | --- | --- | --- | --- | --- | --- | --- | --- | --- | --- | --- |
| Household size (number of persons in the household) | **1-5** | **≥6** | **1-5** | **≥6** | **1-5** | **≥6** | **1-5** | **≥6** | **1-5** | **≥6** | **1-5** | **≥6** | **P-value**  **(for total)** |
| If you were working before COVID-19, has COVID-19 created any inconvenience for you? | N=1,079 | N=176 | N=483 | N=130 | N=618 | N=12 | N=518 | N=8 | N=858 | N=71 | N=3,556 | N=397 |  |
| Loss of earnings | (N=1,073) 864 (85) | (N=175) 148 (85) | (N=441) 120 (35) | (N=115) 35 (53) | (N=573) 221 (43) | (N=11) 5 (66) | (N=489) 256 (55) | (N=7) 4 (66) | (N=800) 201 (29) | (N=67) 18 (39) | (N=3,376) 1,662 (54) | (N=375) 210 (67) | 0.013 |
| Loss of job | (N=1,026) 190 (21) | (N=165) 43 (29) | (N=423) 29 (13) | (N=109) 15 (25) | (N=540) 51 (11) | (N=11) 0 (0) | (N=465) 59 (13) | (N=6) 0 (0) | (N=768) 14 (2) | (N=64) 1 (5) | (N=3,222) 343 (13) | (N=355) 59 (23) | 0.009 |
| Reduction of  working hours | (N=1,043) 423 (42) | (N=167) 69 (59) | (N=434) 181 (44) | (N=112) 47 (72) | (N=558) 195 (38) | (N=12) 6 (57) | (N=477) 231 (52) | (N=7) 2 (50) | (N=792) 285 (39) | (N=70) 34 (61) | (N=3,304) 1,315 (42) | (N=368) 158 (55) | 0.009 |
| Closure of workplace | (N=1,039) 364 (36) | (N=168) 61 (34) | (N=443) 223 (47) | (N=119) 66 (72) | (N=579) 292 (51) | (N=12) 4 (25) | (N=476) 162 (39) | (N=8) 5 (72) | (N=768) 58 (8) | (N=65) 5 (7) | (N=3,305) 1,099 (35) | (N=372) 141 (42) | 0.155 |
| Did you continue to work during COVID-19? | (N=1,079) 884 (80) | (N=176) 135 (78) | (N=483) 424 (73) | (N=130) 108 (63) | (N=618) 450 (70) | (N=12) 10 (83) | (N=518) 384 (67) | (N=8) 4 (56) | (N=858) 712 (80) | (N=71) 56 (74) | (N=3,556) 2,854 (75) | (N=397) 313 (72) | 0.564 |
| What are/were your concerns if advised no physical contact/not allowed to go out/allowed to go out only for essential needs? | N=1,273 | N=203 | N=646 | N=181 | N=994 | N=15 | N=696 | N=16 | N=953 | N=81 | N=4,562 | N=496 |  |
| Financial | (N=1,264) 1,050 (87) | (N=202) 165 (80) | (N=602) 317 (60) | (N=173) 102 (63) | (N=935) 266 (33) | (N=15) 5 (24) | (N=662) 306 (41) | (N=16) 9 (49) | (N=935) 282 (27) | (N=80) 20 (37) | (N=4,398) 2,221 (52) | (N=486) 301 (66) | 0.003 |
| Professional/ career progression | (N=1,220) 503 (40) | (N=194) 104 (49) | (N=593) 317 (51) | (N=166) 101 (56) | (N=928) 196 (24) | (N=14) 2 (9) | (N=654) 218 (22) | (N=16) 6 (28) | (N=920) 202 (16) | (N=81) 17 (21) | (N=4,315) 1,436 (30) | (N=471) 230 (46) | <0.001 |

### Suppl. Table 7 Breakdown of economic impacts of COVID-19 and concerns by country and whether or not living with children under 18

Y = living with children under 18; N = not living with children under 18. Values in cells are n (weighted %) of respondents who replied ‘yes’.

| Variable and categories | Thailand | | Malaysia | | UK | | Italy | | Slovenia | | Total | | |
| --- | --- | --- | --- | --- | --- | --- | --- | --- | --- | --- | --- | --- | --- |
| Living with children under 18 | **Y** | **N** | **Y** | **N** | **Y** | **N** | **Y** | **N** | **Y** | **N** | **Y** | **N** | **P-value**  **(for total)** |
| If you were working before COVID-19, has COVID-19 created any inconvenience for you? | N=546 | N=709 | N=276 | N=337 | N=158 | N=472 | N=112 | N=414 | N=462 | N=467 | N=1,554 | N=2,399 |  |
| Loss of earnings | (N=545) 483 (91) | (N=703) 529 (79) | (N=239) 66 (44) | (N=317) 89 (37) | (N=144) 52 (46) | (N=440) 174 (43) | (N=98) 58 (61) | (N=398) 202 (54) | (N=428) 100 (30) | (N=439) 119 (31) | (N=1,454) 759 (62) | (N=2,297) 1,113 (53) | 0.005 |
| Loss of job | (N=525) 121 (27) | (N=666) 112 (19) | (N=227) 20 (26) | (N=305) 24 (10) | (N=139) 10 (13) | (N=412) 41 (9) | (N=92) 12 (9) | (N=379) 47 (14) | (N=409) 6 (3) | (N=423) 9 (3) | (N=1,392) 169 (18) | (N=2,185) 233 (12) | 0.008 |
| Reduction of  working hours | (N=531) 240 (47) | (N=679) 252 (38) | (N=230) 102 (55) | (N=316) 126 (50) | (N=145) 48 (38) | (N=425) 153 (39) | (N=99) 48 (52) | (N=385) 185 (49) | (N=427) 165 (45) | (N=435) 154 (38) | (N=1,432) 603 (47) | (N=2,240) 870 (41) | 0.047 |
| Closure of workplace | (N=528) 216 (43) | (N=679) 209 (30) | (N=247) 141 (66) | (N=315) 148 (44) | (N=151) 73 (46) | (N=440) 223 (52) | (N=96) 39 (44) | (N=388) 128 (38) | (N=413) 27 (7) | (N=420) 36 (9) | (N=1,435) 496 (38) | (N=2,242) 744 (35) | 0.268 |
| Did you continue to work during COVID-19? | (N=546) 412 (74) | (N=709) 607 (84) | (N=276) 242 (65) | (N=337) 290 (74) | (N=158) 124 (71) | (N=472) 336 (69) | (N=112) 85 (73) | (N=414) 303 (65) | (N=462) 386 (81) | (N=467) 382 (78) | (N=1,554) 1,249 (74) | (N=2,399) 1,918 (75) | 0.655 |
| What are/were your concerns if advised no physical contact/not allowed to go out/allowed to go out only for essential needs? | N=664 | N=812 | N=346 | N=481 | N=186 | N=823 | N=144 | N=568 | N=497 | N=537 | N=1,837 | N=3,221 |  |
| Financial | (N=660) 594 (92) | (N=806) 621 (80) | (N=323) 194 (59) | (N=452) 225 (62) | (N=174) 59 (35) | (N=776) 212 (32) | (N=135) 76 (61) | (N=543) 239 (37) | (N=486) 139 (33) | (N=529) 163 (24) | (N=1,778) 1,062 (64) | (N=3,106) 1,460 (47) | <0.001 |
| Professional/ career progression | (N=637) 230 (37) | (N=777) 377 (45) | (N=315) 182 (53) | (N=444) 236 (51) | (N=171) 58 (35) | (N=771) 140 (21) | (N=134) 46 (35) | (N=536) 178 (19) | (N=483) 98 (19) | (N=518) 121 (15) | (N=1,740) 614 (35) | (N=3,046) 1,052 (30) | 0.033 |

### Suppl. Table 8 Breakdown of economic impacts of COVID-19 and concerns by country and type of income

FBP = fixed salary, benefits/pension; CF = contract and freelance; O = other/no income. Values in cells are n (weighted %) of respondents who replied ‘yes’.

| Variable and categories | Thailand | | | Malaysia | | | UK | | | Italy | | | Slovenia | | | Total | | | |
| --- | --- | --- | --- | --- | --- | --- | --- | --- | --- | --- | --- | --- | --- | --- | --- | --- | --- | --- | --- |
| Type of income | **FBP** | **CF** | **O** | **FBP** | **CF** | **O** | **FBP** | **CF** | **O** | **FBP** | **CF** | **O** | **FBP** | **CF** | **O** | **FBP** | **CF** | **O** | **P-value**  **(for total)** |
| If you were working before COVID-19, has COVID-19 created any inconvenience for you? | N=495 | N=738 | N=22 | N=475 | N=125 | N=13 | N=397 | N=210 | N=23 | N=278 | N=228 | N=20 | N=788 | N=101 | N=40 | N=2,433 | N=1,402 | N=118 |  |
| Loss of earnings | (N=493) 320 (74) | (N=733) 674 (91) | (N=22) 18 (89) | (N=428) 69 (26) | (N=117) 79 (65) | (N=11) 7 (92) | (N=361) 91 (28) | (N=200) 125 (67) | (N=23) 10 (50) | (N=253) 87 (39) | (N=224) 157 (75) | (N=19) 16 (95) | (N=731) 128 (21) | (N=96) 70 (77) | (N=40) 21 (53) | (N=2,266) 695 (38) | (N=1,370) 1,105 (81) | (N=115) 72 (69) | <0.001 |
| Loss of job | (N=478) 78 (21) | (N=692) 148 (23) | (N=21) 7 (47) | (N=420) 18 (8) | (N=101) 24 (31) | (N=11) 2 (78) | (N=350) 20 (6) | (N=179) 30 (17) | (N=22) 1 (6) | (N=247) 6 (3) | (N=206) 45 (27) | (N=18) 8 (36) | (N=709) 6 (2) | (N=83) 5 (6) | (N=40) 4 (10) | (N=2,204) 128 (8) | (N=1,261) 252 (22) | (N=112) 22 (27) | <0.001 |
| Reduction of  working hours | (N=479) 226 (52) | (N=710) 259 (36) | (N=21) 7 (45) | (N=429) 163 (51) | (N=106) 60 (56) | (N=11) 5 (12) | (N=358) 89 (24) | (N=189) 102 (60) | (N=23) 10 (48) | (N=256) 111 (45) | (N=210) 113 (56) | (N=18) 9 (26) | (N=735) 227 (33) | (N=89) 67 (81) | (N=38) 25 (70) | (N=2,257) 816 (41) | (N=1,304) 601 (47) | (N=111) 56 (49) | 0.042 |
| Closure of workplace | (N=480) 195 (44) | (N=706) 224 (30) | (N=21) 6 (43) | (N=438) 214 (52) | (N=113) 67 (54) | (N=11) 8 (89) | (N=376) 188 (47) | (N=192) 98 (56) | (N=23) 10 (51) | (N=252) 63 (27) | (N=213) 94 (54) | (N=19) 10 (68) | (N=710) 33 (5) | (N=85) 20 (20) | (N=38) 10 (23) | (N=2,256) 693 (33) | (N=1,309) 503 (40) | (N=112) 44 (46) | 0.015 |
| Did you continue to work during COVID-19? | (N=495) 418 (83) | (N=738) 584 (77) | (N=22) 17 (78) | (N=475) 437 (83) | (N=125) 86 (42) | (N=13) 9 (25) | (N=397) 319 (79) | (N=210) 126 (57) | (N=23) 15 (62) | (N=278) 234 (81) | (N=228) 146 (51) | (N=20) 8 (15) | (N=788) 682 (84) | (N=101) 63 (57) | (N=40) 23 (59) | (N=2,433) 2,090 (82) | (N=1,402) 1,005 (65) | (N=118) 72 (53) | <0.001 |
| What are/were your concerns if advised no physical contact/not allowed to go out/allowed to go out only for essential needs? | N=546 | N=849 | N=81 | N=524 | N=158 | N=145 | N=705 | N=227 | N=77 | N=347 | N=244 | N=121 | N=847 | N=103 | N=84 | N=2,969 | N=1,581 | N=508 |  |
| Financial | (N=543) 402 (81) | (N=843) 753 (89) | (N=80) 60 (76) | (N=488) 231 (58) | (N=149) 110 (83) | (N=138) 78 (39) | (N=658) 131 (22) | (N=219) 116 (56) | (N=73) 24 (34) | (N=324) 102 (30) | (N=238) 165 (66) | (N=116) 48 (43) | (N=830) 190 (23) | (N=102) 74 (61) | (N=83) 38 (40) | (N=2,843) 1,056 (40) | (N=1,551) 1,218 (79) | (N=490) 248 (46) | <0.001 |
| Professional/ career progression | (N=530) 221 (43) | (N=804) 348 (41) | (N=80) 38 (37) | (N=481) 247 (41) | (N=142) 81 (71) | (N=136) 90 (56) | (N=657) 104 (17) | (N=212) 66 (36) | (N=73) 28 (40) | (N=319) 71 (15) | (N=235) 112 (38) | (N=116) 41 (22) | (N=821) 156 (14) | (N=97) 35 (23) | (N=83) 28 (33) | (N=2,808) 799 (24) | (N=1,490) 642 (43) | (N=488) 225 (40) | <0.001 |

### Suppl. Table 9 Breakdown of concerns if advised/not allowed physical contact by country

Values in cells are n (weighted %) of respondents who replied ‘yes’.

| Variable and categories | Thailand | Malaysia | UK | Italy | Slovenia | Total | P-value  (for total) |
| --- | --- | --- | --- | --- | --- | --- | --- |
| What are/were your concerns if advised no physical contact/not allowed to go out/allowed to go out only for essential needs? | N=1,476 | N=827 | N=1,009 | N=712 | N=1,034 | N=5,058 |  |
| Caring responsibilities (e.g. childcare, caring for elderly parents, not having access to care) | (N=1,454) 890 (62) | (N=772) 456 (57) | (N=946) 325 (31) | (N=681) 312 (46) | (N=1,006) 423 (35) | (N=4,859) 2,406 (47) | <0.001 |
| Physical health (e.g. not being able to attend doctor appointments, medication supply for illnesses,  lack of exercise) | (N=1,457) 910 (61) | (N=782) 501 (66) | (N=961) 587 (61) | (N=687) 393 (63) | (N=1,007) 437 (45) | (N=4,894) 2,828 (59) | <0.001 |
| Recreational (e.g. not being able to access recreational facilities like cinemas or restaurants, cancelled  sports or cultural events) | (N=1,425) 580 (38) | (N=763) 407 (49) | (N=963) 571 (58) | (N=683) 352 (47) | (N=1,011) 636 (65) | (N=4,845) 2,546 (51) | <0.001 |
| Sports (e.g. participating in competitive or professional sports activities) | (N=1,400) 546 (38) | (N=755) 302 (39) | (N=943) 214 (22) | (N=675) 174 (24) | (N=997) 331 (36) | (N=4,770) 1,567 (32) | <0.001 |
| Mental health and wellbeing (e.g. boredom, loneliness, anxiety, depression) | (N=1,427) 798 (55) | (N=769) 476 (61) | (N=970) 699 (75) | (N=691) 448 (60) | (N=1,008) 436 (43) | (N=4,865) 2,857 (58) | <0.001 |
| Living arrangements (e.g. not enough living space, passing on illness to family members, domestic abuse) | (N=1,419) 646 (45) | (N=753) 289 (46) | (N=943) 215 (24) | (N=674) 114 (16) | (N=999) 177 (15) | (N=4,788) 1,441 (31) | <0.001 |
| Infrastructure (e.g. access to transport, network services, internet access) | (N=1,409) 651 (46) | (N=750) 308 (45) | (N=935) 212 (24) | (N=672) 163 (28) | (N=996) 195 (19) | (N=4,762) 1,529 (33) | <0.001 |
| Social (e.g. not being able to see friends or attend social or family events) | (N=1,440) 768 (52) | (N=773) 474 (56) | (N=974) 768 (79) | (N=686) 525 (70) | (N=1,015) 725 (69) | (N=4,888) 3,260 (64) | <0.001 |
| Religious and spiritual (e.g. not being able to go to church, mosque, temple etc.) | (N=1,433) 591 (42) | (N=769) 393 (58) | (N=942) 162 (17) | (N=670) 95 (18) | (N=998) 201 (19) | (N=4,812) 1,442 (31) | <0.001 |

### Suppl. Table 10 Breakdown of concerns if advised/not allowed physical contact by country and gender

M = male; F = female; O = other/prefer not to say. Values in cells are n (weighted %) of respondents who replied ‘yes’.

| Variable and categories | Thailand | | | | Malaysia | | | UK | | | Italy | | | | Slovenia | | | Total | | | |
| --- | --- | --- | --- | --- | --- | --- | --- | --- | --- | --- | --- | --- | --- | --- | --- | --- | --- | --- | --- | --- | --- |
| Gender | **M** | **F** | **O** | **M** | | **F** | **O** | **M** | **F** | **O** | | **M** | **F** | **O** | **M** | **F** | **O** | **M** | **F** | **O** | **P-value (for total**  **M vs F)** |
| What are/were your concerns if advised no physical contact/not allowed to go out/allowed to go out only for essential needs? | N=704 | N=766 | N=6 | N=298 | | N=525 | N=4 | N=426 | N=572 | N=11 | | N=222 | N=490 | N=0 | N=366 | N=662 | N=6 | N=2,016 | N=3,015 | N=27 |  |
| Caring responsibilities | (N=697) 430 (61) | (N=751) 456 (62) | (N=6) 4 (67) | (N=282) 170 (53) | | (N=486) 284 (62) | (N=4) 2 (50) | (N=407) 124 (27) | (N=529) 197 (35) | (N=10) 4 (40) | | (N=213) 82 (36) | (N=468) 230 (56) |  | (N=356) 124 (25) | (N=644) 297 (44) | (N=6) 2 (33) | (N=1,955) 930 (42) | (N=2,878) 1,464 (52) | (N=26) 12 (46) | <0.001 |
| Physical health | (N=698) 443 (60) | (N=753) 463 (61) | (N=6) 4 (67) | (N=282) 184 (59) | | (N=496) 314 (74) | (N=4) 3 (75) | (N=414) 255 (62) | (N=537) 323 (61) | (N=10) 9 (90) | | (N=213) 106 (56) | (N=474) 287 (70) |  | (N=356) 148 (44) | (N=645) 287 (46) | (N=6) 2 (33) | (N=1,963) 1,136 (56) | (N=2,905) 1,674 (61) | (N=26) 18 (69) | 0.058 |
| Recreational | (N=681) 267 (39) | (N=738) 310 (38) | (N=6) 3 (50) | (N=275) 160 (54) | | (N=484) 246 (44) | (N=4) 1 (25) | (N=411) 253 (61) | (N=542) 309 (56) | (N=10) 9 (90) | | (N=215) 126 (54) | (N=468) 226 (41) |  | (N=359) 239 (71) | (N=646) 395 (59) | (N=6) 2 (33) | (N=1,941) 1,045 (54) | (N=2,878) 1,486 (47) | (N=26) 15 (58) | 0.007 |
| Sports | (N=670) 276 (40) | (N=724) 268 (35) | (N=6) 2 (33) | (N=275) 131 (47) | | (N=476) 170 (29) | (N=4) 1 (25) | (N=410) 104 (23) | (N=524) 105 (21) | (N=9) 5 (56) | | (N=212) 76 (32) | (N=463) 98 (17) |  | (N=353) 150 (44) | (N=638) 179 (28) | (N=6) 2 (33) | (N=1,920) 737 (38) | (N=2,825) 820 (27) | (N=25) 10 (40) | <0.001 |
| Mental health and wellbeing | (N=684) 377 (55) | (N=737) 418 (55) | (N=6) 3 (50) | (N=279) 167 (62) | | (N=486) 307 (61) | (N=4) 2 (50) | (N=414) 287 (73) | (N=545) 402 (77) | (N=11) 10 (91) | | (N=216) 122 (56) | (N=475) 326 (63) |  | (N=357) 128 (40) | (N=645) 305 (46) | (N=6) 3 (50) | (N=1,950) 1,081 (57) | (N=2,888) 1,758 (60) | (N=27) 18 (67) | 0.326 |
| Living arrangements | (N=679) 323 (46) | (N=734) 320 (44) | (N=6) 3 (50) | (N=275) 106 (48) | | (N=474) 182 (42) | (N=4) 1 (25) | (N=409) 79 (21) | (N=525) 131 (27) | (N=9) 5 (56) | | (N=211) 40 (19) | (N=463) 74 (14) |  | (N=354) 53 (12) | (N=639) 121 (18) | (N=6) 3 (50) | (N=1,928) 601 (31) | (N=2,835) 828 (31) | (N=25) 12 (48) | 0.948 |
| Infrastructure | (N=672) 316 (46) | (N=731) 332 (47) | (N=6) 3 (50) | (N=276) 129 (42) | | (N=470) 177 (48) | (N=4) 2 (50) | (N=407) 102 (27) | (N=520) 106 (21) | (N=8) 4 (50) | | (N=209) 51 (29) | (N=463) 112 (27) |  | (N=353) 60 (14) | (N=637) 133 (24) | (N=6) 2 (33) | (N=1,917) 658 (32) | (N=2,821) 860 (34) | (N=24) 11 (46) | 0.536 |
| Social | (N=689) 369 (53) | (N=745) 395 (51) | (N=6) 4 (67) | (N=280) 179 (62) | | (N=489) 294 (48) | (N=4) 1 (25) | (N=412) 321 (79) | (N=551) 438 (79) | (N=11) 9 (82) | | (N=215) 163 (66) | (N=471) 362 (74) |  | (N=360) 245 (70) | (N=649) 475 (69) | (N=6) 5 (83) | (N=1,956) 1,277 (65) | (N=2,905) 1,964 (63) | (N=27) 19 (70) | 0.503 |
| Religious and spiritual | (N=689) 290 (41) | (N=738) 298 (44) | (N=6) 3 (50) | (N=279) 140 (55) | | (N=486) 251 (61) | (N=4) 2 (50) | (N=408) 73 (19) | (N=524) 86 (14) | (N=10) 3 (30) | | (N=208) 33 (21) | (N=462) 62 (15) |  | (N=355) 77 (24) | (N=637) 124 (14) | (N=6) 0 (0) | (N=1,939) 613 (33) | (N=2,847) 821 (30) | (N=26) 8 (31) | 0.367 |

### Suppl. Table 11 Breakdown of concerns if advised/not allowed physical contact by country and age group

Values in cells are n (weighted %) of respondents who replied ‘yes’.

| Variable and categories | Thailand | | | Malaysia | | | UK | | | Italy | | | Slovenia | | | Total | | | |
| --- | --- | --- | --- | --- | --- | --- | --- | --- | --- | --- | --- | --- | --- | --- | --- | --- | --- | --- | --- |
| Age group | **18-34** | **35-64** | **65+** | **18-34** | **35-64** | **65+** | **18-34** | **35-64** | **65+** | **18-34** | **35-64** | **65+** | **18-34** | **35-64** | **65+** | **18-34** | **35-64** | **65+** | **P-value (for total)** |
| What are/were your concerns if advised no physical contact/not allowed to go out/allowed to go out only for essential needs? | N=223 | N=1,152 | N=101 | N=350 | N=442 | N=35 | N=140 | N=616 | N=253 | N=272 | N=383 | N=57 | N=308 | N=676 | N=50 | N=1,293 | N=3,269 | N=496 |  |
| Caring responsibilities | (N=217) 137 (71) | (N=1,138) 717 (64) | (N=99) 36 (37) | (N=333) 189 (56) | (N=407) 249 (57) | (N=32) 18 (66) | (N=131) 27 (20) | (N=581) 242 (41) | (N=234) 56 (23) | (N=270) 83 (30) | (N=361) 208 (55) | (N=50) 21 (43) | (N=304) 98 (30) | (N=656) 317 (44) | (N=46) 8 (16) | (N=1,255) 534 (46) | (N=3,143) 1,733 (53) | (N=461) 139 (32) | <0.001 |
| Physical health | (N=218) 150 (63) | (N=1,139) 712 (63) | (N=100) 48 (47) | (N=336) 205 (60) | (N=413) 269 (65) | (N=33) 27 (98) | (N=134) 76 (61) | (N=586) 354 (60) | (N=241) 157 (64) | (N=270) 137 (45) | (N=365) 217 (57) | (N=52) 39 (90) | (N=305) 131 (40) | (N=655) 284 (42) | (N=47) 22 (59) | (N=1,263) 699 (56) | (N=3,158) 1,836 (57) | (N=473) 293 (66) | 0.044 |
| Recreational | (N=212) 121 (47) | (N=1,118) 425 (35) | (N=95) 34 (34) | (N=331) 183 (55) | (N=403) 209 (44) | (N=29) 15 (40) | (N=136) 96 (66) | (N=589) 339 (57) | (N=238) 136 (53) | (N=270) 169 (66) | (N=362) 166 (44) | (N=51) 17 (38) | (N=302) 213 (71) | (N=663) 395 (60) | (N=46) 28 (70) | (N=1,251) 782 (59) | (N=3,135) 1,534 (47) | (N=459) 230 (48) | 0.003 |
| Sports | (N=212) 99 (47) | (N=1,096) 428 (38) | (N=92) 19 (18) | (N=329) 140 (47) | (N=397) 154 (31) | (N=29) 8 (29) | (N=133) 40 (28) | (N=575) 133 (22) | (N=235) 41 (14) | (N=269) 93 (40) | (N=356) 74 (19) | (N=50) 7 (20) | (N=301) 114 (41) | (N=653) 206 (36) | (N=43) 11 (31) | (N=1,244) 486 (42) | (N=3,077) 995 (31) | (N=449) 86 (21) | <0.001 |
| Mental health and wellbeing | (N=212) 146 (63) | (N=1,118) 613 (55) | (N=97) 39 (42) | (N=335) 230 (69) | (N=402) 227 (52) | (N=32) 19 (69) | (N=136) 118 (86) | (N=591) 439 (74) | (N=243) 142 (62) | (N=270) 191 (65) | (N=366) 227 (59) | (N=55) 30 (57) | (N=304) 169 (52) | (N=657) 253 (40) | (N=47) 14 (40) | (N=1,257) 854 (67) | (N=3,134) 1,759 (56) | (N=474) 244 (51) | <0.001 |
| Living arrangements | (N=213) 105 (50) | (N=1,111) 518 (48) | (N=95) 23 (26) | (N=330) 142 (47) | (N=394) 137 (45) | (N=29) 10 (40) | (N=134) 47 (35) | (N=576) 144 (24) | (N=233) 24 (10) | (N=270) 60 (21) | (N=353) 52 (16) | (N=51) 2 (14) | (N=304) 76 (22) | (N=651) 100 (17) | (N=44) 1 (1) | (N=1,251) 430 (38) | (N=3,085) 951 (32) | (N=452) 60 (15) | <0.001 |
| Infrastructure | (N=214) 117 (54) | (N=1,101) 502 (46) | (N=94) 32 (34) | (N=331) 149 (42) | (N=390) 152 (46) | (N=29) 7 (47) | (N=134) 37 (31) | (N=569) 133 (23) | (N=232) 42 (16) | (N=269) 59 (22) | (N=353) 91 (28) | (N=50) 13 (35) | (N=302) 63 (18) | (N=649) 121 (19) | (N=45) 11 (19) | (N=1,250) 425 (37) | (N=3,062) 999 (33) | (N=450) 105 (28) | 0.112 |
| Social | (N=216) 147 (59) | (N=1,126) 573 (50) | (N=98) 48 (46) | (N=334) 212 (55) | (N=408) 240 (55) | (N=31) 22 (60) | (N=136) 115 (83) | (N=592) 459 (77) | (N=246) 194 (79) | (N=268) 220 (84) | (N=366) 266 (69) | (N=52) 39 (63) | (N=304) 239 (79) | (N=662) 453 (65) | (N=49) 33 (69) | (N=1,258) 933 (69) | (N=3,154) 1,991 (62) | (N=476) 336 (64) | 0.156 |
| Religious and spiritual | (N=213) 86 (45) | (N=1,120) 468 (43) | (N=100) 37 (37) | (N=334) 180 (65) | (N=406) 198 (51) | (N=29) 15 (61) | (N=133) 14 (15) | (N=574) 111 (19) | (N=235) 37 (13) | (N=268) 27 (12) | (N=352) 64 (17) | (N=50) 4 (25) | (N=304) 51 (15) | (N=650) 142 (19) | (N=44) 8 (24) | (N=1,252) 358 (35) | (N=3,102) 983 (31) | (N=458) 101 (28) | 0.198 |

### Suppl. Table 12 Breakdown of concerns if advised/not allowed physical contact by country and education level

P/S = primary or lower/secondary education; T = tertiary education. Values in cells are n (weighted %) of respondents who replied ‘yes’.

| Variable and categories | Thailand | | | Malaysia | | | UK | | | Italy | | | Slovenia | | | Total | | |
| --- | --- | --- | --- | --- | --- | --- | --- | --- | --- | --- | --- | --- | --- | --- | --- | --- | --- | --- |
| Education level | **P/S** | **T** | **P/S** | | **T** | **P/S** | | **T** | **P/S** | | **T** | **P/S** | | **T** | **P/S** | | **T** | **P-value**  **(for total)** |
| What are/were your concerns if advised no physical contact/not allowed to go out/allowed to go out only for essential needs? | N=909 | N=567 | N=82 | | N=745 | N=247 | | N=762 | N=217 | | N=495 | N=202 | | N=832 | N=1,657 | | N=3,401 |  |
| Caring responsibilities | (N=894) 571 (63) | (N=560) 319 (57) | (N=74) 42 (57) | | (N=698) 414 (60) | (N=231) 78 (30) | | (N=715) 247 (32) | (N=204) 98 (47) | | (N=477) 214 (45) | (N=190) 67 (31) | | (N=816) 356 (40) | (N=1,593) 856 (49) | | (N=3,266) 1,550 (43) | 0.002 |
| Physical health | (N=894) 565 (60) | (N=563) 345 (63) | (N=75) 53 (66) | | (N=707) 448 (63) | (N=238) 146 (63) | | (N=723) 441 (59) | (N=208) 123 (66) | | (N=479) 270 (56) | (N=191) 78 (47) | | (N=816) 359 (43) | (N=1,606) 965 (60) | | (N=3,288) 1,863 (56) | 0.045 |
| Recreational | (N=870) 281 (34) | (N=555) 299 (57) | (N=72) 33 (47) | | (N=691) 374 (55) | (N=236) 120 (52) | | (N=727) 451 (64) | (N=204) 95 (45) | | (N=479) 257 (52) | (N=192) 123 (66) | | (N=819) 513 (62) | (N=1,574) 652 (46) | | (N=3,271) 1,894 (60) | <0.001 |
| Sports | (N=855) 317 (36) | (N=545) 229 (43) | (N=71) 25 (38) | | (N=684) 277 (43) | (N=230) 34 (17) | | (N=713) 180 (26) | (N=203) 44 (23) | | (N=472) 130 (27) | (N=190) 75 (39) | | (N=807) 256 (32) | (N=1,549) 495 (32) | | (N=3,221) 1,072 (32) | 0.953 |
| Mental health and wellbeing | (N=877) 486 (54) | (N=550) 312 (59) | (N=74) 46 (61) | | (N=695) 430 (62) | (N=238) 174 (76) | | (N=732) 525 (74) | (N=209) 137 (58) | | (N=482) 311 (63) | (N=190) 90 (45) | | (N=818) 346 (40) | (N=1,588) 933 (58) | | (N=3,277) 1,924 (60) | 0.256 |
| Living arrangements | (N=866) 422 (46) | (N=553) 224 (42) | (N=71) 32 (47) | | (N=682) 257 (39) | (N=232) 46 (23) | | (N=711) 169 (25) | (N=204) 37 (17) | | (N=470) 77 (15) | (N=189) 36 (14) | | (N=810) 141 (16) | (N=1,562) 573 (33) | | (N=3,226) 868 (26) | <0.001 |
| Infrastructure | (N=858) 396 (46) | (N=551) 255 (48) | (N=70) 32 (45) | | (N=680) 276 (44) | (N=229) 44 (23) | | (N=706) 168 (24) | (N=203) 55 (30) | | (N=469) 108 (23) | (N=189) 35 (18) | | (N=807) 160 (21) | (N=1,549) 562 (35) | | (N=3,213) 967 (29) | 0.004 |
| Social | (N=887) 440 (49) | (N=553) 328 (62) | (N=72) 38 (54) | | (N=701) 436 (63) | (N=242) 183 (77) | | (N=732) 585 (80) | (N=207) 157 (67) | | (N=479) 368 (77) | (N=194) 137 (69) | | (N=821) 588 (70) | (N=1,602) 955 (60) | | (N=3,286) 2,305 (73) | <0.001 |
| Religious and spiritual | (N=882) 391 (44) | (N=551) 200 (36) | (N=71) 42 (60) | | (N=698) 351 (51) | (N=232) 36 (17) | | (N=710) 126 (17) | (N=202) 36 (20) | | (N=468) 59 (13) | (N=190) 28 (18) | | (N=808) 173 (21) | (N=1,577) 533 (35) | | (N=3,235) 909 (24) | <0.001 |

### Suppl. Table 13 Breakdown of concerns if advised/not allowed physical contact by country and household size

Values in cells are n (weighted %) of respondents who replied ‘yes’.

| Variable and categories | Thailand | | | Malaysia | | | UK | | | Italy | | | Slovenia | | | Total | | |
| --- | --- | --- | --- | --- | --- | --- | --- | --- | --- | --- | --- | --- | --- | --- | --- | --- | --- | --- |
| Household size (number of persons in household) | **1-5** | **>=6** | **1-5** | | **>=6** | **1-5** | | **>=6** | **1-5** | | **>=6** | **1-5** | | **>=6** | **1-5** | | **>=6** | **P-value**  **(for total)** |
| What are/were your concerns if advised no physical contact/not allowed to go out/allowed to go out only for essential needs? | N=1,273 | N=203 | N=646 | | N=181 | N=994 | | N=15 | N=696 | | N=16 | N=953 | | N=81 | N=4,562 | | N=496 |  |
| Caring responsibilities | (N=1,251) 766 (62) | (N=203) 124 (59) | (N=603) 347 (61) | | (N=169) 109 (46) | (N=931) 312 (30) | | (N=15) 13 (80) | (N=665) 305 (46) | | (N=16) 7 (43) | (N=925) 388 (34) | | (N=81) 35 (42) | (N=4,375) 2,118 (47) | | (N=484) 288 (53) | 0.213 |
| Physical health | (N=1,256) 792 (62) | (N=201) 118 (54) | (N=609) 390 (71) | | (N=173) 111 (49) | (N=947) 579 (61) | | (N=14) 8 (65) | (N=671) 383 (63) | | (N=16) 10 (69) | (N=926) 408 (46) | | (N=81) 29 (42) | (N=4,409) 2,552 (60) | | (N=485) 276 (51) | 0.060 |
| Recreational | (N=1,229) 493 (38) | (N=196) 87 (39) | (N=596) 321 (49) | | (N=167) 86 (49) | (N=949) 565 (58) | | (N=14) 6 (53) | (N=667) 344 (47) | | (N=16) 8 (42) | (N=930) 594 (65) | | (N=81) 42 (55) | (N=4,371) 2,317 (51) | | (N=474) 229 (46) | 0.226 |
| Sports | (N=1,207) 479 (39) | (N=193) 67 (31) | (N=587) 238 (36) | | (N=168) 64 (46) | (N=929) 211 (22) | | (N=14) 3 (10) | (N=659) 168 (24) | | (N=16) 6 (34) | (N=917) 305 (36) | | (N=80) 26 (34) | (N=4,299) 1,401 (32) | | (N=471) 166 (36) | 0.383 |
| Mental health and wellbeing | (N=1,236) 697 (57) | (N=191) 101 (46) | (N=600) 369 (62) | | (N=169) 107 (61) | (N=956) 690 (75) | | (N=14) 9 (71) | (N=675) 436 (59) | | (N=16) 12 (80) | (N=927) 409 (44) | | (N=81) 27 (36) | (N=4,394) 2,601 (59) | | (N=471) 256 (51) | 0.096 |
| Living arrangements | (N=1,224) 574 (48) | (N=195) 72 (34) | (N=585) 219 (44) | | (N=168) 70 (50) | (N=928) 206 (23) | | (N=15) 9 (60) | (N=658) 112 (16) | | (N=16) 2 (20) | (N=918) 163 (15) | | (N=81) 14 (17) | (N=4,313) 1,274 (30) | | (N=475) 167 (38) | 0.072 |
| Infrastructure | (N=1,218) 564 (47) | (N=191) 87 (42) | (N=582) 233 (43) | | (N=168) 75 (48) | (N=921) 209 (24) | | (N=14) 3 (38) | (N=656) 160 (28) | | (N=16) 3 (26) | (N=915) 184 (19) | | (N=81) 11 (15) | (N=4,292) 1,350 (32) | | (N=470) 179 (40) | 0.113 |
| Social | (N=1,243) 667 (52) | (N=197) 101 (49) | (N=602) 369 (51) | | (N=171) 105 (68) | (N=959) 757 (79) | | (N=15) 11 (76) | (N=670) 511 (70) | | (N=16) 14 (78) | (N=934) 667 (70) | | (N=81) 58 (68) | (N=4,408) 2,971 (65) | | (N=480) 289 (60) | 0.270 |
| Religious and spiritual | (N=1,236) 511 (43) | (N=197) 80 (40) | (N=599) 296 (58) | | (N=170) 97 (57) | (N=928) 159 (17) | | (N=14) 3 (11) | (N=655) 92 (18) | | (N=15) 3 (36) | (N=917) 169 (18) | | (N=81) 32 (26) | (N=4,335) 1,227 (30) | | (N=477) 215 (43) | 0.005 |

### Suppl. Table 14 Breakdown of concerns if advised/not allowed physical contact by country and whether or not living with children under 18

Values in cells are n (weighted %) of respondents who replied ‘yes’.

| Variable and categories | Thailand | | Malaysia | | UK | | Italy | | Slovenia | | Total | | |
| --- | --- | --- | --- | --- | --- | --- | --- | --- | --- | --- | --- | --- | --- |
| Living with children under 18 | **Y** | **N** | **Y** | **N** | **Y** | **N** | **Y** | **N** | **Y** | **N** | **Y** | **N** | **P-value**  **(for total)** |
| What are/were your concerns if advised no physical contact/not allowed to go out/allowed to go out only for essential needs? | N=664 | N=812 | N=346 | N=481 | N=186 | N=823 | N=144 | N=568 | N=497 | N=537 | N=1,837 | N=3,221 |  |
| Caring responsibilities | (N=657) 487 (73) | (N=797) 403 (51) | (N=318) 217 (65) | (N=454) 239 (52) | (N=177) 109 (49) | (N=769) 216 (27) | (N=138) 88 (63) | (N=543) 224 (43) | (N=484) 278 (53) | (N=522) 145 (22) | (N=1,774) 1,179 (64) | (N=3,085) 1,227 (38) | <0.001 |
| Physical health | (N=659) 458 (67) | (N=798) 452 (55) | (N=321) 199 (60) | (N=461) 302 (70) | (N=179) 103 (61) | (N=782) 484 (61) | (N=138) 77 (56) | (N=549) 316 (64) | (N=484) 217 (44) | (N=523) 220 (46) | (N=1,781) 1,054 (59) | (N=3,113) 1,774 (59) | 0.984 |
| Recreational | (N=644) 220 (36) | (N=781) 360 (41) | (N=316) 169 (48) | (N=447) 238 (49) | (N=179) 102 (55) | (N=784) 469 (59) | (N=139) 66 (40) | (N=544) 286 (49) | (N=486) 284 (60) | (N=525) 352 (68) | (N=1,764) 841 (46) | (N=3,081) 1,705 (53) | 0.013 |
| Sports | (N=633) 267 (41) | (N=767) 279 (35) | (N=318) 137 (45) | (N=437) 165 (34) | (N=173) 52 (24) | (N=770) 162 (21) | (N=135) 38 (29) | (N=540) 136 (23) | (N=478) 175 (41) | (N=519) 156 (33) | (N=1,737) 669 (39) | (N=3,033) 898 (29) | <0.001 |
| Mental health and wellbeing | (N=641) 415 (63) | (N=786) 383 (48) | (N=318) 190 (56) | (N=451) 286 (65) | (N=180) 139 (80) | (N=790) 560 (74) | (N=139) 91 (60) | (N=552) 357 (60) | (N=481) 197 (44) | (N=527) 239 (43) | (N=1,759) 1,032 (59) | (N=3,106) 1,825 (58) | 0.841 |
| Living arrangements | (N=641) 366 (54) | (N=778) 280 (37) | (N=311) 118 (55) | (N=442) 171 (39) | (N=174) 56 (36) | (N=769) 159 (21) | (N=134) 24 (19) | (N=540) 90 (16) | (N=479) 93 (21) | (N=520) 84 (11) | (N=1,739) 657 (42) | (N=3,049) 784 (24) | <0.001 |
| Infrastructure | (N=632) 322 (50) | (N=777) 329 (43) | (N=310) 131 (48) | (N=440) 177 (42) | (N=172) 37 (29) | (N=763) 175 (23) | (N=135) 30 (18) | (N=537) 133 (30) | (N=477) 81 (17) | (N=519) 114 (20) | (N=1,726) 601 (37) | (N=3,036) 928 (31) | 0.018 |
| Social | (N=651) 347 (52) | (N=789) 421 (52) | (N=322) 194 (53) | (N=451) 280 (57) | (N=179) 141 (82) | (N=795) 627 (78) | (N=140) 109 (77) | (N=546) 416 (69) | (N=488) 341 (69) | (N=527) 384 (70) | (N=1,780) 1,132 (61) | (N=3,108) 2,128 (66) | 0.098 |
| Religious and spiritual | (N=641) 307 (49) | (N=792) 284 (36) | (N=319) 174 (58) | (N=450) 219 (58) | (N=171) 30 (19) | (N=771) 132 (16) | (N=133) 23 (20) | (N=537) 72 (18) | (N=479) 118 (20) | (N=519) 83 (18) | (N=1,743) 652 (39) | (N=3,069) 790 (28) | <0.001 |

### Suppl. Table 15 Breakdown of concerns if advised/not allowed physical contact by country and income type

FBP = fixed salary, benefits/pension; CF = contract and freelance; O = other/no income. Values in cells are n (weighted %) of respondents who replied ‘yes’.

| Variable and categories | Thailand | | | Malaysia | | | UK | | | Italy | | | Slovenia | | | Total | | | |
| --- | --- | --- | --- | --- | --- | --- | --- | --- | --- | --- | --- | --- | --- | --- | --- | --- | --- | --- | --- |
| Type of income | **FBP** | **CF** | **O** | **FBP** | **CF** | **O** | **FBP** | **CF** | **O** | **FBP** | **CF** | **O** | **FBP** | **CF** | **O** | **FBP** | **CF** | **O** | **P-value (for total)** |
| What are/were your concerns if advised no physical contact/not allowed to go out/allowed to go out only for essential needs? | N=546 | N=849 | N=81 | N=524 | N=158 | N=145 | N=705 | N=227 | N=77 | N=347 | N=244 | N=121 | N=847 | N=103 | N=84 | N=2,969 | N=1,581 | N=508 |  |
| Caring responsibilities | (N=540) 372 (72) | (N=836) 481 (57) | (N=78) 37 (39) | (N=490) 307 (58) | (N=145) 78 (64) | (N=137) 71 (47) | (N=661) 223 (32) | (N=213) 83 (32) | (N=72) 19 (26) | (N=328) 167 (49) | (N=236) 101 (41) | (N=117) 44 (44) | (N=826) 362 (36) | (N=97) 42 (31) | (N=83) 19 (23) | (N=2,845) 1,431 (47) | (N=1,527) 785 (51) | (N=487) 190 (38) | 0.028 |
| Physical health | (N=543) 381 (70) | (N=835) 482 (56) | (N=79) 47 (49) | (N=497) 324 (63) | (N=146) 89 (71) | (N=139) 88 (66) | (N=672) 415 (62) | (N=216) 124 (60) | (N=73) 48 (63) | (N=333) 204 (68) | (N=236) 122 (51) | (N=118) 67 (59) | (N=826) 345 (44) | (N=98) 56 (58) | (N=83) 36 (42) | (N=2,871) 1,669 (59) | (N=1,531) 873 (58) | (N=492) 286 (57) | 0.826 |
| Recreational | (N=535) 243 (43) | (N=812) 296 (35) | (N=78) 41 (42) | (N=483) 253 (46) | (N=143) 78 (48) | (N=137) 76 (56) | (N=671) 386 (54) | (N=218) 134 (65) | (N=74) 51 (71) | (N=331) 153 (46) | (N=236) 136 (50) | (N=116) 63 (47) | (N=828) 511 (62) | (N=101) 63 (75) | (N=82) 62 (75) | (N=2,848) 1,546 (52) | (N=1,510) 707 (46) | (N=487) 293 (58) | 0.024 |
| Sports | (N=531) 264 (53) | (N=791) 249 (29) | (N=78) 33 (32) | (N=474) 190 (35) | (N=145) 63 (47) | (N=136) 49 (39) | (N=660) 133 (18) | (N=213) 57 (28) | (N=70) 24 (30) | (N=325) 72 (22) | (N=234) 70 (26) | (N=116) 32 (28) | (N=818) 265 (34) | (N=96) 34 (46) | (N=83) 32 (45) | (N=2,808) 924 (32) | (N=1,479) 473 (32) | (N=483) 170 (36) | 0.582 |
| Mental health and wellbeing | (N=533) 339 (65) | (N=816) 410 (50) | (N=78) 49 (50) | (N=485) 297 (61) | (N=146) 86 (58) | (N=138) 93 (66) | (N=676) 485 (75) | (N=221) 157 (74) | (N=73) 57 (80) | (N=335) 213 (60) | (N=238) 147 (55) | (N=118) 88 (68) | (N=826) 346 (43) | (N=99) 42 (38) | (N=83) 48 (53) | (N=2,855) 1,680 (59) | (N=1,520) 842 (55) | (N=490) 335 (63) | 0.125 |
| Living arrangements | (N=533) 268 (51) | (N=808) 352 (43) | (N=78) 26 (27) | (N=474) 181 (48) | (N=142) 54 (55) | (N=137) 54 (27) | (N=655) 128 (19) | (N=216) 65 (34) | (N=72) 22 (30) | (N=325) 57 (17) | (N=233) 38 (16) | (N=116) 19 (14) | (N=821) 138 (14) | (N=95) 15 (13) | (N=83) 24 (29) | (N=2,808) 772 (27) | (N=1,494) 524 (38) | (N=486) 145 (26) | <0.001 |
| Infrastructure | (N=530) 279 (56) | (N=800) 335 (42) | (N=79) 37 (35) | (N=473) 179 (46) | (N=141) 55 (39) | (N=136) 74 (48) | (N=654) 134 (21) | (N=210) 56 (30) | (N=71) 22 (29) | (N=325) 74 (30) | (N=230) 56 (23) | (N=117) 33 (26) | (N=819) 157 (19) | (N=94) 15 (13) | (N=83) 23 (25) | (N=2,801) 823 (32) | (N=1,475) 517 (36) | (N=486) 189 (35) | 0.370 |
| Social | (N=537) 322 (58) | (N=824) 398 (48) | (N=79) 48 (51) | (N=491) 303 (55) | (N=146) 81 (59) | (N=136) 90 (52) | (N=681) 531 (78) | (N=219) 177 (79) | (N=74) 60 (81) | (N=335) 256 (72) | (N=233) 173 (63) | (N=118) 96 (78) | (N=834) 589 (68) | (N=98) 66 (67) | (N=83) 70 (86) | (N=2,878) 2,001 (67) | (N=1,520) 895 (58) | (N=490) 364 (67) | 0.004 |
| Religious and spiritual | (N=532) 235 (49) | (N=823) 326 (39) | (N=78) 30 (35) | (N=486) 254 (57) | (N=145) 68 (57) | (N=138) 71 (62) | (N=659) 121 (17) | (N=210) 31 (16) | (N=73) 10 (12) | (N=322) 43 (20) | (N=231) 36 (14) | (N=117) 16 (17) | (N=821) 168 (18) | (N=94) 22 (31) | (N=83) 11 (14) | (N=2,820) 821 (29) | (N=1,503) 483 (34) | (N=489) 138 (33) | 0.195 |

### Suppl. Table 16 Breakdown of maximum number of days that people thought they could cope by country

Values in cells are n (weighted %) of respondents who replied ‘yes’.

| Variable and categories | Thailand | Malaysia | UK | Italy | Slovenia | Total | P-value |
| --- | --- | --- | --- | --- | --- | --- | --- |
| What is the maximum number of days you think you could cope without meeting family or friends not living in your household in person? | N=1,476 | N=827 | N=1,009 | N=712 | N=1,034 | N=5,058 | <0.001 |
| 1 to 14 days | 957 (66) | 201 (31) | 192 (21) | 127 (23) | 261 (34) | 1,738 (39) |  |
| >14 to 28 days | 223 (13) | 110 (16) | 98 (11) | 95 (14) | 169 (16) | 695 (14) |  |
| 29 days+ | 296 (21) | 516 (52) | 719 (68) | 490 (63) | 604 (50) | 2,625 (47) |  |
| What is the maximum number of days you think you could cope with not going out in public, assuming that you have sufficient supplies of food, medicines and other essential items? | N=1,476 | N=827 | N=1,009 | N=712 | N=1,034 | N=5,058 | <0.001 |
| 1 to 14 days | 805 (54) | 270 (41) | 393 (40) | 304 (45) | 601 (61) | 2,373 (49) |  |
| >14 to 28 days | 249 (17) | 114 (16) | 124 (14) | 161 (21) | 151 (13) | 799 (16) |  |
| 29 days+ | 422 (29) | 443 (43) | 492 (46) | 247 (34) | 282 (26) | 1,886 (35) |  |
| What is the maximum number of days you think you could cope with going out only for essential needs/work? | N=1,476 | N=827 | N=1,009 | N=712 | N=1,034 | N=5,058 | <0.001 |
| 1 to 14 days | 808 (56) | 268 (40) | 272 (29) | 205 (33) | 310 (37) | 1,863 (41) |  |
| >14 to 28 days | 258 (17) | 98 (14) | 100 (10) | 110 (17) | 182 (18) | 748 (15) |  |
| 29 days+ | 410 (26) | 461 (46) | 637 (60) | 397 (51) | 542 (45) | 2,447 (44) |  |

### Suppl. Table 17 Breakdown of maximum number of days that people thought they could cope by country and gender

M = male; F = female; O = other/prefer not to say. Values in cells are n (weighted %) of respondents who replied ‘yes’.

| Variable and categories | Thailand | | | Malaysia | | | UK | | | Italy | | | Slovenia | | | Total | | | |
| --- | --- | --- | --- | --- | --- | --- | --- | --- | --- | --- | --- | --- | --- | --- | --- | --- | --- | --- | --- |
| Gender | **M** | **F** | **O** | **M** | **F** | **O** | **M** | **F** | **O** | **M** | **F** | **O** | **M** | **F** | **O** | **M** | **F** | **O** | **P-value (for total**  **M vs F)** |
| What is the maximum number of days you think you could cope without meeting family or friends not living in your household in person? | N=704 | N=766 | N=6 | N=298 | N=525 | N=4 | N=426 | N=572 | N=11 | N=222 | N=490 | N=0 | N=366 | N=662 | N=6 | N=2,016 | N=3,015 | N=27 | 0.381 |
| 1 to 14 days | 479 (66) | 476 (66) | 2 (33) | 68 (29) | 132 (34) | 1 (25) | 87 (23) | 102 (19) | 3 (27) | 46 (28) | 81 (18) |  | 113 (38) | 147 (31) | 1 (17) | 793 (40) | 938 (37) | 7 (26) |  |
| >14 to 28 days | 99 (12) | 123 (15) | 1 (17) | 40 (14) | 69 (18) | 1 (25) | 43 (13) | 54 (9) | 1 (9) | 28 (11) | 67 (17) |  | 49 (14) | 120 (18) | 0 (0) | 259 (13) | 433 (15) | 3 (11) |  |
| 29 days+ | 126 (23) | 167 (19) | 3 (50) | 190 (57) | 324 (48) | 2 (50) | 296 (64) | 416 (72) | 7 (64) | 148 (61) | 342 (65) |  | 204 (48) | 395 (51) | 5 (83) | 964 (47) | 1,644 (47) | 17 (63) |  |
| What is the maximum number of days you think you could cope with not going out in public, assuming that you have sufficient supplies of food, medicines and other essential items? | N=704 | N=766 | N=6 | N=298 | N=525 | N=4 | N=426 | N=572 | N=11 | N=222 | N=490 |  | N=366 | N=662 | N=6 | N=2,016 | N=3,015 | N=27 | 0.890 |
| 1 to 14 days | 398 (53) | 405 (55) | 2 (33) | 96 (41) | 173 (40) | 1 (25) | 170 (42) | 219 (38) | 4 (36) | 100 (48) | 204 (42) |  | 217 (57) | 382 (65) | 2 (33) | 981 (49) | 1,383 (50) | 9 (33) |  |
| >14 to 28 days | 116 (18) | 132 (16) | 1 (17) | 47 (18) | 66 (14) | 1 (25) | 53 (14) | 71 (13) | 0 (0) | 46 (18) | 115 (24) |  | 40 (14) | 111 (12) | 0 (0) | 302 (16) | 495 (16) | 2 (7) |  |
| 29 days+ | 190 (30) | 229 (29) | 3 (50) | 155 (41) | 286 (46) | 2 (50) | 203 (43) | 282 (49) | 7 (64) | 76 (34) | 171 (34) |  | 109 (29) | 169 (23) | 4 (67) | 733 (35) | 1,137 (35) | 16 (59) |  |
| What is the maximum number of days you think you could cope with going out only for essential needs/work? | N=704 | N=766 | N=6 | N=298 | N=525 | N=4 | N=426 | N=572 | N=11 | N=222 | N=490 |  | N=366 | N=662 | N=6 | N=2,016 | N=3,015 | N=27 | 0.680 |
| 1 to 14 days | 418 (57) | 388 (55) | 2 (33) | 94 (41) | 173 (38) | 1 (25) | 127 (32) | 141 (27) | 4 (36) | 72 (35) | 133 (31) |  | 125 (35) | 183 (40) | 2 (33) | 836 (42) | 1,018 (40) | 9 (33) |  |
| >14 to 28 days | 114 (17) | 142 (17) | 2 (33) | 35 (11) | 62 (17) | 1 (25) | 40 (10) | 60 (10) | 0 (0) | 31 (17) | 79 (17) |  | 73 (23) | 109 (13) | 0 (0) | 293 (16) | 452 (15) | 3 (11) |  |
| 29 days+ | 172 (25) | 236 (27) | 2 (33) | 169 (47) | 290 (45) | 2 (50) | 259 (58) | 371 (62) | 7 (64) | 119 (49) | 278 (52) |  | 168 (43) | 370 (47) | 4 (67) | 887 (42) | 1,545 (45) | 15 (56) |  |

### Suppl. Table 18 Breakdown of maximum number of days that people thought they could cope by country and age group

Values in cells are n (weighted %) of respondents who replied ‘yes’.

| Variable and categories | Thailand | | | Malaysia | | | UK | | | Italy | | | Slovenia | | | Total | | | |
| --- | --- | --- | --- | --- | --- | --- | --- | --- | --- | --- | --- | --- | --- | --- | --- | --- | --- | --- | --- |
| Age group | **18-34** | **35-64** | **65+** | **18-34** | **35-64** | **65+** | **18-34** | **35-64** | **65+** | **18-34** | **35-64** | **65+** | **18-34** | **35-64** | **65+** | **18-34** | **35-64** | **65+** | **P-value (for total)** |
| What is the maximum number of days you think you could cope without meeting family or friends not living in your household in person? | N=223 | N=1,152 | N=101 | N=350 | N=442 | N=35 | N=140 | N=616 | N=253 | N=272 | N=383 | N=57 | N=308 | N=676 | N=50 | N=1,293 | N=3,269 | N=496 | 0.409 |
| 1 to 14 days | 115 (57) | 774 (70) | 68 (67) | 96 (32) | 96 (25) | 9 (55) | 22 (22) | 112 (18) | 58 (24) | 37 (19) | 81 (26) | 9 (19) | 78 (29) | 167 (31) | 16 (49) | 348 (36) | 1,230 (39) | 160 (42) |  |
| >14 to 28 days | 29 (10) | 179 (15) | 15 (15) | 51 (19) | 53 (13) | 6 (22) | 16 (13) | 55 (10) | 27 (12) | 42 (20) | 42 (11) | 11 (17) | 49 (17) | 112 (15) | 8 (18) | 187 (10) | 441 (13) | 67 (16) |  |
| 29 days+ | 79 (33) | 199 (15) | 18 (18) | 203 (49) | 293 (62) | 20 (23) | 102 (65) | 449 (72) | 168 (64) | 193 (62) | 260 (63) | 37 (64) | 181 (54) | 397 (54) | 26 (34) | 758 (50) | 1,598 (48) | 269 (42) |  |
| What is the maximum number of days you think you could cope with not going out in public, assuming that you have sufficient supplies of food, medicines and other essential items? | N=223 | N=1,152 | N=101 | N=350 | N=442 | N=35 | N=140 | N=616 | N=253 | N=272 | N=383 | N=57 | N=308 | N=676 | N=50 | N=1,293 | N=3,269 | N=496 | 0.335 |
| 1 to 14 days | 113 (48) | 643 (58) | 49 (50) | 116 (42) | 141 (36) | 13 (56) | 62 (42) | 222 (37) | 109 (47) | 111 (45) | 170 (44) | 23 (47) | 192 (61) | 382 (59) | 27 (67) | 594 (47) | 1,558 (49) | 221 (53) |  |
| >14 to 28 days | 33 (17) | 192 (16) | 24 (20) | 43 (13) | 65 (17) | 6 (28) | 19 (17) | 85 (14) | 20 (9) | 65 (19) | 82 (19) | 14 (27) | 36 (11) | 107 (14) | 8 (15) | 196 (15) | 531 (16) | 72 (18) |  |
| 29 days+ | 77 (35) | 317 (26) | 28 (30) | 191 (45) | 236 (47) | 16 (16) | 59 (40) | 309 (50) | 124 (45) | 96 (36) | 131 (37) | 20 (26) | 80 (28) | 187 (28) | 15 (19) | 503 (37) | 1,180 (36) | 203 (29) |  |
| What is the maximum number of days you think you could cope with going out only for essential needs/work? | N=223 | N=1,152 | N=101 | N=350 | N=442 | N=35 | N=140 | N=616 | N=253 | N=272 | N=383 | N=57 | N=308 | N=676 | N=50 | N=1,293 | N=3,269 | N=496 | 0.255 |
| 1 to 14 days | 107 (52) | 648 (59) | 53 (56) | 91 (32) | 163 (43) | 14 (62) | 33 (28) | 161 (27) | 78 (36) | 62 (27) | 126 (36) | 17 (32) | 98 (34) | 189 (33) | 23 (51) | 391 (37) | 1,287 (42) | 185 (46) |  |
| >14 to 28 days | 43 (18) | 195 (17) | 20 (17) | 40 (13) | 54 (14) | 4 (15) | 17 (12) | 58 (10) | 25 (8) | 48 (20) | 52 (14) | 10 (20) | 53 (17) | 121 (17) | 8 (19) | 201 (16) | 480 (15) | 67 (16) |  |
| 29 days+ | 73 (30) | 309 (24) | 28 (27) | 219 (55) | 225 (43) | 17 (22) | 90 (60) | 397 (63) | 150 (56) | 162 (53) | 205 (51) | 30 (48) | 157 (49) | 366 (50) | 19 (29) | 701 (48) | 1,502 (43) | 244 (38) |  |

### Suppl. Table 19 Breakdown of maximum number of days that people thought they could cope by country and household size

Values in cells are n (weighted %) of respondents who replied ‘yes’.

| Variable and categories | Thailand | | | Malaysia | | | UK | | | Italy | | | Slovenia | | | Total | | |
| --- | --- | --- | --- | --- | --- | --- | --- | --- | --- | --- | --- | --- | --- | --- | --- | --- | --- | --- |
| Household size (number of persons in household) | **1-5** | **≥6** | **1-5** | | **≥6** | **1-5** | | **≥6** | **1-5** | | **≥6** | **1-5** | | **≥6** | **1-5** | | **≥6** | **P-value**  **(for total)** |
| What is the maximum number of days you think you could cope without meeting family or friends not living in your household in person? | N=1,273 | N=203 | N=646 | | N=181 | N=994 | | N=15 | N=696 | | N=16 | N=953 | | N=81 | N=4,562 | | N=496 | 0.499 |
| 1 to 14 days | 835 (68) | 122 (56) | 152 (30) | | 49 (36) | 191 (21) | | 1 (4) | 122 (23) | | 5 (44) | 247 (35) | | 14 (24) | 1,547 (38) | | 191 (43) |  |
| >14 to 28 days | 189 (13) | 34 (15) | 85 (16) | | 25 (17) | 98 (11) | | 0 (0) | 94 (15) | | 1 (3) | 156 (16) | | 13 (15) | 622 (14) | | 73 (15) |  |
| 29 days+ | 249 (19) | 47 (29) | 409 (54) | | 107 (47) | 705 (68) | | 14 (96) | 480 (63) | | 10 (53) | 550 (49) | | 54 (61) | 2,393 (48) | | 232 (42) |  |
| What is the maximum number of days you think you could cope with not going out in public, assuming that you have sufficient supplies of food, medicines and other essential items? | N=1,273 | N=203 | N=646 | | N=181 | N=994 | | N=15 | N=696 | | N=16 | N=953 | | N=81 | N=4,562 | | N=496 | 0.298 |
| 1 to 14 days | 712 (56) | 93 (43) | 209 (34) | | 61 (59) | 389 (40) | | 4 (40) | 296 (45) | | 8 (58) | 558 (62) | | 43 (55) | 2,164 (49) | | 209 (51) |  |
| >14 to 28 days | 211 (16) | 38 (23) | 86 (15) | | 28 (19) | 121 (13) | | 3 (23) | 159 (21) | | 2 (8) | 139 (13) | | 12 (12) | 716 (16) | | 83 (20) |  |
| 29 days+ | 350 (28) | 72 (34) | 351 (50) | | 92 (22) | 484 (46) | | 8 (37) | 241 (34) | | 6 (34) | 256 (25) | | 26 (32) | 1,682 (35) | | 204 (30) |  |
| What is the maximum number of days you think you could cope with going out only for essential needs/work? | N=1,273 | N=203 | N=646 | | N=181 | N=994 | | N=15 | N=696 | | N=16 | N=953 | | N=81 | N=4,562 | | N=496 | 0.134 |
| 1 to 14 days | 703 (57) | 105 (55) | 215 (37) | | 53 (51) | 269 (29) | | 3 (37) | 202 (33) | | 3 (29) | 292 (38) | | 18 (24) | 1,681 (40) | | 182 (49) |  |
| >14 to 28 days | 222 (18) | 36 (16) | 80 (15) | | 18 (9) | 100 (11) | | 0 (0) | 106 (17) | | 4 (20) | 170 (18) | | 12 (11) | 678 (16) | | 70 (12) |  |
| 29 days+ | 348 (26) | 62 (29) | 351 (48) | | 110 (40) | 625 (60) | | 12 (63) | 388 (51) | | 9 (51) | 491 (44) | | 51 (64) | 2,203 (44) | | 244 (39) |  |

### Suppl. Table 20 Breakdown of maximum number of days that people thought they could cope by country and whether or not living with children under 18

Y = living with children under 18; N = not living with children under 18. Values in cells are n (weighted %) of respondents who replied ‘yes’.

| Variable and categories | Thailand | | | Malaysia | | | UK | | | Italy | | | Slovenia | | | Total | | |
| --- | --- | --- | --- | --- | --- | --- | --- | --- | --- | --- | --- | --- | --- | --- | --- | --- | --- | --- |
| Living with children under 18 | **Y** | **N** | **Y** | | **N** | **Y** | | **N** | **Y** | | **N** | **Y** | | **N** | **Y** | | **N** | **P-value**  **(for total)** |
| What is the maximum number of days you think you could cope without meeting family or friends not living in your household in person? | N=664 | N=812 | N=346 | | N=481 | N=186 | | N=823 | N=144 | | N=568 | N=497 | | N=537 | N=1,837 | | N=3,221 | <0.001 |
| 1 to 14 days | 490 (72) | 467 (60) | 97 (40) | | 104 (25) | 24 (14) | | 168 (22) | 24 (18) | | 103 (24) | 115 (30) | | 146 (38) | 750 (46) | | 988 (35) |  |
| >14 to 28 days | 80 (10) | 143 (17) | 37 (12) | | 73 (19) | 18 (12) | | 80 (11) | 13 (9) | | 82 (16) | 79 (14) | | 90 (18) | 227 (12) | | 468 (16) |  |
| 29 days+ | 94 (18) | 202 (23) | 212 (47) | | 304 (56) | 144 (74) | | 575 (67) | 107 (73) | | 383 (61) | 303 (57) | | 301 (45) | 860 (42) | | 1,765 (50) |  |
| What is the maximum number of days you think you could cope with not going out in public, assuming that you have sufficient supplies of food, medicines and other essential items? | N=664 | N=812 | N=346 | | N=481 | N=186 | | N=823 | N=144 | | N=568 | N=497 | | N=537 | N=1,837 | | N=3,221 | <0.001 |
| 1 to 14 days | 412 (59) | 393 (49) | 120 (57) | | 150 (29) | 60 (36) | | 333 (41) | 62 (44) | | 242 (45) | 290 (62) | | 311 (60) | 944 (56) | | 1,429 (46) |  |
| >14 to 28 days | 100 (16) | 149 (18) | 45 (11) | | 69 (20) | 34 (19) | | 90 (12) | 33 (26) | | 128 (20) | 73 (13) | | 78 (14) | 285 (15) | | 514 (17) |  |
| 29 days+ | 152 (25) | 270 (33) | 181 (33) | | 262 (51) | 92 (46) | | 400 (46) | 49 (31) | | 198 (34) | 134 (25) | | 148 (26) | 608 (29) | | 1,278 (38) |  |
| What is the maximum number of days you think you could cope with going out only for essential needs/work? | N=664 | N=812 | N=346 | | N=481 | N=186 | | N=823 | N=144 | | N=568 | N=497 | | N=537 | N=1,837 | | N=3,221 | 0.004 |
| 1 to 14 days | 407 (63) | 401 (51) | 117 (47) | | 151 (35) | 33 (21) | | 239 (31) | 42 (35) | | 163 (32) | 139 (35) | | 171 (39) | 738 (47) | | 1,125 (38) |  |
| >14 to 28 days | 112 (16) | 146 (18) | 37 (8) | | 61 (18) | 17 (8) | | 83 (11) | 20 (11) | | 90 (18) | 90 (16) | | 92 (18) | 276 (14) | | 472 (16) |  |
| 29 days+ | 145 (21) | 265 (31) | 192 (45) | | 269 (47) | 136 (71) | | 501 (58) | 82 (53) | | 315 (50) | 268 (49) | | 274 (42) | 823 (40) | | 1,624 (46) |  |

### Suppl. Table 21 Breakdown of maximum number of days that people thought they could cope by country and education level

P/S = primary or lower/secondary education; T = tertiary education. Values in cells are n (weighted %) of respondents who replied ‘yes’.

| Variable and categories | Thailand | | | Malaysia | | | UK | | | Italy | | | Slovenia | | | Total | | |
| --- | --- | --- | --- | --- | --- | --- | --- | --- | --- | --- | --- | --- | --- | --- | --- | --- | --- | --- |
| Education level | **P/S** | **T** | **P/S** | | **T** | **P/S** | | **T** | **P/S** | | **T** | **P/S** | | **T** | **P/S** | | **T** | **P-value**  **(for total)** |
| What is the maximum number of days you think you could cope without meeting family or friends not living in your household in person? | N=909 | N=567 | N=82 | | N=745 | N=247 | | N=762 | N=217 | | N=495 | N=202 | | N=832 | N=1,657 | | N=3,401 | <0.001 |
| 1 to 14 days | 659 (69) | 298 (51) | 27 (33) | | 174 (23) | 55 (24) | | 137 (18) | 53 (26) | | 74 (16) | 69 (41) | | 192 (24) | 863 (45) | | 875 (25) |  |
| >14 to 28 days | 122 (12) | 101 (17) | 15 (17) | | 95 (13) | 30 (13) | | 68 (9) | 31 (15) | | 64 (13) | 33 (16) | | 136 (16) | 231 (15) | | 464 (13) |  |
| 29 days+ | 128 (18) | 168 (32) | 40 (50) | | 476 (64) | 162 (63) | | 557 (73) | 133 (59) | | 357 (72) | 100 (43) | | 504 (60) | 563 (41) | | 2,062 (62) |  |
| What is the maximum number of days you think you could cope with not going out in public, assuming that you have sufficient supplies of food, medicines and other essential items? | N=909 | N=567 | N=82 | | N=745 | N=247 | | N=762 | N=217 | | N=495 | N=202 | | N=832 | N=1,657 | | N=3,401 | 0.004 |
| 1 to 14 days | 541 (56) | 264 (47) | 34 (43) | | 236 (32) | 101 (41) | | 292 (40) | 95 (46) | | 209 (43) | 119 (63) | | 482 (58) | 890 (51) | | 1,483 (45) |  |
| >14 to 28 days | 144 (17) | 105 (18) | 15 (17) | | 99 (13) | 31 (15) | | 93 (13) | 41 (20) | | 120 (24) | 23 (12) | | 128 (15) | 254 (16) | | 545 (16) |  |
| 29 days+ | 224 (28) | 198 (35) | 33 (40) | | 410 (55) | 115 (44) | | 377 (48) | 81 (34) | | 166 (33) | 60 (25) | | 222 (27) | 513 (33) | | 1,373 (39) |  |
| What is the maximum number of days you think you could cope with going out only for essential needs/work? | N=909 | N=567 | N=82 | | N=745 | N=247 | | N=762 | N=217 | | N=495 | N=202 | | N=832 | N=1,657 | | N=3,401 | <0.001 |
| 1 to 14 days | 564 (59) | 244 (43) | 35 (43) | | 233 (29) | 87 (35) | | 185 (24) | 70 (35) | | 135 (29) | 75 (42) | | 235 (31) | 831 (46) | | 1,032 (30) |  |
| >14 to 28 days | 156 (17) | 102 (19) | 12 (14) | | 86 (11) | 26 (10) | | 74 (10) | 39 (18) | | 71 (14) | 33 (17) | | 149 (18) | 266 (16) | | 482 (14) |  |
| 29 days+ | 189 (24) | 221 (38) | 35 (43) | | 426 (59) | 134 (54) | | 503 (66) | 108 (48) | | 289 (57) | 94 (41) | | 448 (51) | 560 (38) | | 1,887 (56) |  |

### Suppl. Table 22 Breakdown of maximum number of days that people thought they could cope by country and type of income

FBP = fixed salary, benefits/pension; CF = contract and freelance; O = other. Values in cells are n (weighted %) of respondents who replied ‘yes’.

| Variable and categories | Thailand | | | | Malaysia | | | | UK | | | | Italy | | | | Slovenia | | | | Total | | | |
| --- | --- | --- | --- | --- | --- | --- | --- | --- | --- | --- | --- | --- | --- | --- | --- | --- | --- | --- | --- | --- | --- | --- | --- | --- |
| Type of income | **FBP** | **CF** | **O** | **FBP** | | **CF** | **O** | **FBP** | | **CF** | **O** | **FBP** | | **CF** | **O** | **FBP** | | **CF** | **O** | **FBP** | | **CF** | **O** | **P-value**  **(for total)** |
| What is the maximum number of days you think you could cope without meeting family or friends not living in your household in person? | N=546 | N=849 | N=81 | N=524 | | N=158 | N=145 | N=705 | | N=227 | N=77 | N=347 | | N=244 | N=121 | N=847 | | N=103 | N=84 | N=2,969 | | N=1,581 | N=508 | <0.001 |
| 1 to 14 days | 344 (64) | 577 (69) | 36 (43) | 135 (23) | | 35 (37) | 31 (48) | 134 (22) | | 36 (17) | 22 (24) | 58 (22) | | 47 (27) | 22 (18) | 208 (34) | | 35 (44) | 18 (26) | 879 (33) | | 730 (50) | 129 (34) |  |
| >14 to 28 days | 74 (11) | 134 (14) | 15 (17) | 57 (15) | | 24 (16) | 29 (19) | 69 (11) | | 25 (14) | 4 (7) | 46 (15) | | 30 (12) | 19 (15) | 141 (17) | | 19 (16) | 9 (9) | 387 (14) | | 232 (14) | 76 (14) |  |
| 29 days+ | 128 (25) | 138 (16) | 30 (41) | 332 (62) | | 99 (47) | 85 (33) | 502 (68) | | 166 (69) | 51 (69) | 243 (63) | | 167 (60) | 80 (66) | 498 (49) | | 49 (40) | 57 (65) | 1,703 (53) | | 619 (35) | 303 (51) |  |
| What is the maximum number of days you think you could cope with not going out in public, assuming that you have sufficient supplies of food, medicines and other essential items? | N=546 | N=849 | N=81 | N=524 | | N=158 | N=145 | N=705 | | N=227 | N=77 | N=347 | | N=244 | N=121 | N=847 | | N=103 | N=84 | N=2,969 | | N=1,581 | N=508 | 0.471 |
| 1 to 14 days | 313 (55) | 461 (55) | 31 (39) | 183 (38) | | 46 (39) | 41 (49) | 273 (40) | | 87 (41) | 33 (42) | 147 (45) | | 108 (47) | 49 (40) | 485 (560 | | 66 (75) | 50 (59) | 1,401 (49) | | 768 (51) | 204 (46) |  |
| >14 to 28 days | 85 (16) | 148 (17) | 16 (20) | 70 (18) | | 22 (17) | 22 (10) | 90 (13) | | 28 (17) | 6 (9) | 84 (24) | | 55 (17) | 22 (14) | 129 (14) | | 12 (7) | 10 (14) | 458 (16) | | 265 (16) | 76 (13) |  |
| 29 days+ | 148 (29) | 240 (28) | 34 (40) | 271 (44) | | 90 (44) | 82 (41) | 342 (47) | | 112 (43) | 38 (49) | 116 (30) | | 81 (36) | 50 (46) | 233 (27) | | 25 (18) | 24 (27) | 1,110 (35) | | 548 (33) | 228 (41) |  |
| What is the maximum number of days you think you could cope with going out only for essential needs/work? | N=546 | N=849 | N=81 | N=524 | | N=158 | N=145 | N=705 | | N=227 | N=77 | N=347 | | N=244 | N=121 | N=847 | | N=103 | N=84 | N=2,969 | | N=1,581 | N=508 | <0.001 |
| 1 to 14 days | 297 (59) | 478 (56) | 33 (43) | 181 (38) | | 56 (53) | 31 (29) | 186 (29) | | 64 (31) | 22 (22) | 99 (33) | | 78 (34) | 28 (27) | 250 (38) | | 41 (45) | 19 (27) | 1,013 (39) | | 717 (49) | 133 (30) |  |
| >14 to 28 days | 81 (16) | 159 (18) | 18 (23) | 54 (14) | | 23 (4) | 21 (25) | 68 (10) | | 20 (10) | 12 (16) | 55 (18) | | 30 (12) | 25 (19) | 150 (17) | | 17 (21) | 15 (17) | 408 (15) | | 249 (14) | 91 (21) |  |
| 29 days+ | 168 (25) | 212 (26) | 30 (34) | 289 (48) | | 79 (43) | 93 (46) | 451 (61) | | 143 (58) | 43 (62) | 193 (49) | | 136 (53) | 68 (54) | 447 (45) | | 45 (34) | 50 (57) | 1,548 (46) | | 615 (37) | 284 (50) |  |

### Suppl. Table 23 Breakdown of behavioural changes and acceptance of government public health measures by country

Values in cells are n (weighted %) of respondents who replied ‘yes’.

| Variable and categories | Thailand | Malaysia | UK | Italy | Slovenia | Total | P-value |
| --- | --- | --- | --- | --- | --- | --- | --- |
|  | N=1,476 | N=827 | N=1,009 | N=712 | N=1,034 | N=5,058 |  |
| Did you change your social behaviour before the implementation of government restrictions? | 1,374 (93) | 538 (64) | 712 (68) | 356 (47) | 584 (47) | 3,564 (67) | <0.001 |
| If you answered 'yes' to the previous question: how did you change your social behaviour? |  |  |  |  |  |  |  |
| No physical contact with anyone | (N=1,374) 1,302 (94) | (N=506) 362 (82) | (N=657) 325 (51) | (N=342) 243 (74) | (N=576) 516 (93) | (N=3,455) 2,748 (82) | <0.001 |
| No physical contact only with elderly and those with serious underlying medical conditions | (N=1,374) 1,200 (88) | (N=494) 292 (63) | (N=644) 393 (60) | (N=332) 272 (79) | (N=566) 516 (91) | (N=3,410) 2,673 (79) | <0.001 |
| Going out only for essential needs | (N=1,374) 1,291 (94) | (N=525) 489 (95) | (N=681) 571 (83) | (N=346) 263 (82) | (N=562) 381 (71) | (N=3,488) 2,995 (87) | <0.001 |
| Moving home to stay with parents/relatives | (N=1,374) 677 (54) | (N=489) 99 (26) | (N=627) 30 (8) | (N=326) 27 (6) | (N=552) 33 (5) | (N=3,368) 866 (30) | <0.001 |
| Use of personal protection equipment (e.g. masks and gloves) | (N=1,374) 1,334 (96) | (N=527) 488 (95) | (N=651) 225 (33) | (N=339) 165 (55) | (N=564) 366 (67) | (N=3,455) 2,578 (76) | <0.001 |
| Use of sanitizer products and alcohol | (N=1,374) 1,321 (95) | (N=529) 504 (96) | (N=685) 559 (83) | (N=350) 307 (91) | (N=569) 521 (94) | (N=3,507) 3,212 (92) | <0.001 |
| “I would comply with government enforced quarantine/ isolation/social distancing.” | N=1,476 | N=827 | N=1,009 | N=712 | N=1,034 | N=5,058 | <0.001 |
| Agree | 1,344 (92) | 708 (86) | 822 (80) | 606 (78) | 871 (75) | 4,351 (83) |  |
| Neither agree nor disagree | 92 (5) | 18 (0) | 48 (4) | 36 (7) | 68 (14) | 262 (6) |  |
| Disagree | 40 (3) | 101 (14) | 139 (15) | 70 (15) | 95 (11) | 445 (10) |  |
| “I would enter voluntary quarantine/isolation/social distancing for social/self-responsibility.” | N=1,476 | N=827 | N=1,009 | N=712 | N=1,034 | N=5,058 | <0.001 |
| Agree | 1,354 (92) | 674 (81) | 815 (78) | 566 (76) | 838 (76) | 4,247 (82) |  |
| Neither agree nor disagree | 100 (7) | 48 (4) | 50 (5) | 59 (10) | 91 (13) | 348 (8) |  |
| Disagree | 22 (1) | 105 (15) | 144 (17) | 87 (14) | 105 (11) | 463 (10) |  |
| How much do you agree with quarantine/isolation/social distancing? “It is a necessary strategy to help control COVID-19.” | N=1,476 | N=827 | N=1,009 | N=712 | N=1,034 | N=5,058 | <0.001 |
| Agree | 1,383 (94) | 739 (88) | 853 (83) | 608 (80) | 846 (74) | 4,429 (85) |  |
| Neither agree nor disagree | 65 (4) | 12 (0) | 27 (3) | 28 (5) | 76 (11) | 208 (5) |  |
| Disagree | 28 (2) | 76 (12) | 129 (14) | 76 (15) | 112 (15) | 421 (10) |  |

### Suppl. Table 24 Breakdown of behavioural changes and acceptance of government public health measures by country and gender

M = male; F = female; O = other/prefer not to say. Values in cells are n (weighted %) of respondents who replied ‘yes’.

| Variable and categories | Thailand | | | | Malaysia | | | | UK | | | | Italy | | | | Slovenia | | | | Total | | | |
| --- | --- | --- | --- | --- | --- | --- | --- | --- | --- | --- | --- | --- | --- | --- | --- | --- | --- | --- | --- | --- | --- | --- | --- | --- |
| Gender | **M** | **F** | **O** | **M** | | **F** | **O** | **M** | | **F** | **O** | **M** | | **F** | **O** | **M** | | **F** | **O** | **M** | | **F** | **O** | **P-value (for total**  **M vs F)** |
|  | N=704 | N=766 | N=6 | N=298 | | N=525 | N=4 | N=426 | | N=572 | N=11 | N=222 | | N=490 | N=0 | N=366 | | N=662 | N=6 | N=2,016 | | N=3,015 | N=27 |  |
| Did you change your social behaviour before the implementation of government restrictions? | 660 (94) | 709 (92) | 5 (83) | 184 (60) | | 351 (68) | 3 (75) | 288 (64) | | 415 (71) | 9 (82) | 99 (43) | | 257 (52) |  | 179 (42) | | 402 (51) | 3 (50) | 1,410 (65) | | 2,134 (70) | 20 (74) | 0.039 |
| If you answered 'yes' to the previous question: how did you change your social behaviour? |  |  |  |  | |  |  |  | |  |  |  | |  |  |  | |  |  |  | |  |  |  |
| No physical contact with anyone | (N=660) 626 (93) | (N=709) 671 (95) | (N=5) 5 (100) | (N=173) 122 (75) | | (N=330) 237 (87) | (N=3) 3 (100) | (N=271) 141 (51) | | (N=379) 181 (50) | (N=7) 3 (43) | (N=94) 63 (68) | | (N=248) 180 (78) |  | (N=175) 162 (94) | | (N=398) 351 (892 | (N=3) 3 (100) | (N=1,373) 1,114 (80) | | (N=2,064) 1,620 (83) | (N=18) 14 (78) | 0.227 |
| No physical contact only with elderly and those with serious underlying medical conditions | (N=660) 584 (88) | (N=709) 611 (89) | (N=5) 5 (100) | (N=170) 104 (59) | | (N=321) 186 (67) | (N=3) 2 (67) | (N=268) 148 (58) | | (N=370) 243 (62) | (N=6) 2 (33) | (N=90) 75 (75) | | (N=242) 197 (81) |  | (N=171) 152 (88) | | (N=392) 361 (94) | (N=3) 3 (100) | (N=1,359) 1,063 (77) | | (N=2,034) 1,598 (81) | (N=17) 12 (71) | 0.124 |
| Going out only for essential needs | (N=660) 612 (93) | (N=709) 674 (94) | (N=5) 5 (100) | (N=177) 164 (91) | | (N=345) 322 (99) | (N=3) 3 (100) | (N=277) 234 (84) | | (N=396) 330 (82) | (N=8) 7 (88) | (N=95) 71 (84) | | (N=251) 192 (81) |  | (N=172) 113 (65) | | (N=387) 265 (76) | (N=3) 3 (100) | (N=1,381) 1,194 (87) | | (N=2,088) 1,783 (88) | (N=19) 18 (95) | 0.327 |
| Moving home to stay with parents/relatives | (N=660) 359 (59) | (N=709) 316 (49) | (N=5) 2 (40) | (N=167) 39 (27) | | (N=319) 59 (24) | (N=3) 1 (33) | (N=267) 8 (3) | | (N=354) 22 (11) | (N=6) 0 (0) | (N=91) 7 (3) | | (N=235) 20 (9) |  | (N=167) 11 (3) | | (N=382) 21 (6) | (N=3) 1 (33) | (N=1,352) 424 (32) | | (N=1,999) 438 (28) | (N=17) 4 (24) | 0.207 |
| Use of personal protection equipment (e.g. masks and gloves) | (N=660) 639 (97) | (N=709) 690 (95) | (N=5) 5 (100) | (N=178) 160 (96) | | (N=346) 325 (95) | (N=3) 3 (100) | (N=272) 101 (33) | | (N=371) 121 (33) | (N=8) 3 (38) | (N=93) 38 (59) | | (N=246) 127 (52) |  | (N=173) 122 (73) | | (N=388) 241 (63) | (N=3) 3 (100) | (N=1,376) 1,060 (78) | | (N=2,060) 1,504 (74) | (N=19) 14 (74) | 0.079 |
| Use of sanitizer products and alcohol | (N=660) 628 (95) | (N=709) 688 (95) | (N=5) 5 (100) | (N=178) 167 (96) | | (N=348) 334 (96) | (N=3) 3 (100) | (N=278) 223 (80) | | (N=398) 329 (85) | (N=9) 7 (78) | (N=96) 80 (92) | | (N=254) 227 (91) |  | (N=173) 164 (94) | | (N=393) 354 (94) | (N=3) 3 (100) | (N=1,385) 1,262 (92) | | (N=2,102) 1,932 (93) | (N=20) 18 (90) | 0.474 |
| “I would comply with government enforced quarantine/ isolation/social distancing.” | N=704 | N=766 | N=6 | N=298 | | N=525 | N=4 | N=426 | | N=572 | N=11 | N=222 | | N=490 |  | N=366 | | N=662 | N=6 | N=2,016 | | N=3,015 | N=27 | 0.631 |
| Agree | 636 (92) | 705 (93) | 3 (50) | 262 (93) | | 442 (78) | 4 (100) | 334 (76) | | 480 (85) | 8 (73) | 176 (69) | | 430 (86) |  | 295 (75) | | 571 (75) | 5 (83) | 1,703 (82) | | 2,628 (84) | 20 (74) |  |
| Neither agree nor disagree | 49 (6) | 40 (4) | 3 (50) | 9 (1) | | 9 (0) | 0 (0) | 26 (6) | | 19 (3) | 3 (27) | 14 (10) | | 22 (5) |  | 24 (10) | | 44 (17) | 0 (0) | 122 (6) | | 134 (6) | 6 (22) |  |
| Disagree | 19 (2) | 21 (3) | 0 (0) | 27 (7) | | 74 (22) | 0 (0) | 66 (18) | | 73 (12) | 0 (0) | 32 (21) | | 38 (9) |  | 47 (15) | | 47 (8) | 1 (17) | 191 (11) | | 253 (10) | 1 (4) |  |
| “I would enter voluntary quarantine/isolation/social distancing for social/self-responsibility.” | N=704 | N=766 | N=6 | N=298 | | N=525 | N=4 | N=426 | | N=572 | N=11 | N=222 | | N=490 |  | N=366 | | N=662 | N=6 | N=2,016 | | N=3,015 | N=27 | 0.761 |
| Agree | 644 (91) | 707 (92) | 3 (50) | 258 (93) | | 412 (68) | 4 (100) | 340 (78) | | 465 (78) | 10 (91) | 163 (67) | | 403 (85) |  | 285 (76) | | 548 (77) | 5 (83) | 1,690 (83) | | 2,535 (81) | 22 (81) |  |
| Neither agree nor disagree | 50 (8) | 47 (7) | 3 (50) | 14 (1) | | 34 (8) | 0 (0) | 22 (5) | | 27 (5) | 1 (9) | 21 (14) | | 38 (6) |  | 36 (9) | | 55 (15) | 0 (0) | 143 (7) | | 201 (8) | 4 (15) |  |
| Disagree | 10 (1) | 12 (1) | 0 (0) | 26 (6) | | 79 (25) | 0 (0) | 64 (17) | | 80 (16) | 0 (0) | 38 (19) | | 49 (9) |  | 45 (15) | | 59 (8) | 1 (17) | 183 (10) | | 279 (10) | 1 (4) |  |
| How much do you agree with quarantine/isolation/social distancing? “It is a necessary strategy to help control COVID-19.” | N=704 | N=766 | N=6 | N=298 | | N=525 | N=4 | N=426 | | N=572 | N=11 | N=222 | | N=490 |  | N=366 | | N=662 | N=6 | N=2,016 | | N=3,015 | N=27 | 0.191 |
| Agree | 653 (93) | 725 (95) | 5 (83) | 272 (93) | | 463 (83) | 4 (100) | 342 (77) | | 502 (88) | 9 (82) | 169 (68) | | 439 (91) |  | 285 (75) | | 557 (74) | 4 (67) | 1,721 (83) | | 2,686 (87) | 22 (81) |  |
| Neither agree nor disagree | 38 (5) | 26 (3) | 1 (17) | 6 (0) | | 6 (0) | 0 (0) | 16 (4) | | 11 (3) | 0 (0) | 15 (9) | | 13 (2) |  | 28 (7) | | 47 (15) | 1 (17) | 103 (5) | | 103 (5) | 2 (7) |  |
| Disagree | 13 (1) | 15 (2) | 0 (0) | 20 (6) | | 56 (17) | 0 (0) | 68 (19) | | 59 (10) | 2 (18) | 38 (23) | | 38 (8) |  | 53 (18) | | 58 (12) | 1 (17) | 192 (12) | | 226 (9) | 3 (11) |  |

### Suppl. Table 25 Breakdown of behavioural changes and acceptance of government public health measures by country and education level

P/S = primary or lower/secondary education; T = tertiary education. Values in cells are n (weighted %) of respondents who replied ‘yes’.

| Variable and Categories | Thailand | | | Malaysia | | | UK | | | Italy | | | Slovenia | | | Total | | |
| --- | --- | --- | --- | --- | --- | --- | --- | --- | --- | --- | --- | --- | --- | --- | --- | --- | --- | --- |
| Education level | **P/S** | **T** | **P/S** | | **T** | **P/S** | | **T** | **P/S** | | **T** | **P/S** | | **T** | **P/S** | | **T** | **P-value**  **(for total)** |
|  | N=909 | N=567 | N=82 | | N=745 | N=247 | | N=762 | N=217 | | N=495 | N=202 | | N=832 | N=1,657 | | N=3,401 |  |
| Did you change your social behaviour before the implementation of government restrictions? | 849 (93) | 525 (92) | 52 (64) | | 486 (65) | 147 (60) | | 565 (74) | 99 (46) | | 257 (52) | 99 (41) | | 485 (56) | 1,246 (67) | | 2,318 (69) | 0.369 |
| If you answered 'yes' to the previous question: how did you change your social behaviour? |  |  |  | |  |  | |  |  | |  |  | |  |  | |  |  |
| No physical contact with anyone | (N=849) 816 (95) | (N=525) 486 (91) | (N=47) 41 (85) | | (N=459) 321 (70) | (N=138) 80 (59) | | (N=519) 245 (45) | (N=90) 67 (76) | | (N=252) 176 (71) | (N=97) 92 (96) | | (N=479) 424 (90) | (N=1,221) 1,096 (87) | | (N=2,234) 1,652 (70) | <0.001 |
| No physical contact only with elderly and those with serious underlying medical conditions | (N=849) 771 (90) | (N=525) 429 (81) | (N=43) 29 (64) | | (N=451) 263 (59) | (N=131) 76 (58) | | (N=513) 317 (61) | (N=87) 73 (77) | | (N=245) 199 (82) | (N=91) 83 (93) | | (N=475) 433 (90) | (N=1,201) 1,032 (81) | | (N=2,209) 1,641 (74) | 0.003 |
| Going out only for essential needs | (N=849) 798 (94) | (N=525) 493 (92) | (N=49) 47 (96) | | (N=476) 442 (93) | (N=143) 122 (84) | | (N=538) 449 (82) | (N=93) 69 (84) | | (N=253) 194 (79) | (N=93) 66 (75) | | (N=469) 315 (67) | (N=1,227) 1,102 (90) | | (N=2,261) 1,893 (82) | <0.001 |
| Moving home to stay with parents/relatives | (N=849) 515 (58) | (N=525) 162 (32) | (N=42) 11 (26) | | (N=447) 88 (23) | (N=131) 5 (8) | | (N=496) 25 (8) | (N=84) 10 (6) | | (N=242) 17 (6) | (N=91) 4 (3) | | (N=461) 29 (6) | (N=1,197) 545 (37) | | (N=2,171) 321 (15) | <0.001 |
| Use of personal protection equipment (e.g. masks and gloves) | (N=849) 819 (96) | (N=525) 515 (98) | (N=49) 47 (96) | | (N=478) 441 (91) | (N=136) 55 (35) | | (N=515) 170 (32) | (N=89) 49 (59) | | (N=250) 116 (47) | (N=94) 57 (67) | | (N=470) 309 (68) | (N=1,217) 1,027 (82) | | (N=2,238) 1,551 (62) | <0.001 |
| Use of sanitizer products and alcohol | (N=849) 813 (95) | (N=525) 508 (97) | (N=48) 46 (96) | | (N=481) 458 (95) | (N=142) 120 (83) | | (N=543) 439 (81) | (N=94) 84 (94) | | (N=256) 223 (87) | (N=96) 92 (96) | | (N=473) 429 (92) | (N=1,229) 1,155 (94) | | (N=2,278) 2,057 (89) | <0.001 |
| “I would comply with government enforced quarantine/ isolation/social distancing.” | N=909 | N=567 | N=82 | | N=745 | N=247 | | N=762 | N=217 | | N=495 | N=202 | | N=832 | N=1,657 | | N=3,401 | 0.315 |
| Agree | 843 (93) | 501 (87) | 70 (85) | | 638 (87) | 190 (77) | | 632 (83) | 178 (75) | | 428 (84) | 148 (68) | | 723 (87) | 1,429 (82) | | 2,922 (85) |  |
| Neither agree nor disagree | 43 (4) | 49 (10) | 0 (0) | | 18 (3) | 14 (5) | | 34 (4) | 9 (7) | | 27 (7) | 22 (19) | | 46 (6) | 88 (7) | | 174 (6) |  |
| Disagree | 23 (3) | 17 (3) | 12 (15) | | 89 (11) | 43 (18) | | 96 (13) | 30 (17) | | 40 (9) | 32 (14) | | 63 (7) | 140 (11) | | 305 (9) |  |
| “I would enter voluntary quarantine/isolation/social distancing for social/self-responsibility.” | N=909 | N=567 | N=82 | | N=745 | N=247 | | N=762 | N=217 | | N=495 | N=202 | | N=832 | N=1,657 | | N=3,401 | 0.370 |
| Agree | 842 (92) | 512 (89) | 65 (80) | | 609 (83) | 180 (73) | | 635 (83) | 165 (75) | | 401 (80) | 151 (72) | | 687 (82) | 1,403 (81) | | 2,844 (84) |  |
| Neither agree nor disagree | 55 (7) | 45 (10) | 3 (4) | | 45 (6) | 17 (6) | | 33 (4) | 24 (11) | | 35 (7) | 24 (15) | | 67 (9) | 123 (8) | | 225 (7) |  |
| Disagree | 12 (1) | 10 (2) | 14 (16) | | 91 (11) | 50 (21) | | 94 (13) | 28 (14) | | 59 (13) | 27 (13) | | 78 (9) | 131 (11) | | 332 (10) |  |
| How much do you agree with quarantine/isolation/social distancing? “It is a necessary strategy to help control COVID-19.” | N=909 | N=567 | N=82 | | N=745 | N=247 | | N=762 | N=217 | | N=495 | N=202 | | N=832 | N=1,657 | | N=3,401 | 0.304 |
| Agree | 858 (95) | 525 (91) | 72 (88) | | 667 (90) | 201 (80) | | 652 (85) | 179 (78) | | 429 (84) | 145 (768 | | 701 (85) | 1,455 (84) | | 2,974 (87) |  |
| Neither agree nor disagree | 34 (4) | 31 (7) | 0 (0) | | 12 (2) | 8 (4) | | 19 (3) | 6 (5) | | 22 (5) | 23 (14) | | 53 (6) | 71 (5) | | 137 (5) |  |
| Disagree | 17 (2) | 11 (2) | 10 (12) | | 66 (8) | 38 (17) | | 91 (12) | 32 (17) | | 44 (10) | 34 (19) | | 78 (9) | 131 (11) | | 290 (9) |  |

### Suppl. Table 26 Breakdown of behavioural changes and acceptance of government public health measures by age group

Values in cells are n (weighted %) of respondents who replied ‘yes’.

| Variable and categories | Thailand | | | | Malaysia | | | | UK | | | | Italy | | | | Slovenia | | | | Total | | | |
| --- | --- | --- | --- | --- | --- | --- | --- | --- | --- | --- | --- | --- | --- | --- | --- | --- | --- | --- | --- | --- | --- | --- | --- | --- |
| Age group | **18-34** | **35-64** | **65+** | **18-34** | | **35-64** | **65+** | **18-34** | | **35-64** | **65+** | **18-34** | | **35-64** | **65+** | **18-34** | | **35-64** | **65+** | **18-34** | | **35-64** | **65+** | **P-value**  **(for total)** |
|  | N=223 | N=1,152 | N=101 | N=350 | | N=442 | N=35 | N=140 | | N=616 | N=253 | N=272 | | N=383 | N=57 | N=308 | | N=676 | N=50 | N=1,293 | | N=3,269 | N=496 |  |
| Did you change your social behaviour before the implementation of government restrictions? | 202 (92) | 1,079 (94) | 93 (93) | 233 (63) | | 287 (71) | 18 (37) | 104 (71) | | 448 (69) | 160 (61) | 124 (44) | | 202 (44) | 30 (57) | 178 (54) | | 386 (53) | 20 (25) | 841 (70) | | 2,402 (70) | 321 (57) | 0.004 |
| If you answered 'yes' to the previous question: how did you change your social behaviour? |  |  |  |  | |  |  |  | |  |  |  | |  |  |  | |  |  |  | |  |  |  |
| No physical contact with anyone | (N=202) 180 (91) | (N=1,079) 1,037 (96) | (N=93) 85 (90) | (N=225) 156 (84) | | (N=265) 193 (80) | (N=16) 13 (81) | (N=99) 35 (43) | | (N=412) 200 (51) | (N=146) 90 (61) | (N=120) 79 (72) | | (N=196) 143 (74) | (N=26) 21 (75) | (N=176) 151 (87) | | (N=380) 345 (94) | (N=20) 20 (100) | (N=822) 601 (78) | | (N=2,332) 1,918 (84) | (N=301) 229 (82) | 0.204 |
| No physical contact only with elderly and those with serious underlying medical conditions | (N=202) 168 (88) | (N=1,079) 956 (90) | (N=93) 76 (83) | (N=218) 127 (65) | | (N=261) 158 (61) | (N=15) 7 (73) | (N=98) 60 (60) | | (N=416) 271 (65) | (N=130) 62 (46) | (N=120) 100 (89) | | (N=187) 150 (80) | (N=25) 22 (69) | (N=174) 163 (90) | | (N=374) 340 (92) | (N=18) 13 (87) | (N=812) 618 (78) | | (N=2,317) 1,875 (81) | (N=281) 180 (73) | 0.152 |
| Going out only for essential needs | (N=202) 186 (94) | (N=1,079) 1,022 (95) | (N=93) 83 (89) | (N=230) 212 (98) | | (N=278) 262 (94) | (N=17) 15 (82) | (N=102) 79 (76) | | (N=427) 362 (86) | (N=152) 130 (86) | (N=121) 79 (68) | | (N=198) 159 (79) | (N=27) 25 (99) | (N=174) 102 (55) | | (N=370) 266 (75) | (N=18) 13 (87) | (N=829) 658 (85) | | (N=2,352) 2,071 (88) | (N=307) 266 (89) | 0.153 |
| Moving home to stay with parents/relatives | (N=202) 88 (59) | (N=1,079) 556 (56) | (N=93) 33 (34) | (N=219) 65 (38) | | (N=256) 32 (16) | (N=14) 2 (22) | (N=98) 21 (21) | | (N=398) 8 (2) | (N=131) 1 (2) | (N=120) 16 (11) | | (N=184) 11 (7) | (N=22) 0 (0) | (N=172) 16 (8) | | (N=363) 17 (4) | (N=17) 0 (0) | (N=811) 206 (37) | | (N=2,280) 624 (29) | (N=277) 36 (17) | <0.001 |
| Use of personal protection equipment (e.g. masks and gloves) | (N=202) 198 (98) | (N=1,079) 1,050 (97) | (N=93) 86 (90) | (N=230) 212 (93) | | (N=279) 262 (99) | (N=18) 14 (80) | (N=100) 23 (20) | | (N=417) 157 (40) | (N=134) 45 (35) | (N=121) 48 (39) | | (N=191) 100 (54) | (N=27) 17 (69) | (N=174) 88 (52) | | (N=371) 260 (68) | (N=19) 18 (97) | (N=827) 569 (72) | | (N=2,337) 1,829 (79) | (N=291) 180 (74) | 0.067 |
| Use of sanitizer products and alcohol | (N=202) 197 (96) | (N=1,079) 1,037 (96) | (N=93) 87 (91) | (N=230) 218 (94) | | (N=281) 271 (99) | (N=18) 15 (81) | (N=102) 88 (84) | | (N=436) 352 (82) | (N=147) 119 (84) | (N=122) 103 (84) | | (N=199) 177 (90) | (N=29) 27 (99) | (N=174) 157 (92) | | (N=377) 346 (94) | (N=18) 18 (100) | (N=830) 763 (92) | | (N=2,372) 2,183 (93) | (N=305) 266 (91) | 0.613 |
| “I would comply with government enforced quarantine/ isolation/social distancing.” | N=223 | N=1,152 | N=101 | N=350 | | N=442 | N=35 | N=140 | | N=616 | N=253 | N=272 | | N=383 | N=57 | N=308 | | N=676 | N=50 | N=1,293 | | N=3,269 | N=496 | 0.003 |
| Agree | 189 (90) | 1,058 (92) | 97 (96) | 307 (82) | | 371 (88) | 30 (91) | 120 (85) | | 493 (78) | 209 (80) | 247 (88) | | 311 (77) | 48 (72) | 272 (85) | | 559 (75) | 40 (65) | 1,135 (86) | | 2,792 (83) | 424 (80) |  |
| Neither agree nor disagree | 28 (8) | 63 (5) | 1 (1) | 7 (1) | | 11 (1) | 0 (0) | 3 (1) | | 33 (6) | 12 (5) | 7 (2) | | 24 (5) | 5 (14) | 16 (7) | | 44 (8) | 8 (34) | 61 (4) | | 175 (5) | 26 (13) |  |
| Disagree | 6 (2) | 31 (3) | 3 (3) | 36 (18) | | 60 (11) | 5 (9) | 17 (14) | | 90 (17) | 32 (14) | 18 (10) | | 48 (17) | 4 (14) | 20 (8) | | 73 (17) | 2 (1) | 97 (10) | | 302 (12) | 46 (8) |  |
| “I would enter voluntary quarantine/isolation/social distancing for social/self-responsibility.” | N=223 | N=1,152 | N=101 | N=350 | | N=442 | N=35 | N=140 | | N=616 | N=253 | N=272 | | N=383 | N=57 | N=308 | | N=676 | N=50 | N=1,293 | | N=3,269 | N=496 | 0.327 |
| Agree | 188 (86) | 1,068 (93) | 98 (96) | 294 (79) | | 353 (86) | 27 (68) | 114 (79) | | 497 (78) | 204 (78) | 211 (70) | | 306 (75) | 49 (84) | 247 (80) | | 550 (75) | 41 (74) | 1,054 (80) | | 2,774 (83) | 419 (82) |  |
| Neither agree nor disagree | 33 (13) | 64 (5) | 3 (4) | 23 (7) | | 23 (1) | 2 (9) | 6 (4) | | 30 (5) | 14 (7) | 28 (15) | | 28 (8) | 3 (10) | 28 (9) | | 57 (11) | 6 (20) | 118 (9) | | 202 (6) | 28 (10) |  |
| Disagree | 2 (1) | 20 (2) | 0 (0) | 33 (15) | | 66 (13) | 6 (24) | 20 (17) | | 89 (17) | 35 (15) | 33 (16) | | 49 (17) | 5 (6) | 33 (11) | | 69 (13) | 3 (7) | 121 (11) | | 293 (11) | 49 (8) |  |
| How much do you agree with quarantine/isolation/social distancing? “It is a necessary strategy to help control COVID-19.” | N=223 | N=1,152 | N=101 | N=350 | | N=442 | N=35 | N=140 | | N=616 | N=253 | N=272 | | N=383 | N=57 | N=308 | | N=676 | N=50 | N=1,293 | | N=3,269 | N=496 | 0.271 |
| Agree | 203 (93) | 1,083 (94) | 97 (96) | 313 (85) | | 393 (89) | 33 (100) | 120 (83) | | 521 (83) | 212 (82) | 243 (86) | | 315 (78) | 50 (79) | 254 (79) | | 549 (76) | 43 (67) | 1,133 (86) | | 2,861 (85) | 435 (82) |  |
| Neither agree nor disagree | 18 (7) | 45 (4) | 2 (2) | 5 (0) | | 6 (0) | 1 (0) | 3 (3) | | 16 (3) | 8 (4) | 10 (4) | | 14 (3) | 4 (11) | 28 (12) | | 45 (7) | 3 (18) | 64 (5) | | 126 (4) | 18 (8) |  |
| Disagree | 2 (0) | 24 (2) | 2 (2) | 32 (15) | | 43 (11) | 1 (0) | 17 (14) | | 79 (15) | 33 (14) | 19 (10) | | 54 (19) | 3 (10) | 26 (10) | | 82 (17) | 4 (15) | 96 (9) | | 282 (11) | 43 (10) |  |

### Suppl. Table 27 Breakdown of behavioural changes and acceptance of government public health measures by self-reported level of understanding of COVID-19

H = high/very high/expert level; S = some; N = a little/none at all. Values in cells are n (weighted %) of respondents who replied ‘yes’.

| Variable and categories | Thailand | | | Malaysia | | | UK | | | Italy | | | Slovenia | | | Total | | | |
| --- | --- | --- | --- | --- | --- | --- | --- | --- | --- | --- | --- | --- | --- | --- | --- | --- | --- | --- | --- |
| Self-reported level of understanding of COVID-19 | **H** | **S** | **N** | **H** | **S** | **N** | **H** | **S** | **N** | **H** | **S** | **N** | **H** | **S** | **N** | **H** | **S** | **N** | **P-value**  **(for total)** |
|  | N=965 | N=459 | N=52 | N=435 | N=359 | N=33 | N=647 | N=336 | N=26 | N=368 | N=324 | N=20 | N=713 | N=279 | N=42 | N=3,128 | N=1,757 | N=173 |  |
| Did you change your social behaviour before the implementation of government restrictions? | 898 (94) | 430 (92) | 46 (91) | 285 (64) | 232 (66) | 21 (58) | 468 (69) | 232 (66) | 12 (68) | 200 (52) | 146 (43) | 10 (60) | 429 (52) | 137 (37) | 18 (46) | 2,280 (70) | 1,177 (64) | 107 (65) | 0.091 |
| If you answered 'yes' to the previous question: how did you change your social behaviour? |  |  |  |  |  |  |  |  |  |  |  |  |  |  |  |  |  |  |  |
| No physical contact with anyone | (N=898) 849 (94) | (N=430) 411 (95) | (N=46) 42 (9187) | (N=272) 204 (90) | (N=214) 143 (73) | (N=20) 15 (69) | (N=428) 221 (53) | (N=217) 99 (47) | (N=12) 5 (52) | (N=194) 137 (78) | (N=138) 99 (67) | (N=10) 7 (88) | (N=423) 380 (95) | (N=135) 119 (87) | (N=18) 17 (96) | (N=2,215) 1,791 (85) | (N=1,134) 871 (77) | (N=106) 86 (78) | 0.033 |
| No physical contact only with elderly and those with serious underlying medical conditions | (N=898) 765 (87) | (N=430) 394 (92) | (N=46) 41 (87) | (N=266) 162 (63) | (N=209) 119 (60) | (N=19) 11 (74) | (N=417) 261 (61) | (N=215) 128 (59) | (N=12) 4 (49) | (N=192) 163 (85) | (N=130) 101 (67) | (N=10) 8 (94) | (N=418) 379 (91) | (N=131) 122 (92) | (N=17) 15 (95) | (N=2,191) 1,730 (80) | (N=1,115) 864 (77) | (N=104) 79 (79) | 0.744 |
| Going out only for essential needs | (N=898) 844 (93) | (N=430) 405 (95) | (N=46) 42 (87) | (N=280) 266 (99) | (N=225) 205 (89) | (N=20) 18 (99) | (N=444) 381 (86) | (N=225) 182 (80) | (N=12) 8 (66) | (N=196) 145 (80) | (N=140) 109 (83) | (N=10) 9 (95) | (N=415) 283 (72) | (N=129) 87 (73) | (N=18) 11 (60) | (N=2,233) 1,919 (88) | (N=1,149) 988 (87) | (N=106) 88 (84) | 0.711 |
| Moving home to stay with parents/relatives | (N=898) 345 (45) | (N=430) 298 (67) | (N=46) 34 (73) | (N=261) 45 (24) | (N=209) 48 (25) | (N=19) 6 (40) | (N=404) 17 (5) | (N=212) 12 (10) | (N=11) 1 (24) | (N=189) 17 (6) | (N=127) 9 (7) | (N=10) 1 (10) | (N=405) 19 (3) | (N=129) 14 (9) | (N=18) 0 (0) | (N=2,157) 443 (25) | (N=1,107) 381 (36) | (N=104) 42 (42) | <0.001 |
| Use of personal protection equipment (e.g. masks and gloves) | (N=898) 874 (97) | (N=430) 418 (96) | (N=46) 42 (81) | (N=280) 266 (99) | (N=227) 203 (90) | (N=20) 19 (99) | (N=421) 153 (38) | (N=218) 68 (28) | (N=12) 4 (17) | (N=194) 90 (46) | (N=135) 69 (66) | (N=10) 6 (66) | (N=416) 289 (71) | (N=130) 71 (59) | (N=18) 6 (38) | (N=2,209) 1,672 (78) | (N=1,140) 829 (74) | (N=106) 77 (69) | 0.172 |
| Use of sanitizer products and alcohol | (N=898) 863 (96) | (N=430) 416 (95) | (N=46) 42 (81) | (N=281) 270 (99) | (N=228) 215 (91) | (N=20) 19 (100) | (N=447) 374 (85) | (N=226) 179 (85) | (N=12) 6 (30) | (N=198) 170 (90) | (N=142) 129 (93) | (N=10) 8 (94) | (N=418) 385 (95) | (N=133) 125 (95) | (N=18) 11 (70) | (N=2,242) 2,062 (94) | (N=1,159) 1,064 (92) | (N=106) 86 (78) | <0.001 |
| “I would comply with government enforced quarantine/ isolation/social distancing.” | N=965 | N=459 | N=52 | N=435 | N=359 | N=33 | N=647 | N=336 | N=26 | N=368 | N=324 | N=20 | N=713 | N=279 | N=42 | N=3,128 | N=1,757 | N=173 | 0.370 |
| Agree | 903 (95) | 402 (88) | 39 (81) | 378 (93) | 305 (79) | 25 (76) | 511 (79) | 291 (83) | 20 (87) | 303 (76) | 284 (79) | 19 (97) | 607 (75) | 232 (75) | 32 (70) | 2,702 (85) | 1,514 (82) | 135 (80) |  |
| Neither agree nor disagree | 39 (3) | 44 (9) | 9 (10) | 5 (0) | 9 (1) | 4 (1) | 29 (3) | 18 (6) | 1 (2) | 17 (4) | 18 (11) | 1 (3) | 45 (16) | 19 (10) | 4 (7) | 135 (6) | 108 (7) | 19 (4) |  |
| Disagree | 23 (2) | 13 (3) | 4 (9) | 52 (7) | 45 (20) | 4 (23) | 107 (18) | 27 (12) | 5 (11) | 48 (21) | 22 (10) | 0 (0) | 61 (9) | 28 (15) | 6 (24) | 291 (10) | 135 (11) | 19 (16) |  |
| “I would enter voluntary quarantine/isolation/social distancing for social/self-responsibility.” | N=965 | N=459 | N=52 | N=435 | N=359 | N=33 | N=647 | N=336 | N=26 | N=368 | N=324 | N=20 | N=713 | N=279 | N=42 | N=3,128 | N=1,757 | N=173 | 0.091 |
| Agree | 909 (95) | 401 (85) | 44 (90) | 357 (86) | 294 (76) | 23 (75) | 516 (78) | 284 (80) | 15 (60) | 293 (78) | 258 (74) | 15 (91) | 587 (78) | 219 (74) | 32 (69) | 2,662 (84) | 1,456 (79) | 129 (77) |  |
| Neither agree nor disagree | 41 (4) | 51 (13) | 8 (10) | 21 (1) | 21 (10) | 6 (1) | 29 (5) | 18 (5) | 3 (8) | 27 (8) | 30 (12) | 2 (6) | 58 (14) | 26 (9) | 7 (23) | 176 (6) | 146 (10) | 26 (8) |  |
| Disagree | 15 (1) | 7 (1) | 0 (0) | 57 (13) | 44 (14) | 4 (23) | 102 (17) | 34 (15) | 8 (32) | 48 (15) | 36 (13) | 3 (4) | 68 (9) | 34 (17) | 3 (7) | 290 (9) | 155 (11) | 18 (15) |  |
| How much do you agree with quarantine/isolation/social distancing? “It is a necessary strategy to help control COVID-19.” | N=965 | N=459 | N=52 | N=435 | N=359 | N=33 | N=647 | N=336 | N=26 | N=368 | N=324 | N=20 | N=713 | N=279 | N=42 | N=3,128 | N=1,757 | N=173 | 0.688 |
| Agree | 920 (96) | 418 (91) | 45 (90) | 392 (91) | 319 (85) | 28 (86) | 540 (82) | 293 (83) | 20 (85) | 304 (77) | 285 (82) | 19 (82) | 589 (73) | 226 (78) | 31 (72) | 2,745 (85) | 1,541 (85) | 143 (84) |  |
| Neither agree nor disagree | 26 (2) | 33 (8) | 6 (8) | 5 (0) | 5 (0) | 2 (1) | 16 (3) | 10 (3) | 1 (2) | 10 (2) | 18 (9) | 0 (0) | 45 (12) | 27 (9) | 4 (7) | 102 (4) | 93 (6) | 13 (4) |  |
| Disagree | 19 (1) | 8 (2) | 1 (2) | 38 (9) | 35 (15) | 3 (13) | 91 (15) | 33 (13) | 5 (13) | 54 (21) | 21 (9) | 1 (18) | 79 (16) | 26 (13) | 7 (21) | 281 (11) | 123 (10) | 17 (12) |  |

### Suppl. Table 28 Breakdown of self-reported level of understanding of COVID-19 by country

Values in cells are n (weighted %) of respondents who replied ‘yes’.

| Variable and categories | Thailand | Malaysia | UK | Italy | Slovenia | Total | P-value |
| --- | --- | --- | --- | --- | --- | --- | --- |
|  | N=1,476 | N=827 | N=1,009 | N=712 | N=1,034 | N=5,058 | <0.001 |
| High/very high/expert level understanding | 965 (63) | 435 (51) | 647 (59) | 368 (47) | 713 (66) | 3,128 (59) |  |
| Some understanding | 459 (33) | 359 (38) | 336 (38) | 324 (50) | 279 (30) | 1,757 (36) |  |
| A little/none at all | 52 (4) | 33 (11) | 26 (4) | 20 (3) | 42 (4) | 173 (5) |  |

### Suppl. Table 29 Breakdown of self-reported level of understanding of COVID-19 by demographic characteristics

H = high/very high/expert level; S = some; N = a little/none at all. Values in cells are n (weighted %) of respondents who replied ‘yes’.

| Variable and categories | Thailand | | | | Malaysia | | | | UK | | | | Italy | | | | Slovenia | | | | Total | | | |
| --- | --- | --- | --- | --- | --- | --- | --- | --- | --- | --- | --- | --- | --- | --- | --- | --- | --- | --- | --- | --- | --- | --- | --- | --- |
| Self-reported understanding of COVID-19 | **H** | **S** | **N** | **H** | | **S** | **N** | **H** | | **S** | **N** | **H** | | **S** | **N** | **H** | | **S** | **N** | **H** | | **S** | **N** | **P-value (for total)** |
| Gender |  |  |  |  | |  |  |  | |  |  |  | |  |  |  | |  |  |  | |  |  | 0.058 |
| Male | 458 (65) | 224 (31) | 22 (4) | 153 (55) | | 130 (30) | 15 (15) | 280 (61) | | 134 (35) | 12 (4) | 130 (51) | | 87 (46) | 5 (3) | 269 (64) | | 84 (31) | 13 (5) | 1,290 (60) | | 659 (34) | 67 (6) |  |
| Female | 504 (61) | 232 (35) | 30 (4) | 280 (47) | | 228 (46) | 17 (7) | 358 (56) | | 200 (40) | 14 (3) | 238 (44) | | 237 (53) | 15 (3) | 439 (68) | | 194 (29) | 29 (3) | 1,819 (57) | | 1,091 (39) | 105 (4) |  |
| Other/prefer not to say | 3 (50) | 3 (50) | 0 (0) | 2 (50) | | 1 (25) | 1 (25) | 9 (82) | | 2 (18) | 0 (0) |  | |  |  | 5 (83) | | 1 (17) | 0 (0) | 19 (70) | | 7 (26) | 1 (4) |  |
| Age group |  |  |  |  | |  |  |  | |  |  |  | |  |  |  | |  |  |  | |  |  | 0.033 |
| 18-34 | 143 (62) | 69 (34) | 11 (4) | 170 (48) | | 167 (48) | 13 (9) | 74 (44) | | 58 (48) | 8 (8) | 119 (39) | | 143 (57) | 10 (5) | 186 (59) | | 106 (35) | 16 (6) | 692 (52) | | 543 (41) | 58 (6) |  |
| 35-64 | 746 (62) | 371 (35) | 35 (3) | 244 (54) | | 179 (32) | 19 (14) | 411 (67) | | 193 (32) | 12 (2) | 220 (54) | | 153 (42) | 10 (4) | 492 (69) | | 158 (27) | 26 (5) | 2,113 (62) | | 1,054 (33) | 102 (5) |  |
| 65+ | 76 (68) | 19 (25) | 6 (7) | 21 (52) | | 13 (42) | 1 (6) | 162 (59) | | 85 (39) | 6 (2) | 29 (42) | | 28 (58) | 0 (0) | 35 (68) | | 15 (32) | 0 (0) | 323 (60) | | 160 (38) | 13 (3) |  |
| Education level |  |  |  |  | |  |  |  | |  |  |  | |  |  |  | |  |  |  | |  |  | <0.001 |
| Primary or lower/secondary | 537 (60) | 341 (36) | 31 (4) | 42 (51) | | 30 (36) | 10 (13) | 140 (52) | | 101 (44) | 6 (4) | 92 (43) | | 114 (53) | 11 (4) | 124 (63) | | 67 (33) | 11 (4) | 935 (56) | | 653 (39) | 69 (6) |  |
| Tertiary | 428 (74) | 118 (22) | 21 (4) | 393 (51) | | 329 (46) | 23 (3) | 507 (64) | | 235 (32) | 20 (3) | 276 (58) | | 210 (41) | 9 (2) | 589 (71) | | 212 (26) | 31 (3) | 2,193 (66) | | 1,104 (31) | 104 (3) |  |
| Healthcare worker status |  |  |  |  | |  |  |  | |  |  |  | |  |  |  | |  |  |  | |  |  | 0.001 |
| Healthcare worker | 172 (72) | 59 (26) | 8 (3) | 128 (49) | | 79 (50) | 6 (1) | 90 (76) | | 24 (21) | 4 (3) | 45 (67) | | 18 (29) | 1 (4) | 291 (78) | | 44 (21) | 6 (1) | 726 (70) | | 224 (28) | 25 (2) |  |
| Non-healthcare worker | 793 (61) | 400 (33) | 44 (4) | 307 (52) | | 280 (35) | 27 (13) | 557 (57) | | 312 (39) | 22 (4) | 323 (46) | | 306 (50) | 19 (3) | 422 (63) | | 235 (32) | 36 (5) | 2,402 (57) | | 1,533 (38) | 148 (5) |  |

### Suppl. Table 30 Breakdown of self-reported understanding of public health measures by self-reported level of understanding of COVID-19

(H = high/very high/expert level; S = some; N = a little/none at all). Values in cells are n (weighted %) of respondents who replied ‘yes’.

| Variable and categories | Thailand | | | Malaysia | | | UK | | | Italy | | | Slovenia | | | Total | | | |
| --- | --- | --- | --- | --- | --- | --- | --- | --- | --- | --- | --- | --- | --- | --- | --- | --- | --- | --- | --- |
| Self-reported level of understanding of COVID-19 | **H** | **S** | **N** | **H** | **S** | **N** | **H** | **S** | **N** | **H** | **S** | **N** | **H** | **S** | **N** | **H** | **S** | **N** | **P-value** |
| How would you rate your level of understanding of the current quarantine/isolation/social distancing requirements for  COVID-19? | N=965 | N=459 | N=52 | N=435 | N=359 | N=33 | N=647 | N=336 | N=26 | N=368 | N=324 | N=20 | N=713 | N=279 | N=42 | N=3,128 | N=1,757 | N=173 | <0.001 |
| H | 855 (89) | 116 (23) | 19 (24) | 399 (89) | 193 (52) | 9 (21) | 532 (81) | 182 (57) | 8 (21) | 338 (93) | 213 (71) | 7 (36) | 652 (89) | 212 (59) | 24 (46) | 2,776 (88) | 916 (50) | 67 (27) |  |
| S | 102 (10) | 323 (71) | 11 (12) | 31 (7) | 157 (39) | 15 (52) | 98 (15) | 129 (35) | 11 (46) | 22 (5) | 106 (28) | 10 (38) | 50 (10) | 55 (32) | 12 (44) | 303 (10) | 770 (43) | 59 (39) |  |
| N | 8 (1) | 20 (6) | 22 (64) | 5 (4) | 9 (9) | 9 (27) | 17 (4) | 25 (8) | 7 (33) | 8 (2) | 5 (1) | 3 (26) | 11 (1) | 12 (9) | 6 (10) | 49 (2) | 71 (6) | 47 (34) |  |

### Suppl. Table 31 What were the three most common ways people received communication on COVID-19, and what are the three most preferred ways to receive COVID-19 communications? Breakdown by country

Values in cells are n (weighted %) of respondents who replied ‘yes’.

| Variable and categories | Thailand | Malaysia | UK | Italy | Slovenia | Total | P-value |
| --- | --- | --- | --- | --- | --- | --- | --- |
|  | N=1,476 | N=827 | N=1,009 | N=712 | N=1,034 | N=5,058 |  |
| How do/did you receive information about COVID-19? |  |  |  |  |  |  |  |
| Face-to-face (e.g. doctors or health workers) | 1,096 (78) | 275 (19) | 155 (15) | 276 (32) | 413 (34) | 2,215 (40) | <0.001 |
| Traditional media (TV, radio, newspapers) | 1,407 (95) | 795 (93) | 940 (93) | 650 (85) | 994 (95) | 4,786 (93) | 0.012 |
| Print materials (leaflets, brochures) | 803 (55) | 256 (32) | 403 (36) | 119 (23) | 479 (43) | 2,060 (40) | <0.001 |
| Online (websites, email) | 1,101 (69) | 779 (90) | 918 (89) | 651 (88) | 964 (87) | 4,413 (83) | <0.001 |
| Social media and messenger apps | 1,279 (83) | 786 (95) | 773 (77) | 528 (75) | 731 (66) | 4,097 (79) | <0.001 |
| Government/institution’s web page | 1,134 (74) | 682 (75) | 698 (70) | 580 (79) | 784 (60) | 3,878 (71) | <0.001 |
| WHO web page | 367 (20) | 550 (56) | 380 (36) | 334 (39) | 397 (30) | 2,028 (34) | <0.001 |
| How would you prefer to receive information about COVID-19? |  |  |  |  |  |  |  |
| Face-to-face (e.g doctors or health workers) | 1,200 (83) | 417 (44) | 361 (36) | 584 (77) | 577 (55) | 3,139 (61) | <0.001 |
| Traditional media (TV, radio, newspapers) | 1,347 (90) | 759 (91) | 648 (64) | 467 (62) | 806 (76) | 4,027 (78) | <0.001 |
| Print materials | 893 (63) | 340 (40) | 418 (41) | 149 (29) | 481 (52) | 2,281 (48) | <0.001 |
| Online (websites, email) | 1,105 (71) | 742 (88) | 812 (75) | 473 (71) | 856 (79) | 3,988 (76) | <0.001 |
| Social media and messenger apps | 1,245 (82) | 659 (85) | 330 (31) | 292 (50) | 470 (50) | 2,996 (61) | <0.001 |
| Government/institution’s web page | 1,181 (77) | 731 (86) | 741 (74) | 605 (77) | 845 (71) | 4,103 (77) | 0.009 |
| WHO web page | 586 (36) | 703 (82) | 609 (58) | 531 (64) | 678 (55) | 3,107 (56) | <0.001 |

### Suppl. Table 32 What were the three most common ways people received communications on COVID-19, and what are the three most preferred ways to receive COVID-19 communications? Breakdown by country and gender

M = male; F = female; O = other/prefer not to say. Values in cells are n (weighted %) of respondents who replied ‘yes’.

| Variable and categories | Thailand | | | Malaysia | | | UK | | | Italy | | | Slovenia | | | Total | | | |
| --- | --- | --- | --- | --- | --- | --- | --- | --- | --- | --- | --- | --- | --- | --- | --- | --- | --- | --- | --- |
| Gender | **M** | **F** | **O** | **M** | **F** | **O** | **M** | **F** | **O** | **M** | **F** | **O** | **M** | **F** | **O** | **M** | **F** | **O** | **P-value**  **(for total**  **M vs F)** |
|  | N=704 | N=766 | N=6 | N=298 | N=525 | N=4 | N=426 | N=572 | N=11 | N=222 | N=490 | N=0 | N=366 | N=662 | N=6 | N=2,016 | N=3,015 | N=27 |  |
| How do/did you receive information about COVID-19? |  |  |  |  |  |  |  |  |  |  |  |  |  |  |  |  |  |  |  |
| Face-to-face | 563 (81) | 529 (75) | 4 (67) | 93 (17) | 180 (21) | 2 (50) | 68 (16) | 84 (14) | 3 (27) | 82 (29) | 194 (34) |  | 126 (31) | 285 (37) | 2 (33) | 932 (40) | 1,272 (41) | 11 (41) | 0.591 |
| Traditional media (TV, radio, newspapers) | 669 (94) | 732 (96) | 6 (100) | 284 (92) | 507 (93) | 4 (100) | 390 (92) | 539 (95) | 11 (100) | 199 (82) | 451 (88) |  | 353 (98) | 635 (93) | 6 (100) | 1,895 (92) | 2,864 (94) | 27 (100) | 0.468 |
| Print materials (leaflets, brochures) | 398 (54) | 402 (56) | 3 (50) | 94 (37) | 162 (26) | 0 (0) | 171 (37) | 227 (36) | 5 (45) | 31 (27) | 88 (20) |  | 168 (44) | 307 (41) | 4 (67) | 862 (42) | 1,186 (39) | 12 (44) | 0.265 |
| Online (websites, email) | 509 (69) | 586 (69) | 6 (100) | 281 (92) | 495 (89) | 3 (75) | 379 (87) | 528 (91) | 11 (100) | 201 (85) | 450 (90) |  | 336 (84) | 622 (90) | 6 (100) | 1,706 (82) | 2,681 (84) | 26 (96) | 0.332 |
| Social media and messenger apps | 595 (84) | 678 (82) | 6 (100) | 281 (96) | 502 (94) | 3 (75) | 312 (74) | 450 (79) | 11 (100) | 154 (70) | 374 (80) |  | 256 (66) | 470 (67) | 5 (83) | 1,598 (78) | 2,474 (80) | 25 (93) | 0.589 |
| Government/institution’s web page | 540 (73) | 589 (74) | 5 (83) | 246 (80) | 432 (69) | 4 (100) | 282 (69) | 409 (71) | 7 (64) | 170 (74) | 410 (83) |  | 260 (59) | 518 (61) | 6 (100) | 1,498 (71) | 2,358 (71) | 22 (81) | 0.881 |
| WHO web page | 150 (18) | 214 (22) | 3 (50) | 173 (52) | 374 (60) | 3 (75) | 136 (34) | 239 (39) | 5 (45) | 81 (27) | 253 (50) |  | 108 (26) | 286 (33) | 3 (50) | 648 (30) | 1,366 (38) | 14 (52) | 0.003 |
| How would you prefer to receive information about COVID-19? |  |  |  |  |  |  |  |  |  |  |  |  |  |  |  |  |  |  |  |
| Face-to-face | 594 (85) | 603 (82) | 3 (50) | 146 (39) | 270 (50) | 1 (25) | 163 (36) | 195 (37) | 3 (27) | 171 (75) | 413 (79) |  | 182 (53) | 389 (57) | 6 (100) | 1,256 (59) | 1,870 (63) | 13 (48) | 0.209 |
| Traditional media (TV, radio, newspapers) | 644 (89) | 697 (91) | 6 (100) | 267 (91) | 488 (92) | 4 (100) | 278 (66) | 365 (63) | 5 (45) | 134 (57) | 333 (67) |  | 274 (76) | 530 (77) | 2 (33) | 1,597 (77) | 2,413 (79) | 17 (63) | 0.395 |
| Print materials | 446 (65) | 442 (61) | 5 (83) | 115 (39) | 223 (41) | 2 (50) | 177 (41) | 237 (41) | 4 (36) | 46 (33) | 103 (25) |  | 165 (53) | 314 (51) | 2 (33) | 949 (49) | 1,319 (47) | 13 (48) | 0.408 |
| Online (websites, email) | 516 (70) | 583 (71) | 6 (100) | 269 (92) | 469 (83) | 4 (100) | 334 (71) | 470 (78) | 8 (73) | 151 (72) | 322 (70) |  | 290 (74) | 561 (84) | 5 (83) | 1,560 (75) | 2,405 (77) | 23 (85) | 0.403 |
| Social media and messenger apps | 589 (84) | 650 (80) | 6 (100) | 239 (85) | 416 (87) | 4 (100) | 134 (29) | 195 (34) | 1 (9) | 88 (52) | 204 (48) |  | 161 (43) | 307 (57) | 2 (33) | 1,211 (60) | 1,772 (63) | 13 (48) | 0.364 |
| Government/institution’s web page | 575 (78) | 601 (75) | 5 (83) | 270 (93) | 457 (79) | 4 (100) | 293 (69) | 440 (78) | 8 (73) | 181 (73) | 424 (82) |  | 278 (64) | 561 (77) | 6 (100) | 1,597 (75) | 2,483 (78) | 23 (85) | 0.335 |
| WHO web page | 248 (36) | 334 (36) | 4 (67) | 242 (80) | 457 (83) | 4 (100) | 234 (54) | 370 (62) | 5 (45) | 143 (54) | 388 (74) |  | 209 (49) | 466 (60) | 3 (50) | 1,076 (52) | 2,015 (59) | 16 (59) | 0.020 |

### Suppl. Table 33 What were the three most common ways people received communications on COVID-19, and what are the three most preferred ways to receive COVID-19 communications? Breakdown by country and age group

Values in cells are n (weighted %) of respondents who replied ‘yes’.

| Variable and categories | Thailand | | | Malaysia | | | UK | | | Italy | | | Slovenia | | | Total | | | |
| --- | --- | --- | --- | --- | --- | --- | --- | --- | --- | --- | --- | --- | --- | --- | --- | --- | --- | --- | --- |
| Age group | **18-34** | **35-64** | **65+** | **18-34** | **35-64** | **65+** | **18-34** | **35-64** | **65+** | **18-34** | **35-64** | **65+** | **18-34** | **35-64** | **65+** | **18-34** | **35-64** | **65+** | **P-value**  **(for total)** |
|  | N=223 | N=1,152 | N=101 | N=350 | N=442 | N=35 | N=140 | N=616 | N=253 | N=272 | N=383 | N=57 | N=308 | N=676 | N=50 | N=1,293 | N=3,269 | N=496 |  |
| How do/did you receive information about COVID-19? |  |  |  |  |  |  |  |  |  |  |  |  |  |  |  |  |  |  |  |
| Face-to-face | 125 (68) | 892 (82) | 79 (82) | 141 (20) | 124 (16) | 10 (23) | 25 (17) | 107 (17) | 23 (8) | 112 (37) | 152 (34) | 12 (23) | 111 (32) | 282 (30) | 20 (48) | 514 (37) | 1,557 (42) | 144 (40) | 0.424 |
| Traditional media (TV, radio, newspapers) | 210 (94) | 1,099 (95) | 98 (96) | 337 (89) | 424 (95) | 34 (100) | 130 (93) | 567 (92) | 243 (97) | 247 (92) | 352 (90) | 51 (70) | 299 (98) | 647 (96) | 48 (91) | 1,223 (93) | 3,089 (94) | 474 (90) | 0.336 |
| Print materials (leaflets, brochures) | 107 (54) | 652 (59) | 44 (44) | 104 (31) | 146 (35) | 6 (20) | 34 (22) | 258 (40) | 111 (43) | 34 (12) | 71 (19) | 14 (41) | 140 (45) | 319 (46) | 20 (31) | 419 (37) | 1,446 (43) | 195 (38) | 0.106 |
| Online (websites, email) | 199 (84) | 853 (71) | 49 (35) | 328 (86) | 418 (94) | 33 (91) | 129 (89) | 575 (92) | 214 (82) | 242 (90) | 358 (89) | 51 (82) | 289 (93) | 632 (91) | 43 (74) | 1,187 (87) | 2,836 (85) | 390 (69) | <0.001 |
| Social media and messenger apps | 206 (91) | 1,008 (86) | 65 (55) | 329 (93) | 424 (98) | 33 (91) | 104 (76) | 485 (78) | 184 (74) | 214 (79) | 274 (73) | 40 (77) | 243 (80) | 462 (70) | 26 (42) | 1,096 (86) | 2,653 (81) | 348 (63) | <0.001 |
| Government/institution’s web page | 166 (73) | 902 (78) | 66 (61) | 298 (71) | 360 (81) | 24 (61) | 108 (77) | 459 (74) | 131 (53) | 219 (73) | 318 (81) | 43 (78) | 226 (68) | 528 (71) | 30 (29) | 1,017 (72) | 2,567 (77) | 294 (54) | <0.001 |
| WHO web page | 100 (31) | 256 (19) | 11 (6) | 260 (62 | 274 (53) | 16 (39) | 60 (45) | 271 (40) | 49 (18) | 129 (39) | 176 (38) | 29 (42) | 127 (39) | 255 (30) | 15 (19) | 676 (44) | 1,232 (33) | 120 (22) | <0.001 |
| How would you prefer to receive information about COVID-19? |  |  |  |  |  |  |  |  |  |  |  |  |  |  |  |  |  |  |  |
| Face-to-face | 152 (77) | 965 (87) | 83 (84) | 198 (53) | 203 (34) | 16 (53) | 48 (33) | 218 (37) | 95 (39) | 230 (78) | 313 (80) | 41 (71) | 187 (57) | 365 (53) | 25 (59) | 815 (59) | 2,064 (61) | 260 (62) | 0.785 |
| Traditional media (TV, radio, newspapers) | 194 (85) | 1,056 (91) | 97 (93) | 327 (90) | 402 (91) | 30 (99) | 89 (65) | 396 (64) | 163 (64) | 179 (60) | 247 (58) | 41 (72) | 228 (73) | 534 (75) | 44 (83) | 1,017 (78) | 2,635 (78) | 375 (80) | 0.712 |
| Print materials | 118 (64) | 720 (65) | 55 (54) | 143 (41) | 179 (37) | 18 (45) | 40 (27) | 256 (44) | 122 (52) | 43 (15) | 88 (24) | 18 (50) | 149 (50) | 308 (48) | 24 (63) | 493 (44) | 1,551 (48) | 237 (54) | 0.073 |
| Online (websites, email) | 187 (83) | 867 (73) | 51 (41) | 312 (87) | 399 (91) | 31 (77) | 98 (59) | 522 (84) | 192 (74) | 180 (74) | 253 (68) | 40 (75) | 250 (79) | 567 (83) | 39 (71) | 1,027 (78) | 2,608 (79) | 353 (66) | <0.001 |
| Social media and messenger apps | 196 (91) | 986 (85) | 63 (55) | 285 (88) | 349 (86) | 25 (75) | 34 (21) | 219 (37) | 77 (31) | 105 (38) | 156 (48) | 31 (65) | 134 (48) | 317 (51) | 19 (49) | 754 (64) | 2,027 (64) | 215 (52) | 0.005 |
| Government/institution’s web page | 177 (79) | 936 (80) | 68 (60) | 323 (93) | 381 (81) | 27 (82) | 108 (71) | 468 (77) | 165 (71) | 235 (83) | 325 (82) | 45 (65) | 252 (75) | 557 (76) | 36 (56) | 1,095 (81) | 2,667 (79) | 341 (64) | <0.001 |
| WHO web page | 145 (55) | 415 (31) | 26 (20) | 320 (92) | 357 (72) | 26 (77) | 98 (65) | 387 (60) | 124 (46) | 226 (79) | 266 (64) | 39 (53) | 231 (73) | 427 (59) | 20 (26) | 1,020 (72) | 1,852 (53) | 235 (39) | <0.001 |

### Suppl. Table 34 What were the three most common ways people received communications on COVID-19, and what are the three most preferred ways to receive COVID-19 communications? Breakdown by country and education level

P/S = primary or lower/secondary education; T = tertiary education. Values in cells are n (weighted %) of respondents who replied ‘yes’.

| Variable and categories | Thailand | | Malaysia | | UK | | Italy | | Slovenia | | Total | | |
| --- | --- | --- | --- | --- | --- | --- | --- | --- | --- | --- | --- | --- | --- |
| Education level | **P/S** | **T** | **P/S** | **T** | **P/S** | **T** | **P/S** | **T** | **P/S** | **T** | **P/S** | **T** | **P-value**  **(for total)** |
|  | N=909 | N=567 | N=82 | N=745 | N=247 | N=762 | N=217 | N=495 | N=202 | N=832 | N=1,657 | N=3,401 |  |
| How do/did you receive information about COVID-19? |  |  |  |  |  |  |  |  |  |  |  |  |  |
| Face-to-face | 781 (83) | 315 (55) | 13 (14) | 262 (37) | 32 (14) | 123 (16) | 72 (28) | 204 (39) | 48 (29) | 365 (43) | 946 (43) | 1,269 (35) | <0.001 |
| Traditional media (TV, radio, newspapers) | 865 (95) | 542 (95) | 76 (92) | 719 (97) | 234 (95) | 706 (92) | 192 (82) | 458 (93) | 196 (95) | 798 (96) | 1,563 (92) | 3,223 (94) | 0.155 |
| Print materials (leaflets, brochures) | 547 (57) | 256 (45) | 26 (32) | 230 (31) | 90 (34) | 313 (38) | 39 (26) | 80 (16) | 91 (40) | 388 (47) | 793 (42) | 1,267 (38) | 0.062 |
| Online (websites, email) | 605 (65) | 496 (87) | 74 (89) | 705 (95) | 212 (85) | 706 (93) | 190 (85) | 461 (93) | 179 (83) | 785 (94) | 1,260 (79) | 3,153 (92) | <0.001 |
| Social media and messenger apps | 757 (81) | 522 (91) | 78 (95) | 708 (94) | 196 (79) | 577 (75) | 173 (78) | 355 (70) | 150 (65) | 581 (68) | 1,354 (80) | 2,743 (77) | 0.146 |
| Government/institution’s web page | 689 (73) | 445 (78) | 59 (73) | 623 (85) | 171 (70) | 527 (71) | 166 (77) | 414 (81) | 123 (49) | 661 (78) | 1,208 (69) | 2,670 (77) | <0.001 |
| WHO web page | 139 (15) | 228 (42) | 44 (53) | 506 (67) | 68 (30) | 312 (42) | 84 (35) | 250 (49) | 59 (24) | 338 (39) | 394 (29) | 1,634 (44) | <0.001 |
| How would you prefer to receive information about COVID-19? |  |  |  |  |  |  |  |  |  |  |  |  |  |
| Face-to-face | 806 (87) | 394 (68) | 36 (42) | 381 (53) | 104 (39) | 257 (34) | 170 (75) | 414 (81) | 111 (56) | 466 (54) | 1,227 (65) | 1,912 (53) | <0.001 |
| Traditional media (TV, radio, newspapers) | 830 (90) | 517 (90) | 75 (91) | 684 (92) | 149 (63) | 499 (66) | 133 (60) | 334 (68) | 145 (74) | 661 (80) | 1,332 (79) | 2,695 (76) | 0.100 |
| Print materials | 608 (66) | 285 (49) | 35 (40) | 305 (40) | 126 (47) | 292 (37) | 48 (32) | 101 (21) | 105 (57) | 376 (45) | 922 (52) | 1,359 (39) | <0.001 |
| Online (websites, email) | 632 (68) | 473 (82) | 71 (87) | 671 (90) | 186 (68) | 626 (81) | 156 (74) | 317 (64) | 160 (77) | 696 (83) | 1,205 (74) | 2,783 (80) | <0.001 |
| Social media and messenger apps | 753 (81) | 492 (86) | 72 (87) | 587 (79) | 90 (32) | 240 (31) | 106 (55) | 186 (38) | 111 (55) | 359 (42) | 1,132 (67) | 1,864 (49) | <0.001 |
| Government/institution’s web page | 711 (75) | 470 (83) | 69 (86) | 662 (90) | 194 (75) | 547 (72) | 173 (74) | 432 (86) | 138 (63) | 707 (84) | 1,285 (75) | 2,818 (81) | 0.001 |
| WHO web page | 246 (30) | 340 (61) | 66 (81) | 637 (85) | 122 (50) | 487 (65) | 149 (60) | 382 (74) | 123 (49) | 555 (64) | 706 (50) | 2,401 (67) | <0.001 |

### Suppl. Table 35 Most prevalent topic areas with unclear or conflicting COVID-19 information, and most prevalent ‘fake news’, breakdown by country

Values in cells are n (weighted %) of respondents who replied ‘yes’.

| Variable and categories | Thailand | Malaysia | UK | Italy | Slovenia | Total | P-value |
| --- | --- | --- | --- | --- | --- | --- | --- |
|  | N=1,476 | N=827 | N=1,009 | N=712 | N=1,034 | N=5,058 |  |
| Have you seen any unclear or conflicting information about COVID-19 in the last month? |  |  |  |  |  |  |  |
| Ways to avoid the infection | 564 (36) | 409 (47) | 679 (68) | 410 (64) | 682 (64) | 2,744 (54) | <0.001 |
| Symptoms of COVID-19 | 568 (36) | 353 (42) | 590 (62) | 328 (44) | 494 (44) | 2,333 (45) | <0.001 |
| What to do in case of symptoms | 506 (34) | 295 (37) | 438 (43) | 293 (45) | 435 (42) | 1,967 (40) | 0.058 |
| Social distancing guidance | 490 (33) | 292 (42) | 568 (56) | 314 (42) | 559 (51) | 2,223 (44) | <0.001 |
| Quarantine/isolation | 529 (36) | 314 (39) | 547 (54) | 292 (41) | 559 (52) | 2,241 (44) | <0.001 |
| Penalties if disobey restrictions | 614 (41) | 384 (42) | 620 (60) | 378 (52) | 508 (45) | 2,504 (47) | <0.001 |
| Risks in case of infection | 527 (34) | 327 (37) | 542 (54) | 330 (49) | 493 (46) | 2,219 (43) | <0.001 |
| Numbers of coronavirus cases/deaths related to COVID-19 | 563 (37) | 284 (47) | 741 (72) | 457 (66) | 463 (46) | 2,508 (52) | <0.001 |
| Government support schemes (e.g. financial) | 779 (51) | 432 (53) | 438 (46) | 492 (69) | 572 (51) | 2,713 (53) | <0.001 |
| Testing | 531 (34) | 376 (39) | 734 (72) | 520 (72) | 534 (49) | 2,695 (51) | <0.001 |
| Travel restrictions (e.g. curfew, restricted hours of movement) | 520 (33) | 407 (43) | 641 (62) | 382 (55) | 533 (45) | 2,483 (46) | <0.001 |
| Have you come across news about the following COVID-19 topics that seemed fake to you? |  |  |  |  |  |  |  |
| General spread of fear | 668 (42) | 606 (70) | 693 (72) | 382 (58) | 771 (69) | 3,120 (60) | <0.001 |
| Coronavirus as an engineered modified virus | 543 (32) | 613 (65) | 819 (81) | 613 (82) | 864 (75) | 3,452 (63) | <0.001 |
| Minimisation of risks | 440 (27) | 416 (39) | 579 (55) | 540 (69) | 731 (62) | 2,706 (48) | <0.001 |
| Numbers of infected/deceased people | 512 (33) | 400 (47) | 615 (61) | 475 (75) | 574 (54) | 2,576 (51) | <0.001 |
| Unreasonable health recommendations | 517 (32) | 545 (55) | 574 (57) | 385 (50) | 650 (60) | 2,671 (49) | <0.001 |
| Pharmaceutical conspiracy | 490 (32) | 440 (50) | 525 (54) | 489 (63) | 673 (61) | 2,617 (49) | <0.001 |
| Home-made recipes to make sanitizer products | 538 (32) | 573 (61) | 557 (56) | 516 (70) | 603 (51) | 2,787 (51) | <0.001 |
| Alternative drugs/cure | 537 (33) | 581 (60) | 697 (67) | 444 (58) | 612 (51) | 2,871 (51) | <0.001 |
| Fear toward products coming from infected countries | 458 (29) | 549 (63) | 483 (49) | 425 (56) | 519 (48) | 2,434 (46) | <0.001 |

### Suppl. Table 36 Most prevalent topic areas with unclear or conflicting COVID-19 information, and most prevalent ‘fake news’, breakdown by country and education level

P/S = primary or lower/secondary education; T = tertiary education. Values in cells are n (weighted %) of respondents who replied ‘yes’.

| Variable and categories | Thailand | | Malaysia | | UK | | Italy | | Slovenia | | Total | | |
| --- | --- | --- | --- | --- | --- | --- | --- | --- | --- | --- | --- | --- | --- |
| Education level | **P/S** | **T** | **P/S** | **T** | **P/S** | **T** | **P/S** | **T** | **P/S** | **T** | **P/S** | **T** | **P-value**  **(for total)** |
|  | N=909 | N=567 | N=82 | N=745 | N=247 | N=762 | N=217 | N=495 | N=202 | N=832 | N=1,657 | N=3,401 |  |
| Have you seen any unclear or conflicting information about COVID-19 in the last month? |  |  |  |  |  |  |  |  |  |  |  |  |  |
| Ways to avoid the infection | 276 (33) | 288 (51) | 37 (46) | 372 (49) | 153 (66) | 526 (69) | 119 (65) | 291 (60) | 125 (63) | 557 (67) | 710 (50) | 2,034 (62) | <0.001 |
| Symptoms | 268 (33) | 300 (53) | 36 (43) | 317 (41) | 146 (65) | 444 (59) | 94 (42) | 234 (48) | 96 (44) | 398 (46) | 640 (42) | 1,693 (51) | <0.001 |
| What to do in case of symptoms | 245 (31) | 261 (47) | 32 (38) | 263 (36) | 96 (42) | 342 (44) | 94 (46) | 199 (43) | 80 (42) | 355 (41) | 547 (38) | 1,420 (43) | 0.026 |
| Social distancing guidance | 249 (31) | 241 (42) | 36 (44) | 256 (34) | 113 (51) | 455 (61) | 92 (41) | 222 (46) | 109 (50) | 450 (53) | 599 (41) | 1,624 (51) | <0.001 |
| Quarantine/isolation | 278 (34) | 251 (45) | 32 (40) | 282 (38) | 123 (51) | 424 (56) | 84 (41) | 208 (43) | 102 (50) | 457 (55) | 619 (41) | 1,622 (50) | <0.001 |
| Penalties if disobey restrictions | 315 (38) | 299 (52) | 34 (40) | 350 (48) | 143 (56) | 477 (62) | 103 (50) | 275 (56) | 101 (44) | 407 (47) | 696 (44) | 1,808 (55) | <0.001 |
| Risks in case of infection | 257 (31) | 270 (49) | 32 (36) | 295 (39) | 127 (54) | 415 (55) | 105 (50) | 225 (46) | 93 (45) | 400 (47) | 614 (40) | 1,605 (49) | <0.001 |
| Numbers of coronavirus cases/deaths related to COVID-19 | 284 (33) | 279 (52) | 42 (50) | 242 (33) | 172 (70) | 569 (74) | 140 (67) | 317 (65) | 107 (50) | 356 (41) | 745 (49) | 1,763 (56) | 0.001 |
| Government support schemes (e.g. financial) | 402 (47) | 377 (69) | 44 (54) | 388 (52) | 103 (50) | 335 (43) | 138 (69) | 354 (71) | 108 (50) | 464 (54) | 795 (52) | 1,918 (55) | 0.257 |
| Testing | 258 (31) | 273 (49) | 31 (38) | 345 (45) | 161 (68) | 573 (75) | 145 (70) | 375 (76) | 95 (48) | 439 (51) | 690 (46) | 2,005 (62) | <0.001 |
| Travel restrictions (e.g. curfew, restricted hours of movement) | 248 (30) | 272 (49) | 36 (42) | 371 (49) | 142 (59) | 499 (65) | 112 (55) | 270 (55) | 96 (41) | 437 (51) | 634 (42) | 1,849 (56) | <0.001 |
| Have you come across news about the following COVID-19 topics that seemed fake to you? |  |  |  |  |  |  |  |  |  |  |  |  |  |
| General spread of fear | 308 (37) | 360 (64) | 56 (69) | 550 (73) | 182 (76) | 511 (68) | 116 (60) | 266 (54) | 147 (66) | 624 (74) | 809 (57) | 2,311 (67) | <0.001 |
| Coronavirus as an engineered modified virus | 209 (26) | 334 (61) | 52 (62) | 561 (76) | 193 (80) | 626 (82) | 174 (80) | 439 (89) | 156 (70) | 708 (84) | 784 (56) | 2,668 (79) | <0.001 |
| Minimisation of risks | 178 (23) | 262 (47) | 31 (36) | 385 (51) | 128 (52) | 451 (59) | 141 (63) | 399 (81) | 122 (56) | 609 (71) | 600 (41) | 2,106 (62) | <0.001 |
| Numbers of infected/deceased people | 231 (29) | 281 (51) | 40 (47) | 360 (49) | 152 (62) | 463 (61) | 153 (719 | 322 (67) | 118 (55) | 456 (54) | 694 (49) | 1,882 (57) | <0.001 |
| Unreasonable health recommendations | 204 (27) | 313 (57) | 45 (52) | 500 (66) | 131 (55) | 443 (59) | 101 (46) | 284 (60) | 122 (58) | 528 (64) | 603 (44) | 2,068 (61) | <0.001 |
| Pharmaceutical conspiracy | 239 (29) | 251 (45) | 41 (49) | 399 (54) | 131 (56) | 394 (52) | 138 (60) | 351 (71) | 125 (58) | 548 (64) | 674 (46) | 1,943 (57) | <0.001 |
| Home-made recipes to make sanitizer products | 230 (27) | 308 (55) | 51 (59) | 522 (69) | 158 (62) | 399 (51) | 149 (68) | 367 (75) | 104 (46) | 499 (59) | 692 (47) | 2,095 (59) | <0.001 |
| Alternative drugs/cure | 240 (28) | 297 (53) | 48 (57) | 533 (71) | 168 (65) | 529 (69) | 125 (55) | 319 (66) | 105 (44) | 507 (61) | 686 (46) | 2,185 (64) | <0.001 |
| Fear toward products coming from infected countries | 197 (25) | 261 (46) | 52 (62) | 497 (67) | 127 (52) | 356 (46) | 126 (55) | 299 (59) | 100 (46) | 419 (51) | 602 (44) | 1,832 (51) | <0.001 |

### Suppl. Table 37 Most prevalent topic areas with unclear or conflicting COVID-19 information, and most prevalent ‘fake news’, breakdown by country and self-reported level of understanding of COVID-19

H = high/very high/expert level; S = some; N = a little/none at all. Values in cells are n (weighted %) of respondents who replied ‘yes’.

| Variable and categories | Thailand | | | Malaysia | | | UK | | | Italy | | | Slovenia | | | Total | | | |
| --- | --- | --- | --- | --- | --- | --- | --- | --- | --- | --- | --- | --- | --- | --- | --- | --- | --- | --- | --- |
| Self-reported level of understanding of COVID-19 | **H** | **S** | **N** | **H** | **S** | **N** | **H** | **S** | **N** | **H** | **S** | **N** | **H** | **S** | **N** | **H** | **S** | **N** | **P-value (for total)** |
|  | N=965 | N=459 | N=52 | N=435 | N=359 | N=33 | N=647 | N=336 | N=26 | N=368 | N=324 | N=20 | N=713 | N=279 | N=42 | N=3,128 | N=1,757 | N=173 |  |
| Have you seen any unclear or conflicting information about COVID-19 in the last month? |  |  |  |  |  |  |  |  |  |  |  |  |  |  |  |  |  |  |  |
| Ways to avoid the infection | 401 (40) | 145 (32) | 18 (19) | 197 (43) | 191 (46) | 21 (63) | 416 (63) | 248 (76) | 15 (53) | 202 (54) | 193 (72) | 15 (73) | 445 (61) | 211 (73) | 26 (53) | 1,661 (51) | 988 (58) | 95 (51) | 0.094 |
| Symptoms of COVID-19 | 400 (40) | 150 (33) | 18 (19) | 170 (36) | 167 (49) | 16 (51) | 363 (58) | 210 (66) | 17 (79) | 147 (31) | 163 (53) | 18 (81) | 312 (40) | 164 (54) | 18 (41) | 1,392 (42) | 854 (50) | 87 (49) | 0.026 |
| What to do in case of symptoms | 361 (37) | 129 (30) | 16 (17) | 134 (34) | 145 (41) | 16 (39) | 272 (39) | 156 (49) | 10 (59) | 138 (34) | 144 (55) | 11 (49) | 285 (37) | 130 (52) | 20 (40) | 1,190 (37) | 704 (44) | 73 (37) | 0.041 |
| Social distancing guidance | 349 (37) | 124 (27) | 17 (19) | 132 (36) | 144 (43) | 16 (62) | 355 (52) | 199 (62) | 14 (70) | 163 (38) | 140 (45) | 11 (65) | 362 (47) | 170 (58) | 27 (64) | 1,361 (42) | 777 (46) | 85 (54) | 0.168 |
| Quarantine/isolation | 379 (39) | 139 (32) | 11 (11) | 153 (33) | 145 (39) | 16 (71) | 338 (49) | 193 (59) | 16 (76) | 148 (39) | 135 (44) | 9 (39) | 372 (50) | 165 (58) | 22 (41) | 1,390 (43) | 777 (46) | 74 (50) | 0.397 |
| Penalties if disobey restrictions | 477 (49) | 126 (28) | 11 (11) | 186 (35) | 180 (46) | 18 (56) | 381 (54) | 225 (68) | 14 (66) | 187 (47) | 180 (56) | 11 (69) | 324 (44) | 162 (48) | 22 (53) | 1,555 (47) | 873 (48) | 76 (47) | 0.906 |
| Risks in case of infection | 381 (38) | 132 (29) | 14 (15) | 152 (29) | 158 (43) | 17 (50) | 337 (50) | 191 (62) | 14 (46) | 158 (43) | 156 (53) | 16 (73) | 312 (46) | 159 (45) | 22 (45) | 1,340 (41) | 796 (46) | 83 (42) | 0.343 |
| Numbers of coronavirus cases/deaths related to COVID-19 | 416 (42) | 134 (29) | 13 (15) | 129 (41) | 137 (50) | 18 (68) | 463 (66) | 261 (81) | 17 (77) | 233 (67) | 214 (66) | 10 (57) | 284 (43) | 156 (53) | 23 (57) | 1,525 (50) | 902 (54) | 81 (54) | 0.276 |
| Government support schemes (e.g. financial) | 583 (60) | 178 (38) | 18 (20) | 208 (46) | 203 (61) | 21 (62) | 269 (40) | 158 (53) | 11 (56) | 248 (67) | 227 (71) | 17 (78) | 372 (48) | 176 (59) | 24 (48) | 1,680 (52) | 942 (55) | 91 (50) | 0.590 |
| Testing | 392 (39) | 124 (29) | 15 (15) | 181 (36) | 179 (46) | 16 (32) | 467 (70) | 249 (74) | 18 (77) | 266 (71) | 239 (71) | 15 (86) | 357 (48) | 154 (55) | 23 (31) | 1,663 (50) | 945 (53) | 87 (39) | 0.108 |
| Travel restrictions (e.g. curfew, restricted hours of movement) | 391 (39) | 118 (25) | 11 (11) | 209 (37) | 178 (46) | 20 (62) | 398 (60) | 228 (71) | 15 (52) | 192 (50) | 176 (58) | 14 (78) | 341 (43) | 167 (50) | 25 (41) | 1,531 (44) | 867 (49) | 85 (47) | 0.356 |
| Have you come across news about the following COVID-19 topics that seemed fake to you? |  |  |  |  |  |  |  |  |  |  |  |  |  |  |  |  |  |  |  |
| General spread of fear | 488 (47) | 158 (36) | 22 (23) | 320 (65) | 266 (80) | 20 (56) | 449 (70) | 228 (73) | 16 (81) | 208 (57) | 163 (59) | 11 (61) | 518 (71) | 222 (65) | 31 (66) | 1,983 (61) | 1,037 (60) | 100 (54) | 0.594 |
| Coronavirus as an engineered modified virus | 390 (37) | 134 (26) | 19 (19) | 327 (71) | 266 (62) | 20 (46) | 532 (83) | 268 (79) | 19 (70) | 320 (87) | 277 (80) | 16 (60) | 598 (80) | 231 (65) | 35 (75) | 2,167 (66) | 1,176 (60) | 109 (49) | 0.007 |
| Minimisation of risks | 305 (30) | 120 (24) | 15 (13) | 222 (38) | 176 (41) | 18 (32) | 377 (56) | 191 (56) | 11 (39) | 277 (64) | 249 (74) | 14 (54) | 510 (64) | 196 (57) | 25 (47) | 1,691 (48) | 932 (49) | 83 (33) | 0.063 |
| Numbers of infected/deceased people | 345 (34) | 148 (33) | 19 (18) | 206 (49) | 174 (48) | 20 (39) | 392 (58) | 207 (66) | 16 (75) | 252 (76) | 214 (75) | 9 (63) | 377 (51) | 172 (62) | 25 (61) | 1,572 (49) | 915 (55) | 89 (45) | 0.105 |
| Unreasonable health recommendations | 387 (36) | 113 (26) | 17 (17) | 286 (54) | 237 (53) | 22 (63) | 375 (55) | 186 (58) | 13 (71) | 211 (57) | 163 (44) | 11 (54) | 440 (59) | 186 (65) | 24 (48) | 1,699 (50) | 885 (47) | 87 (50) | 0.538 |
| Pharmaceutical conspiracy | 358 (36) | 112 (25) | 20 (21) | 238 (53) | 188 (48) | 14 (38) | 355 (55) | 158 (51) | 12 (56) | 266 (69) | 209 (57) | 14 (65) | 453 (61) | 192 (61) | 28 (45) | 1,670 (52) | 859 (46) | 88 (40) | 0.059 |
| Home-made recipes to make sanitizer products | 400 (38) | 122 (24) | 16 (15) | 309 (62) | 241 (62) | 23 (57) | 366 (56) | 179 (55) | 12 (68) | 274 (78) | 227 (62) | 15 (71) | 411 (52) | 170 (51) | 22 (45) | 1,760 (52) | 939 (49) | 88 (48) | 0.390 |
| Alternative drugs/cure | 409 (39) | 112 (24) | 16 (16) | 305 (57) | 257 (75) | 19 (20) | 468 (72) | 214 (62) | 15 (50) | 243 (64) | 188 (52) | 13 (66) | 430 (53) | 159 (45) | 23 (58) | 1,855 (54) | 930 (49) | 86 (33) | 0.004 |
| Fear toward products coming from infected countries | 330 (33) | 109 (23) | 19 (20) | 297 (65) | 234 (68) | 18 (39) | 317 (50) | 155 (48) | 11 (44) | 226 (58) | 187 (55) | 12 (64) | 352 (47) | 145 (49) | 22 (46) | 1,522 (47) | 830 (46) | 82 (39) | 0.456 |
